# Multicenter international cohort study of HA20 reveals novel genetic architecture and phenotypic evolution

**DOI:** 10.64898/2026.01.01.25342105

**Authors:** Tingyan He, Jun Wang, Manuel Carpio Tumba, Shihao Wang, Youyou Luo, Jie Chen, Guomin Li, Zhou Shu, Song Zhang, Deborah L Stone, Yanyan Huang, Qianying Lv, Wen Xiong, Jinbo Wang, Zhongxun Yu, Charlotte Vera Cuff, Elizabeth Kairis, Akuti Kethri, Atif Towheed, Kyr Goyette, Urekha Karri, Jiebiao Wang, Chen Liu, Tina Romeo, Laia Alsina Manrique De Lara, Daniel L Rosenberg, Daniel Clemente, Juan Carlos Lopez Robledillo, Zanhua Rong, Xue Zhao, Lijun Jiang, Juan Carlos Aldave-Becerra, Ana Beatriz Muñoz-Urribarri, Prasad Thomas Oommen, Priscilla Campbell-Stokes, Meifang Zhu, Peng Liu, Li Guo, Yiping Xu, Zihua Yu, Huajuan Tong, Xiaojian Qiu, Yazhi Zhang, Hongbo Chen, Changming Zhang, Junbin Ou, Congcong Liu, Jinxiang Liu, Yunyan Shen, Jianshe Cao, Xinping Zhang, Kangkang Yang, Ying Bao, Zhijuan Li, Jie Cao, Yuanhui Duan, Fujuan Liu, Buyun Shi, Min Sun, Li Ma, Yongxing Chen, Wei Yang, Xu Han, Shuangyue Ma, Jie Luo, Weiyue Gu, Guoliang Yu, Weina Shi, Ruiqin Zhao, Lei Sun, Wenhui Li, Yunfei An, Xuemei Tang, Xiaodong Zhao, Tongxin Han, Jing Ma, Yan Li, Yurong Piao, Fei Sun, Dongfeng Zhang, Meina Yin, Shaoling Zheng, Tianwang Li, Huizhong Niu, Li Lin, Shiyue Mei, Fang Zhou, Sirui Yang, Danlu Li, Mei Yan, Huasong Zeng, Ping Zeng, Wenjie Zheng, Xiaozhong Li, Xiaolin Li, Yuling Liu, Lijuan Huang, Haiguo Yu, Zhidan Fan, Min Shen, Meiping Lu, Zhihao Fang, Casey Allison Rimland, Hongmei Song, Lance W Peterson, Maryam Ali Yousuf Al-Nesf, Sami Aqel, Dalal Sideeg Mudawi, Nikita S Raje, Voytek Slowik, Julia Harris, Brenda Snyder, Selma Scheffler-Mendoza, Marco Yamazaki-Nakashimada, Megan A Cooper, Yu Lung Lau, Kader Cetin Gedik, Wenjie Wang, Xiaochuan Wang, Amanda K Ombrello, Ying Huang, Hai-Lin Wu, Li Sun, Huawei Mao, Xiaomin Yu, Zhihong Liu, Ivona Aksentijevich, Daniel L Kastner, Daniella M Schwartz, Jun Yang, Qing Zhou

## Abstract

**Background:** Haploinsufficiency of A20 (HA20) is an immune dysregulation disorder caused by loss-of-function *TNFAIP3* mutations. This international multicenter study aimed to delineate its clinical spectrum, genetic basis, and natural history.

**Methods:** A cross-sectional, retrospective analysis was conducted in HA20 patients with pathogenic or likely pathogenic *TNFAIP3* variants. Clinical, laboratory and treatment data were assessed. Clustering analysis was applied to evaluate clinical features and disease phenotypes. Patients were stratified by age (< 16 years vs ≥ 16 years), country of origin (China vs U.S.), and gender.

**Results:** A total of 185 patients from 41 clinics across 7 countries were included (median onset 3.3 years). Common clinical features were mucocutaneous involvement (80.5%), recurrent fever (63.3%), gastrointestinal symptoms (58.6%), cytopenias (56.6%), arthritis/arthralgia (46.7%), and recurrent infections (35.5%). Compared with adults and patients from the U.S. cohort, intestinal ulcers were significantly more frequent in children and patients from the Chinese cohort (*P* = 0.002 and *P* < 0.0001, respectively), whereas uveitis was less common in these groups (*P* = 0.001 and *P* < 0.0001, respectively). Hierarchical clustering of clinical disease phenotypes based on Pearson distance identified two major clusters, an autoinflammation-predominant phenotype and an autoimmune-predominant phenotype. The autoinflammation-predominant phenotype was more common in children and patients from the Chinese cohort (*P* = 0.033 and *P* = 0.003, respectively). A total of 89 pathogenic *TNFAIP3* mutations were identified, including 46 novel variants. Large deletions were associated with neurologic disease and developmental delay (*P* = 0.0054 and *P* = 0.0245, respectively); however, there were no clear association between disruptions of specific functional A20 domains and onset age or phenotype. Therapeutically, TNF and IL-1 inhibitors were effective in most patients, with thalidomide and JAK inhibitors used in refractory cases; 51.5% had achieved minimal disease activity at the most recent follow-up.

**Conclusions:** HA20 is a common dominantly inherited immune dysregulation disorder with phenotypic heterogeneity and potential age-dependent evolution. This large international cohort highlights diagnostic and therapeutic strategies to advance evaluation and management of HA20.

## INTRODUCTION

Haploinsufficiency of A20 (HA20) is a monogenic immune dysregulation disease caused by loss-of-function (LOF) mutations in *TNFAIP3.*^1^ HA20 was initially characterized as an early-onset autoinflammatory disease resembling Behçet’s disease (BD).^1^ However, since its initial description, the clinical spectrum of HA20 has greatly expanded.^2, 3^ Patients can present with diverse clinical phenotypes, including classic autoinflammation, systemic or organ-specific autoimmunity, and/or immunodeficiency.^3, 4^ In addition to BD, HA20 patients have been diagnosed with early-onset inflammatory bowel disease (IBD), periodic fever with aphthous pharyngitis and adenitis (PFAPA), systemic juvenile idiopathic arthritis (sJIA), systemic lupus erythematosus (SLE), polyarticular JIA/oligoarticular JIA, autoimmune thyroiditis, autoimmune hepatitis (AIH), autoimmune lymphoproliferative syndrome (ALPS), common variable immunodeficiency (CVID), and many others.^2–15^ While nearly 200 HA20 cases have been described to date, most have been small case reports or series that were subsequently collected and reviewed. This could lead to an inaccurate representation of the natural history due to reporting or ascertainment bias. To date, no large multicenter studies have followed patients over time to systematically describe this expanded clinical spectrum.

In addition to being a clinically heterogeneous disease, HA20 is also characterized by substantial genetic and molecular heterogeneity. A20 is a protein with several important functional domains including the ovarian tumor (OTU) domain and 7 zinc finger domains. Each of these has critical roles in ubiquitin editing including destabilization of linear and K63-linked ubiquitin and ligation of K48-linked ubiquitin.^4^ Together, these functions destabilize and deactivate critical proteins involved in immune activation, apoptosis, and necroptosis. HA20-causal mutations have been reported throughout the *TNFAIP3* gene, and previous studies have suggested that the specific localization of *TNFAIP3* mutations could underlie phenotypic variability.^16, 17^ The genetic landscape of pathogenic *TNFAIP3* mutations and phenotype-genotype correlations remain incompletely characterized.^4^

Another major area of uncertainty concerns the optimal treatment protocol for HA20 patients. To date, patients have been treated with glucocorticoids (GCs), conventional synthetic disease-modifying antirheumatic drugs (csDMARDs), biological DMARDs (bDMARDs), and targeted synthetic DMARDs (tsDMARDs).^3, 18^ Responses to treatment vary greatly among individuals with similar disease phenotypes, even in those carrying identical mutations or affected members from the same family. Follow-up has been limited in some studies – particularly in early descriptions within 2-3 years of the initial disease discovery.^1, 3, 18, 19^ Hence, long-term treatment responses and adverse effects have not yet been explored.

Herein, we described the clinical manifestations, genetic landscape, treatments, and outcomes of 185 patients with HA20 from a large international multicenter cohort.

## METHODS

### Participants

Patients with HA20 were referred to the study from rheumatology and immunology clinics after undergoing genetic testing that revealed a suspected pathogenic variant in *TNFAIP3*. Patients seen at the University of Pittsburgh and National Institutes of Health were evaluated with a comprehensive rheumatologic examination and a standardized set of clinical laboratory studies. The Chinese study was designed as a retrospective cross-sectional study. It was approved by ethics committees at the Shenzhen Children’s Hospital in China (Study KXXSC202403006) and a waiver of written informed consent from the Chinese patients was granted as the research involved minimal risk and used anonymized data. The U.S. study was designed as a prospective longitudinal cohort study, but also collected retrospective data upon enrollment (i.e. cross-sectional) and was approved by the Institutional Review Boards of the University of Pittsburgh (IRB Studies 22060106, 24050013) and National Institutes of Health (Study 94-HG-0105).

### Inclusion and Exclusion Criteria

#### Inclusion criteria

Patients were eligible for inclusion if they met the following criteria, 1. Confirmed HA20 diagnosis established by genetic testing demonstrating either, a heterozygous pathogenic or likely pathogenic variant in *TNFAIP3*, or a heterozygous deletion of 6q23 including *TNFAIP3*, identified by WES, multigene panel testing, chromosomal microarray analysis, or next-generation sequencing–based clinical assays. 2. Novel coding variants were functionally validated using ectopic overexpression of wild-type (WT) versus mutant A20 in HEK293T cells, followed by assessment of A20-mediated suppression of TNF-induced NF-κB activation with a luciferase reporter assay (Cignal, Qiagen). Most variants were tested in WT HEK293T cells, and a subset of variants (n = 9) was tested in both WT and A20-deficient HEK293T cells to confirm LOF. Splice null mutations and complete deletions were considered pathogenic by American College of Medical Genetics (ACMG) standards.^20, 21^

#### Exclusion criteria

Patients were excluded if they met any of the following criteria, 1. Presence of somatic *TNFAIP3* pathogenic variants rather than germline variants. 2. Pathogenic variants in other genes potentially associated with autoinflammatory or autoimmune phenotypes. 3. Missense *TNFAIP3* variants, which were all categorized as variants of uncertain significance (VUS) according to ACMG guidelines,^21^ or variants previously reported as pathogenic but with conflicting or insufficient evidence regarding pathogenicity per ACMG guidelines. Most missense variants identified in this study demonstrated normal or inconclusive results on functional assays. To ensure a high level of diagnostic certainty and minimize misclassification or analytical bias, all missense variants in this study were excluded from the primary analyses.

### Data collection

As a retrospective cross-sectional study, all clinical features were recorded based on cumulative patient histories at the time of enrollment. Data included both pre-treatment and post-treatment observations obtained through patient interviews and review of medical records, thereby reflecting the overall disease course rather than a single time point.

Laboratory data and medication use were collected at four time points, prior to genetic diagnosis, at initiation of treatment following diagnosis, at maximal response (defined as lowest disease activity during longitudinal follow-up), and at the most recent follow-up. Laboratory assessments included peripheral leukocyte count, hemoglobin, platelet count, erythrocyte sedimentation rate (ESR), C-reactive protein (CRP), serum amyloid A (SAA), serum ferritin, autoantibodies, Coombs’ test, and serum levels of IgG, IgE, C3, and C4.

Disease phenotypes and diagnoses were directly assigned by physicians who assessed the patients at each referral center, including BD, IBD, JIA, SLE, PFAPA, possible immunodeficiency, autoimmune hepatitis, and others. Each phenotype was carefully reviewed by two independent authors to confirm diagnostic criteria were met. BD was defined by 2012 International Criteria.^22^ IBD was established by the combination of chronic gastrointestinal symptoms, with or without laboratory abnormalities, coupled with characteristic findings on imaging and endoscopy or colonoscopy, JIA was diagnosed following the Classification Criteria for JIA by EULAR in 2001^23^ or Pediatric Rheumatology International Trials Organization International Consensus (PRINTO) in 2018.^24^ SLE was defined by ACR1997, SLICC or the new EULAR/ACR classification criteria in 2019.^25–27^ Diagnostic criteria for PFAPA included onset of disease in early childhood, regularly recurring episodes of fever, presence of aphthous stomatitis, pharyngitis, and/or cervical adenitis during flares, asymptomatic intervals between flares with normal growth, and absence of signs of respiratory tract infection during flares.^28^

Minimal disease activity was defined as the absence of clinical symptoms or signs (by patient or physician report), global assessment scales for physicians (PGA) ≤ 2/10, and normal inflammatory markers (CRP ≤ 0.5 mg/L, ESR ≤20 mm/h, and/or SAA ≤ 10 mg/L). Partial response was defined as clinical improvement but failure to achieve minimal disease activity (i.e. symptoms or signs improved but still present by patient/physician report, PGA > 2/10, decreased but not completely normalized levels of inflammatory markers). Nonresponse was defined as absence of improvement or disease progression in clinical symptoms or signs, PGA, and/or inflammatory markers. Disease flare was defined as relapsed or worsening clinical symptoms or signs accompanied by PGA > 2/10, CRP level of > 30 mg/L and/or ESR > 30 mm/h. Maximal response for each patient was defined as the lowest disease activity (based on clinical symptoms and signs, PGA, CRP, and ESR) achieved during the period of longitudinal follow-up, even if higher than minimal disease activity.

Infections were defined as acute symptoms or signs accompanied by elevated inflammatory markers and positive pathogen detection, which was resolved by antibiotic or antiviral therapy rather than GC or immunosuppressants.

### Genetic testing and DNA sequencing

Genomic DNA was extracted from peripheral blood from patients and parents. For Chinese patients, the exonic regions and flanking splicing or intronic junctions of the whole genome were captured and sequenced using an Illumina HiSeq 2000 sequencer conducted by Chigene (Beijing, China). The FASTQ files were mapped to the human reference genome (hg19) and variants were called by GATK best practice. Within the U.S. cohort, mutations were identified using commercial multigene panel assays and/or clinical WES analysis.

### NF-κB reporter assay

NF-κB reporter assay was performed as previously described.^1^ Preformulated Cignal NF-κB Pathway Reporter plasmids (Qiagen) (pGL4.32 [luc2P/NF-κBRE/Hygro] luciferase reporter plus pRLCMV-Renilla vector) were used to co-transfect in HEK293T cells with equal amount of expression plasmids for GFP-tag *TNFAIP3* wild type or mutants using Polyjet transfection reagents (Promega, Cat.No.E2691). After 48 hours, cells were left untreated or stimulated with TNF (20 ng/mL). Luciferase activity (the firefly (luc) and renilla luciferase (ren) activity) was measured 5 hours afterwards by the dual-glo luciferase assay system (Promega, Cat.No. E2940). Results for NF-κB activity are expressed as fold induction by normalizing firefly luciferase activity to Renilla luciferase activity and further normalizing to the mean value of the wild-type control.

### Statistical analysis

All analyses were performed based on patients for whom the relevant data were available. Missing data points were excluded from both the numerator and denominator. When evaluating clinical response, all patients with treatment information were included in the denominator, including those lacking clinical response data, to avoid artificially inflating the treatment response rate.

To explore the potential drivers of clinical heterogeneity of HA20, patients were grouped by clinical features through hierarchical clustering based on the Hamming distance metric and complete linkage (Supplementary Figure 1). Based on this analysis, the major features associated with heterogeneity were defined as age and country of origin. Further *s*ubgroup analyses were performed based on age, country of origin (China vs U.S.), and gender. Pediatric and adult subgroups were defined according to age at genetic diagnosis, with pediatric patients classified as those diagnosed before 16 years of age and adult patients as those diagnosed at or after 16 years, consistent with the age-based classification used for certain rheumatic diseases proposed by EULAR.^23, 29^ This stratification was designed to investigate potential differences in disease presentation related to developmental factors, to capture potential progression of disease features over time. Analyses stratified by country of origin were performed to explore potential geographical, environment and healthcare-related differences in disease presentation and management. Given the known impact of sex and gender on immune-mediated diseases, analyses stratified by gender were also performed to determine whether gender-related differences contribute to the observed heterogeneity in HA20 phenotypes.

Data analysis and visualization was performed with GraphPad Prism 9.0 statistical software (GraphPad Software Inc., La Jolla, CA, USA), R Studio, and Morpheus (Broad Institute). Continuous variables were presented by the median and range, and categorical variables were presented as percentages and frequencies. Normal distribution of data was tested by Kolmogorov-Smirnov test. As no data followed a normal distribution, Mann-Whitney U test was performed. Clinical disease phenotypes were grouped according to their co-occurrence patterns across individuals through hierarchical clustering based on the Pearson distance metric and complete linkage. Correlation analysis between laboratory abnormalities and clinical features or disease phenotypes was based on Fisher’s exact test, while the direction (positive or negative) of association was determined using Pearson correlation. *P* < 0.05 was assumed to be significantly different.

## RESULTS

### General characteristics and clinical features

A total of 185 patients with HA20 from 112 unrelated families were included in this study, comprising 116 patients from the Chinese cohort and 69 from the U.S. cohort. The median age at disease onset was 3.3 years (range, 0.04 to 33 years) (Table 1, Figure 1A). The median age at diagnosis of all patients was 10.2 years (range, 0.17 to 72 years) (Table 1, Figure 1A). Among all patients, 109 (63.3%) were children, and 78 (44.8%) were male (Table 1). The median follow-up duration was 15 months (range, 0 to 241 months) (Table 1). The most common clinical features in HA20 patients included mucocutaneous involvement, recurrent fever, gastrointestinal symptoms, cytopenias, arthritis/arthralgia, and recurrent infection (Table 1 and Figure 1B).

**Fig 1.**
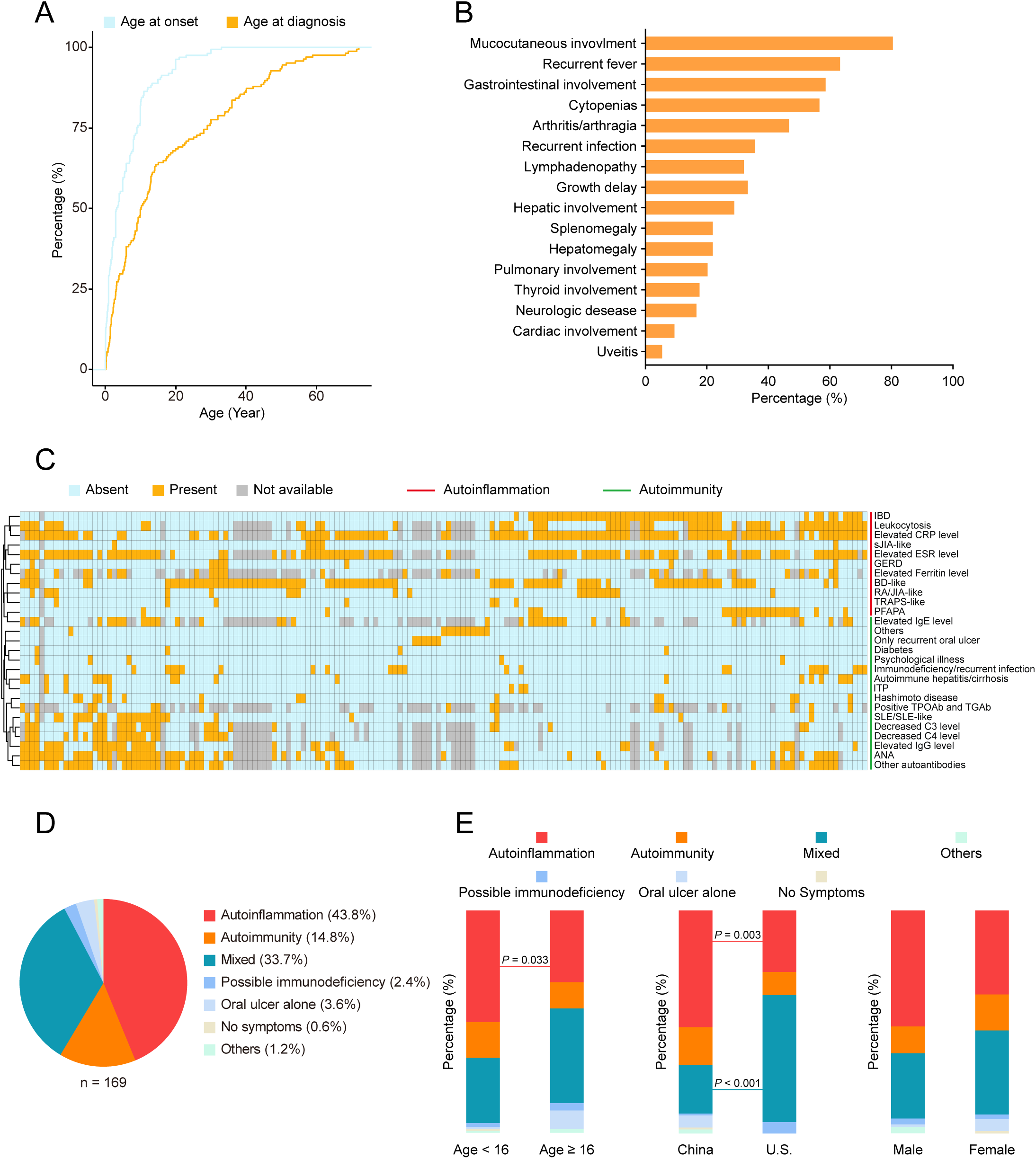
Clinical phenotype of HA20 patients. **A**, Cumulative distribution function plot shows proportion of patients presenting at various disease onset ages and diagnosed ages, respectively. **B**, Bar graph shows percentage of patients presenting with various clinical manifestations. **C**, Pearson-based heatmap shows variation and clustering of clinical disease phenotypes and laboratory abnormalities in HA20. Each column represents an HA20 patient. IBD, inflammatory bowel disease; GERD, gastroesophageal reflux disease; BD, Behçet’s disease; RA, rheumatoid arthritis; sJIA, systemic juvenile idiopathic arthritis; TRAPS, tumor necrosis factor receptor–associated periodic syndrome; PFAPA, periodic fever with aphthous pharyngitis and adenitis; ITP, immune thrombocytopenia; SLE, systemic lupus erythematosus; ESR, erythrocyte sedimentation rate; CRP, C-reactive protein; ANA, antinuclear antibodies; TPOAb, anti-thyroid peroxidase antibody; TGAb, anti-thyroglobulin antibody. **D**, Pie chart shows percentage of patients presenting with autoinflammation-predominant disease, autoimmunity-predominant disease, mixed autoinflammation-autoimmunity disease, or other predominant disease phenotypes. **E**, Stacked bar chart with 100% total shows percentage of patients presenting with autoinflammation-predominant, autoimmunity-predominant, or mixed autoinflammation-autoimmunity disease across different subgroups stratified by age (Left), country of origin (Middle), and gender (Right). Statistical analysis was performed using Mann-Whitney U test. *P* < 0.05 was assumed to be significantly different.

**Table 1.**
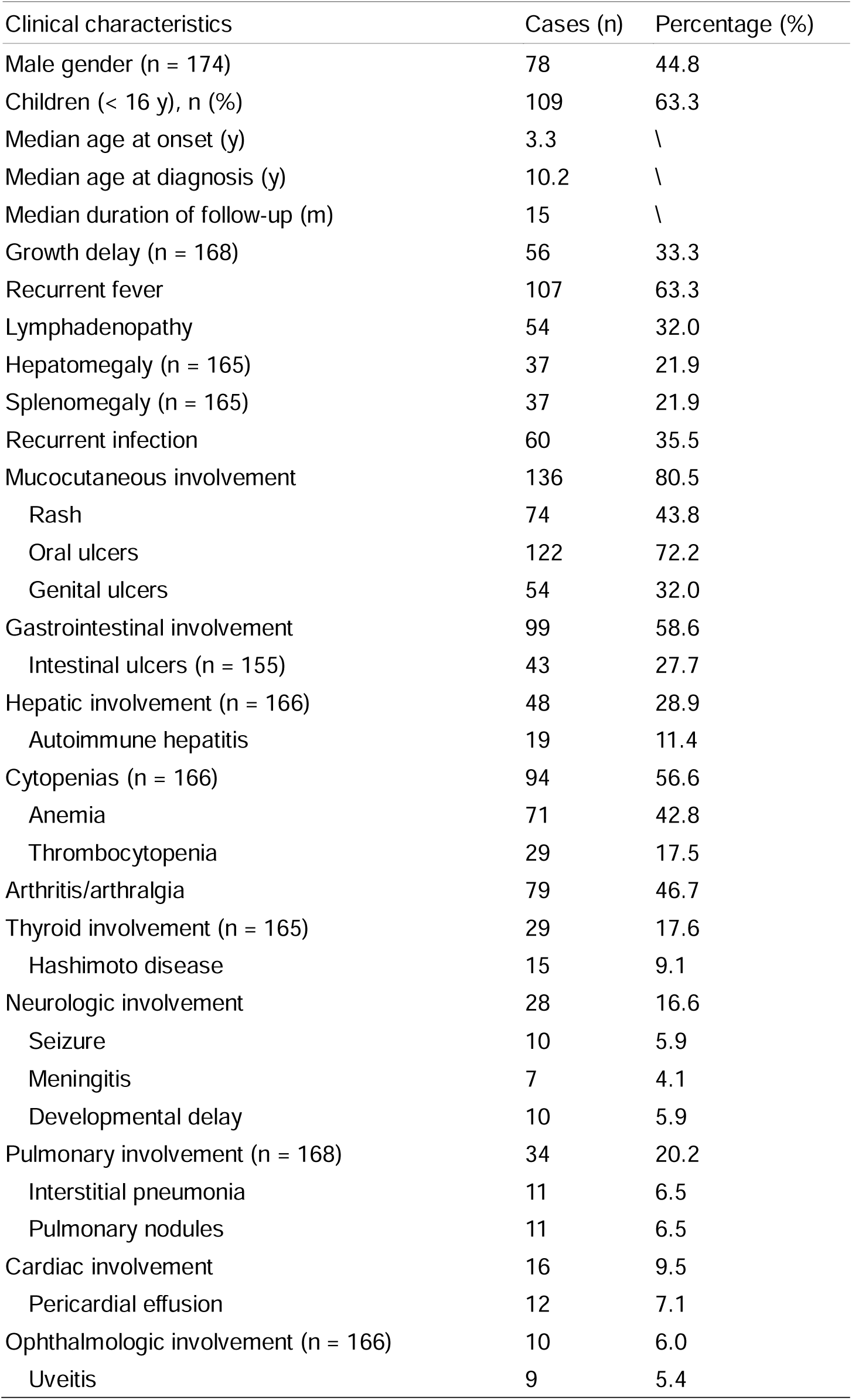
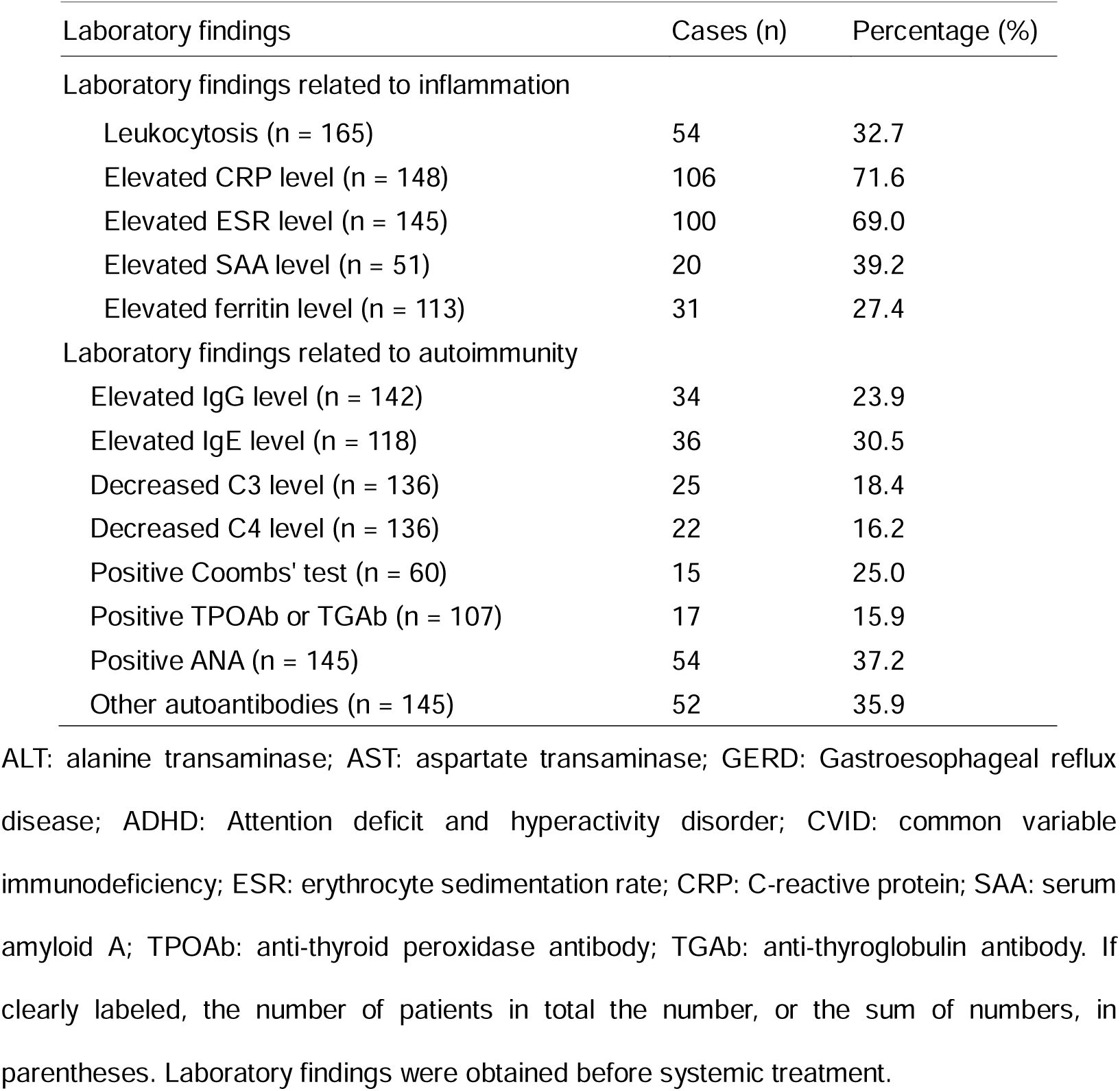
Clinical features and laboratory findings of HA20.

Constitutional symptoms – 107 patients (63.3%) had recurrent fever. 56 patients (33.3%) presented with growth delay. Lymphadenopathy, hepatomegaly, and splenomegaly were seen in 54 patients (32.0%), 37 patients (21.9%) and 37 patients (21.9%), respectively (Table 1 and Figure 1B). 60 patients (35.5%) were ever diagnosed as recurrent infection, which could have been related to the underlying disease, immunomodulatory medication use, or both. Common infections included bronchitis, pneumonia, and gastrointestinal infections. 11 patients (6.5%) had urinary tract infections, two had suppurative lymphnoditis, and six presented with Salmonella infection (Supplementary Table 1 and 2).

Mucocutaneous involvement – 136 patients (80.5%) exhibited mucocutaneous manifestations. Recurrent oral ulcers, skin rash, and genital ulcers were observed in 122 patients (72.2%), 74 patients (43.8%), and 54 patients (32.0%), respectively (Supplementary Table 1). HA20-associated skin involvement was heterogeneous and included urticariform rash, psoriasiform rash, erythema nodosum, pustular eruption, vasculitis-like lesions, purpura, epifolliculitis, alopecia, vitiligo-like rash, and chilblain-like rash. Less common mucocutaneous manifestations included perianal ulcers and migratory glossitis.

Gastrointestinal and hepatic involvement – 99 patients (58.6%) presented with gastrointestinal manifestations, including 62 (37.6%) reporting abdominal pain and 64 (37.9%) experiencing diarrhea. Ten patients (6.0%) had intestinal hemorrhage. Gastrointestinal endoscopy revealed ulcerative disease in over one-third of the patients, including 43 (27.7%) of 155 patients with intestinal ulcers, ten (6.4%) of 157 with gastric ulcers, and six (3.8%) with esophageal ulcers (Supplementary Table 1, Figure 2A). Six patients (3.6%) developed intestinal perforation (Supplementary Table 1); biopsies of selected cases revealed variable patterns of inflammation (Figure 2B). Elevated transaminases were present in 47 patients (28.3%), including 19 (11.4%) with autoimmune hepatitis. Seven (4.2%) patients reported jaundice, and four (2.4%) developed cirrhosis (Supplementary Table 1). Liver biopsies were consistent with autoimmune hepatitis in some cases but revealed nonspecific inflammation in others (Figure 2C).

**Fig 2.**
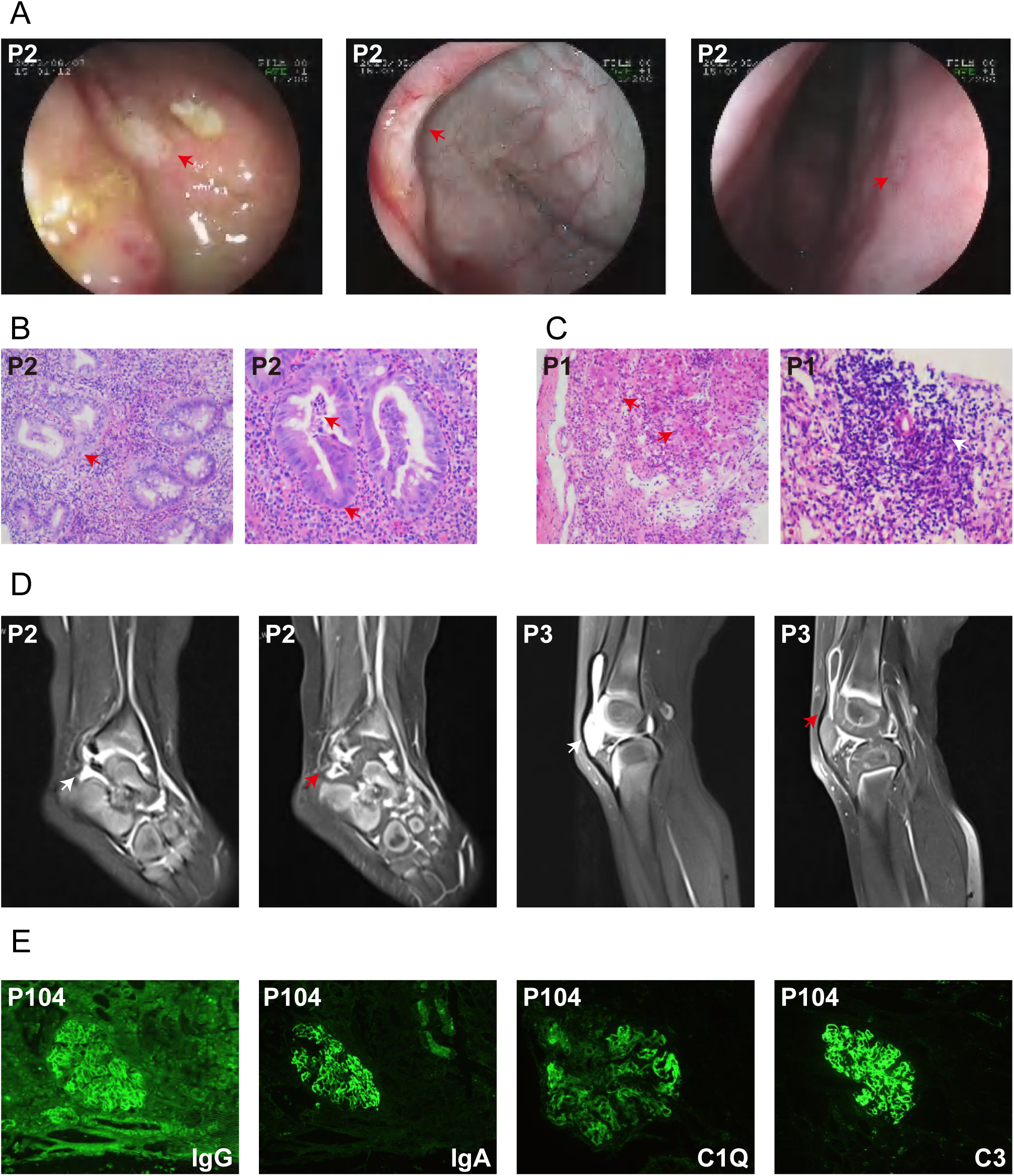
Imaging, colonoscopy, and pathological findings in HA20 patients with major clinical phenotypic subgroups. **A-B**, Representative images of autoinflammation-predominant disease, diagnosed as inflammatory bowel disease prior to genetic diagnosis: colonoscopy images (P2) of terminal ileum, ileocecal valve, and descending colon; Red arrow, ulcers (A); Intestinal biopsy, H&E staining (P2), Red arrow indicates neutrophil infiltration (B). **C-E**, Representative images of autoimmunity-predominant disease features diagnosed as autoimmune hepatitis (C), polyarticular or oligoarticular juvenile idiopathic arthritis (JIA) (D), and lupus nephritis (E) prior to genetic diagnosis. Hepatic biopsy, H&E staining (P1) (C). Red arrow indicates piecemeal necrosis; White arrow indicates infiltration of abundant lymphocytes. Ankle MRI imaging (P2), knee MRI (P3) (D). White arrow indicates joint effusions and synovitis; Red arrow indicates thickened and intensified synovial membrane in patients diagnosed with polyarticular/oligoarticular JIA. Immunofluorescent probe for immune complexes in kidney biopsy from a patient diagnosed with systemic lupus erythematosus (P104) (E).

Hematologic abnormalities – 94 patients (56.6%) had cytopenias, including anemia and lymphopenia in 71 (42.8%) and 45 (27.1%) patients, respectively. One patient developed microangiopathic hemolytic anemia. Thrombocytopenia was seen in 29 patients (17.5%). Neutropenia was found in 30 patients (18.1%) (Supplementary Table 1). Three adult patients (1.8%) developed B cell lymphoma (Supplementary Table 1).

Musculoskeletal involvement – 79 patients (46.7%) experienced arthritis and/or arthralgia, including four (2.4%) with concurrent myositis (Figures 1B and 2D, Supplementary Table 1). Twelve patients (7.1%) reported myalgia without myositis (Supplementary Table 1).

Thyroid involvement – 29 patients (17.6%) had autoimmune thyroiditis, including Graves’ disease, Hashimoto’s disease, and other types of thyroiditis. 19 (11.5%) experienced hypothyroidism, and three (1.8%) had temporary hyperthyroidism (Supplementary Table 1).

Neurologic involvement – 28 patients (16.6%) presented with neurologic symptoms, including seizure (5.9%), meningitis (4.1%), developmental delay (5.9%), cerebral vasculitis (2.4%), stroke (2.4%), sensorineural hearing loss (1.8%), and peripheral neuropathy (1.2%). Three patients developed recurrent Bell’s palsy (Supplementary Table 1).

Pulmonary involvement – 34 patients (20.2%) had pulmonary involvement, including pleural effusion (7.1%), interstitial pneumonia (6.5%), pulmonary nodules (6.5%), asthma (2.4%), cysts (0.6%), consolidation (1.8%), and pneumothorax (0.6%) (Supplementary Table 1).

Cardiac involvement – 16 patients (9.5%) had cardiac manifestations, including pericardial effusion (7.1%), decreased ejection fraction (3.0%), coronary vasculitis or coronary artery dilation (3.0%), and non-compaction cardiomyopathy (0.6%) (Supplementary Table 1).

Ocular manifestations – 10 patients (6.0%) had ocular involvement, presenting with uveitis (5.4%), xerophthalmia, retinal vasculitis, macular degeneration, cataracts, corneal ulcers, or vitreous hemorrhage (Supplementary Table 1).

Other – Other disease manifestations included lupus nephritis (Figure 2E), autoimmune diabetes, fibromyalgia, and premature ovarian failure (Supplementary table 1).

### Subgroup analysis of clinical features

Hamming-based clustering of patients identified two major groups, Group 1 (n = 110; median age, 13 years) and Group 2 (n = 54; median age, 5.95 years) (*P* = 0.00027, Wilcoxon rank-sum test) (Supplementary Figure 1). Within Group 1, two additional subgroups were delineated, Group 1A characterized by autoimmune-predominant and mixed clinical presentations, and Group 1B, characterized by autoinflammation-predominant and mixed clinical presentations.

We further examined the distribution of clinical manifestations across subgroups stratified by age (pediatric vs. adult), country of origin (China vs U.S.), and gender using Mann–Whitney U test. Growth delay, hepatomegaly and intestinal ulceration were significantly more frequent in pediatric patients than in adults (4.7-fold, 2.2-fold and 3.1-fold, respectively; *P* = 0.0001, 0.03 and *P* = 0.002, respectively) (Supplementary Table 3). In contrast, oral ulcers, genital ulcers, and uveitis were less common in children (1.3-fold, 2.5-fold, and 14.9-fold, respectively; *P* = 0.004, 0.002 and 0.001, respectively) (Supplementary Table 3).

There was no significant difference in age at disease onset between cohorts (*P* = 0.09) (Supplementary Table 1). However, the Chinese cohort had a significantly younger age at diagnosis and a higher proportion of pediatric patients (*P* < 0.0001 and = 0.0005, respectively). Compared with the U.S. cohort, growth delay (1.7-fold) and intestinal ulceration (5.8-fold) were more frequent in the Chinese cohort (*P* = 0.0385 and *P* < 0.0001, respectively), whereas genital ulcers, elevated ALT/AST, cytopenias, arthritis/arthralgia and uveitis were less prevalent (*P <* 0.0001) (Supplementary Table 1).

We further performed a subgroup analysis in pediatric patients between the two cohorts, as most pediatric cases underwent comprehensive rheumatologic or immunologic evaluations. The findings were largely consistent with those obtained when adult patients were included (Supplementary Table 1 and 4).

Apart from a 2.2-fold higher prevalence of genital ulcers among female patients, no other clinical features demonstrated significant gender-based differences (Supplementary Table 5).

### Clinical disease phenotypes

Hierarchical clustering of clinical disease phenotypes based on Pearson distance identified two major clusters, an autoinflammation-predominant phenotype and an autoimmune-predominant phenotype (Figure 1C). We therefore evaluated the predominant presentation of each HA20 patient in this study. Among 169 patients with detailed clinical records, 74 patients (43.8%) presented with autoinflammation-predominant features, defined as mainly autoinflammatory diagnoses without evidence of autoimmunity. Only 25 individuals (14.8%) manifested with autoimmunity-predominant disease, defined as autoimmune diagnoses without a major autoinflammatory component. However, 57 individuals (33.7%) had a mixed autoinflammation-autoimmunity phenotype. One adult patient presented with squamous cell carcinoma, one adult patients with SLE and vasculitis developed acoustic neuroma, and another three adult patients had B-cell lymphoma; an infant (younger than the median onset age) found by a family segregation analysis denied any symptoms, whose mother was the proband (Figure 1D and Supplementary Table 2). The most common autoinflammation-predominant phenotypes included BD, IBD, PFAPA, and sJIA or TRAPS-like. The most common autoimmunity-predominant disease phenotypes included SLE, polyJIA/oligoJIA/RA, autoimmune hepatitis or cirrhosis, and Hashimoto disease (Figure 1C).

We further compared the distribution of predominant disease categories across different subgroups. Autoinflammation-predominant disease was 1.6-fold more frequent in pediatric patients (< 16 years) than in adults (≥ 16 years) (*P* = 0.033) (Figure 1E Left). When stratified by country, 58 patients (52.3%) in the Chinese cohort exhibited autoinflammation-predominant disease, significantly higher than in the U.S. cohort (27.6%) (*P* = 0.003) (Figure 1E Middle). In contrast, mixed autoinflammation–autoimmunity was less frequent in the Chinese cohort (21.6%) compared with the U.S. cohort (56.9%) (*P* < 0.001) (Figure 1E Middle). No significant gender-based differences in the distribution of predominant disease categories were observed (Figure 1E Right).

We further conducted multiple linear regression analysis based on Least suqares with predominant phenotype as the outcome and age and country as variables. Consistent with the stratified analyses, both age and country contributed to the likelihood of an autoinflammation-predominant phenotype (*P* = 0.0169 and 0.0067, respectively) (Supplementary Table 6).

### Laboratory findings

Laboratory tests obtained during periods of active disease demonstrated leukocytosis (32.7%), elevated CRP (71.6%), ESR (69.0%) and SAA (39.2%), hyperglobulinemia (23.9%), elevated serum IgE level (30.5%), decreased levels of C3 (18.4%) and C4 (16.2%), positive Coombs’ test (25.0%), and positive TPOAb and TGAb (15.9%), and ANA positivity (37.2%). Other autoantibodies (35.9%) included anti-dsDNA, anti-SSA, anti-PAIgG, anti-PAIgM, anti-cardiolipin, anti-ribosomal P protein, anti-Scl, and anti-U1 RNP (Supplementary Table 2). Leukocytosis and elevated IgE levels were more common in children than in adults (1.9-fold and 2.6 fold, respectively; *P* = 0.015 and 0.03, respectively) (Supplementary Table 3). In contrast, reduced C4 levels (2.5-fold; *P* = 0.02), positive anti-thyroid antibodies (2.8-fold; *P* = 0.026), ANA positivity (1.8-fold; *P* = 0.023), and other autoantibodies (2.3-fold; *P* = 0.0005) were all less common in children than in adults (Supplementary Table 3).

We further performed association analysis between laboratory abnormalities and clinical manifestations or disease phenotypes using Fisher’s exact test and Pearson correlation. TPOAb and TGAb positivity showed a strong association with thyroid disease. Reduced C3 or C4 levels were significantly associated with rash and splenomegaly. ANA and other autoantibodies were negatively associated with intestinal ulceration (Supplementary Figure 2A). ANA positivity, other autoantibodies, and reduced C3 or C4 were enriched in patients with SLE/SLE-like disease but negatively associated with IBD. Elevated serum IgG was positively associated with SLE/SLE-like disease but negatively associated with IBD or BD-like disease (Supplementary Figure 2B).

### Genetic analysis

From 185 patients, a total of 89 *TNFAIP3* pathogenic variants were identified, including 24 nonsense mutations (68 patients), 31 small deletions of one or more bases (56 patients), 10 larger deletions of exons or segments of chromosome 6 including *TNFAIP3* (15 patients), 5 splicing site mutations (9 patients), 12 duplications of one or more bases (30 patients), 3 small insertions (3 patients), and 4 indels (4 patients) (Figure 3A, 3B and Supplementary Table 7). There were 43 known variants and 46 novel variants. The most common mutations were R271*, R183*, deletion of *TNFAIP3*, R45*, K643*, S79Afs*17, each found in at least 6 patients (Figure 3A). All mutations were predicted to result in the expression of a truncating protein or a heterozygous deletion, leading to haploinsufficiency of A20. All novel coding mutations were functionally validated using a reporter assay that assessed their ability to suppress NF-κB activation downstream of TNF stimulation (Supplementary Figure 4).

**Fig 3.**
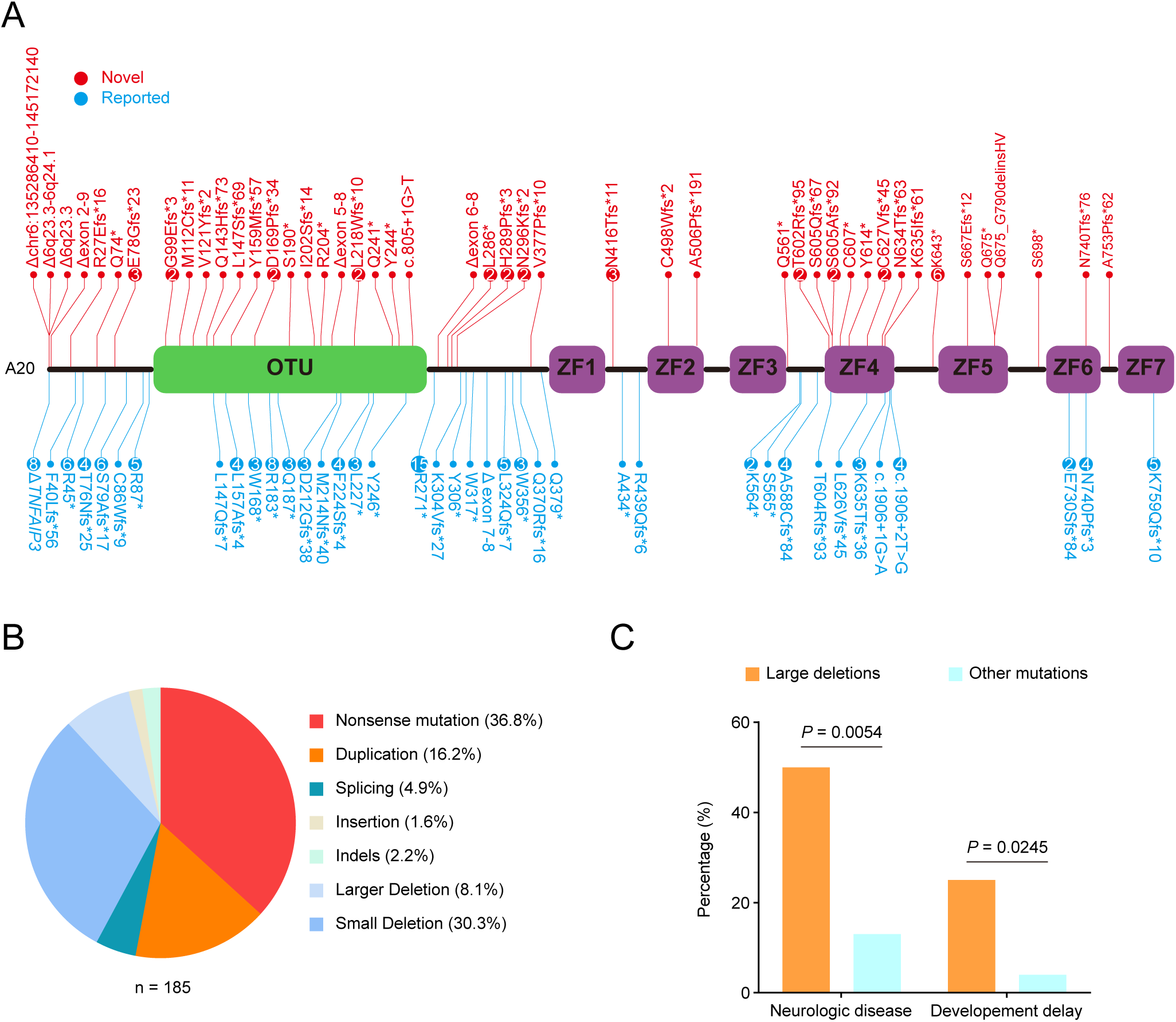
Genetic analysis of HA20 patients. **A**, Schematic representation of *TNFAIP3* gene shows variants identified in HA20 patients. Red font indicates novel variants; Blue font indicates reported variants. Recurrent variants in this cohort are indicated by larger circles with the number of recurrences inside. **B**, Pie chart shows percentage of patients with various mutation types. **C**, Bar chart shows percentage of patients with large deletion mutations or other mutations presenting with neurologic disease (*P* = 0.0031, Mann-Whitney U test) or developmental delay (*P* = 0.038, Mann-Whitney U test).

We further analyzed the clinical phenotypes of patients with mutations localized to various A20 domains. There was no significant difference in age of disease onset or the percentages of patients with various clinical features and disease categories between patients with the OTU domain mutations and those with ZnF domain mutations (all *P* > 0.05) (Supplementary Figure 3A and Supplementary Figure 3B). These clinical phenotypes were caused by mutations dispersed across all exons of the *TNFAIP3* gene and all domains of the A20 protein (Figure 3A). In contrast, large deletions, defined as deletions involving three or more exons, were associated with a higher frequency of neurologic disease (50.0% vs. 13.9%; *P* = 0.0054) and developmental delay (25.0% vs. 4.4%; *P* = 0.0245) compared with other mutation types (Figure 3C).

### Treatment

Compared with treatment before diagnosis, fewer patients received GC alone, while more patients were treated with b/tsDMARDs alone or in combination with GC after diagnosis (*P* = 0.0048, *P* < 0.0001, *P* = 0.01, and *P* = 0.0245, respectively) (Figure 4A).

**Fig 4.**
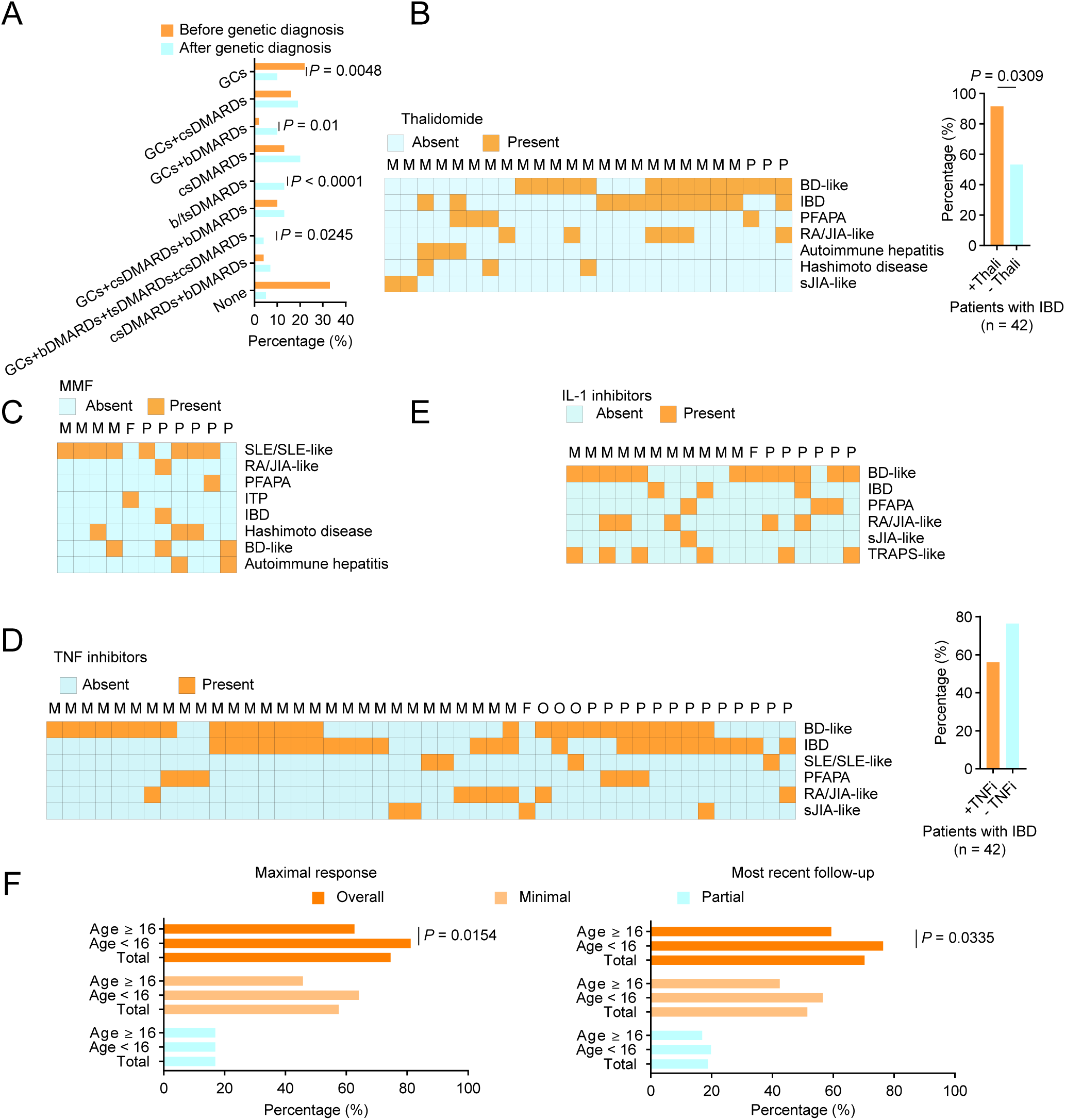
Treatment and clinical response of HA20 patients. **A**, Bar graph shows the distribution of treatment types among patients prior to diagnosis and immediately after the initial diagnosis. Statistical analysis was performed using Mann-Whitney U test. *P* < 0.05 was assumed to be significantly different. GCs, glucocorticoids; csDMARDs, conventional synthetic disease-modifying antirheumatic drugs; bDMARDs, biological DMARDs; tsDMARDs, targeted synthetic DMARDs. **B**, Heatmap shows variation of disease phenotypes and clinical response in HA20 patients receiving thalidomide. Bar graph illustrates the percentage of HA20 patients presenting with IBD who achieved minimal disease activity at the most recent follow-up, stratified by treatment with or without thalidomide. Thali, thalidomide. Statistical analysis was performed by Mann-Whitney U test. *P* < 0.05 was assumed to be significantly different. **C**, Heatmap shows variation of disease phenotypes and clinical response in HA20 patients receiving MMF. **D**, Heatmap shows variation of disease phenotypes and clinical response in HA20 patients receiving IL-1 inhibitors. **E**, Heatmap shows variation of disease phenotypes and clinical response in HA20 patients receiving TNF inhibitors. Bar graph illustrates the percentage of HA20 patients presenting with IBD who achieved minimal disease activity at the most recent follow-up, stratified by treatment with or without TNF inhibitors (*P* = 0.2072, Mann-Whitney U test). TNFi, TNF inhibitors. In Panel B to E, each column represents an HA20 patient. M, minimal disease activity; P, partial response; F, disease flare; O, others, including data not available and no response; BD, Behçet’s disease; IBD, inflammatory bowel disease; PFAPA, periodic fever with aphthous pharyngitis and adenitis; RA, rheumatoid arthritis; sJIA, systemic juvenile idiopathic arthritis; SLE, systemic lupus erythematosus; JIA, juvenile idiopathic arthritis; ITP, immune thrombocytopenia; TRAPS, tumor necrosis factor receptor–associated periodic syndrome. **F** Line graph shows the clinical response to treatment over time by disease status. PGA, global assessment scales for physicians, where 10 represents the highest level of disease activity; P, prednisone; I, infliximab; T, thalidomide; A, azathioprine; IV, intra-venous immunoglobulin; MMF, mycophenolate mofetil; H, hydroxychloroquine; Ad, adalimumab; M, methotrexate; E, Etanercept; Tof, tofacitinib; Me, mesalazine; Bar, baricitinib; CS, corticosteroids; Can, canakinumab; C, colchicine; Apr, apremilast. Note, for P137, treatment initiation was delayed because symptoms of chest pain were initially misattributed to atherosclerosis instead of coronary vasculitis. **G** Bar graph shows the percentage of patients with overall response, minimal disease activity, and partial response during the maximal response period and at the most recent follow-up in the total cohort and by age group. Statistical analysis was performed by Mann-Whitney U test. *P* < 0.05 was assumed to be significantly different.

GC were administered to 74 patients (55.2%) and to 39 patients (29.1%) as initial treatment after genetic diagnosis and at the most recent follow-up, respectively (Table 2 and Supplementary Table 8). Among the 13 patients initially treated with GC alone, 8 (61.5%) later switched to the addition of DMARDs during the disease course and subsequently weaned off GC by the most recent follow-up, including 6 patients who completely discontinued GC. Among the 61 patients who initially received GC in combination with DMARDs, 55 (90.2%) were weaned off GC by the most recent follow-up, including 29 who fully discontinued GC (Supplementary Table 8).

**Table 2.**
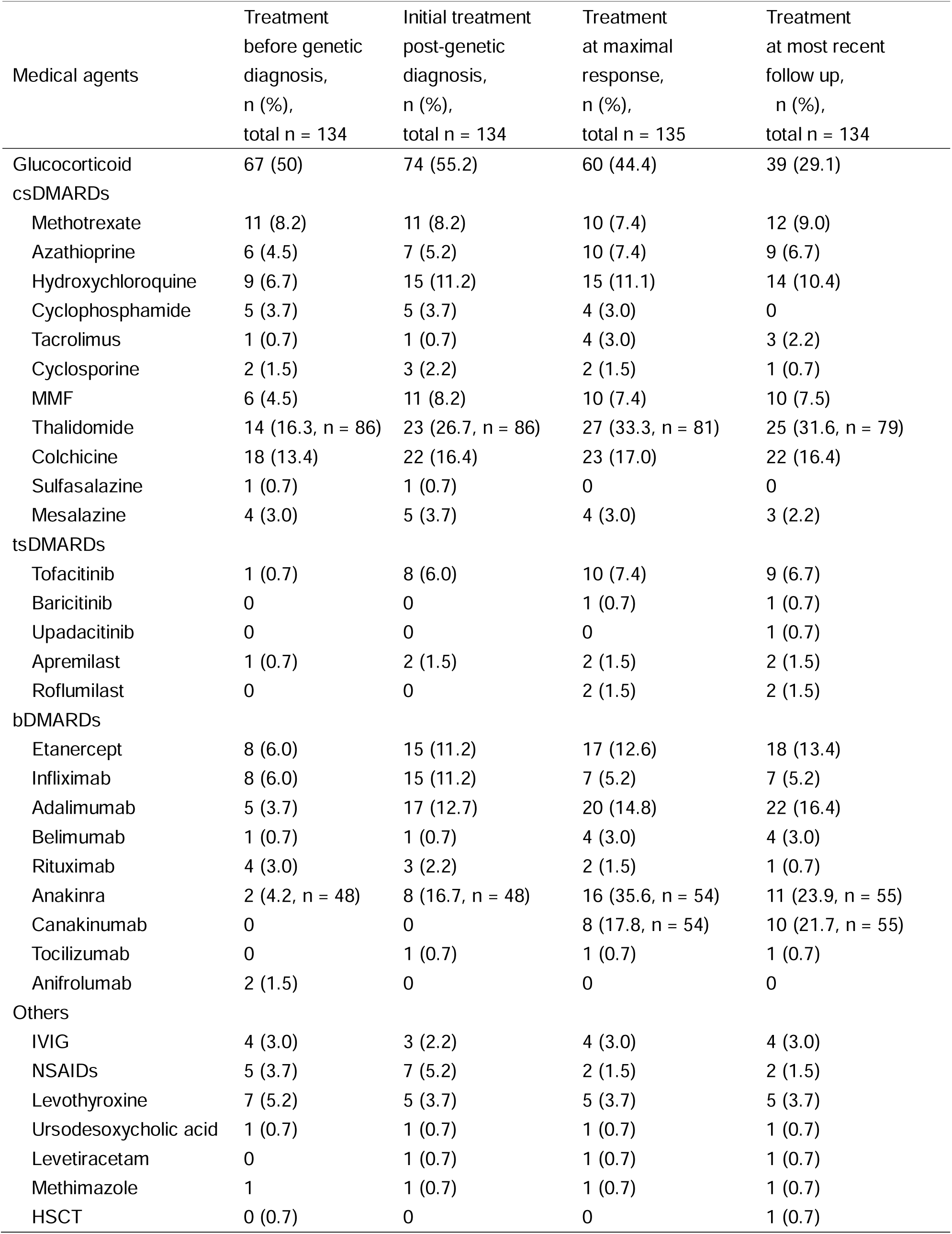
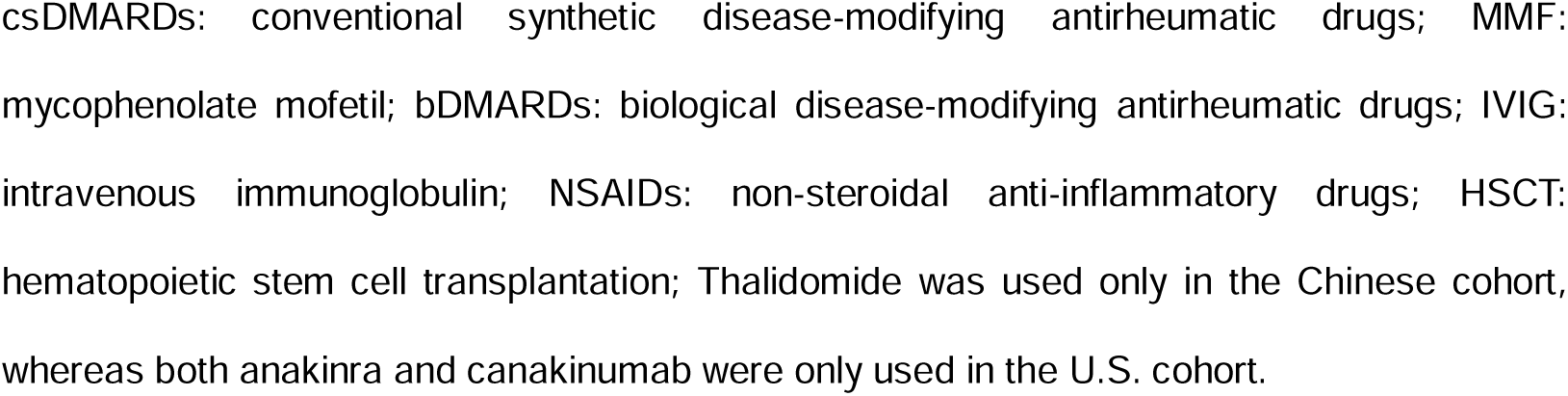
Treatment of HA20 patients.

|  | Treatment<br>before genetic<br>diagnosis,<br>n (%),<br>total n = 134 | Initial treatment<br>post-genetic<br>diagnosis,<br>n (%),<br>total n = 134 | Treatment<br>at maximal<br>response,<br>n (%),<br>total n = 135 | Treatment<br>at most recent<br>follow up,<br>n (%),<br>total n = 134 |
| --- | --- | --- | --- | --- |
| Medical agents |  |  |  |  |
| Glucocorticoid | 67 (50) | 74 (55.2) | 60 (44.4) | 39 (29.1) |
| csDMARDs |  |  |  |  |
| Methotrexate | 11 (8.2) | 11 (8.2) | 10 (7.4) | 12 (9.0) |
| Azathioprine | 6 (4.5) | 7 (5.2) | 10 (7.4) | 9 (6.7) |
| Hydroxychloroquine | 9 (6.7) | 15 (11.2) | 15 (11.1) | 14 (10.4) |
| Cyclophosphamide | 5 (3.7) | 5 (3.7) | 4 (3.0) | 0 |
| Tacrolimus | 1 (0.7) | 1 (0.7) | 4 (3.0) | 3 (2.2) |
| Cyclosporine | 2 (1.5) | 3 (2.2) | 2 (1.5) | 1 (0.7) |
| MMF | 6 (4.5) | 11 (8.2) | 10 (7.4) | 10 (7.5) |
| Thalidomide | 14 (16.3, n = 86) | 23 (26.7, n = 86) | 27 (33.3, n = 81) | 25 (31.6, n = 79) |
| Colchicine | 18 (13.4) | 22 (16.4) | 23 (17.0) | 22 (16.4) |
| Sulfasalazine | 1 (0.7) | 1 (0.7) | 0 | 0 |
| Mesalazine | 4 (3.0) | 5 (3.7) | 4 (3.0) | 3 (2.2) |
| tsDMARDs |  |  |  |  |
| Tofacitinib | 1 (0.7) | 8 (6.0) | 10 (7.4) | 9 (6.7) |
| Baricitinib | 0 | 0 | 1 (0.7) | 1 (0.7) |
| Upadacitinib | 0 | 0 | 0 | 1 (0.7) |
| Apremilast | 1 (0.7) | 2 (1.5) | 2 (1.5) | 2 (1.5) |
| Roflumilast | 0 | 0 | 2 (1.5) | 2 (1.5) |
| bDMARDs |  |  |  |  |
| Etanercept | 8 (6.0) | 15 (11.2) | 17 (12.6) | 18 (13.4) |
| Infliximab | 8 (6.0) | 15 (11.2) | 7 (5.2) | 7 (5.2) |
| Adalimumab | 5 (3.7) | 17 (12.7) | 20 (14.8) | 22 (16.4) |
| Belimumab | 1 (0.7) | 1 (0.7) | 4 (3.0) | 4 (3.0) |
| Rituximab | 4 (3.0) | 3 (2.2) | 2 (1.5) | 1 (0.7) |
| Anakinra | 2 (4.2, n = 48) | 8 (16.7, n = 48) | 16 (35.6, n = 54) | 11 (23.9, n = 55) |
| Canakinumab | 0 | 0 | 8 (17.8, n = 54) | 10 (21.7, n = 55) |
| Tocilizumab | 0 | 1 (0.7) | 1 (0.7) | 1 (0.7) |
| Anifrolumab | 2 (1.5) | 0 | 0 | 0 |
| Others |  |  |  |  |
| IVIG | 4 (3.0) | 3 (2.2) | 4 (3.0) | 4 (3.0) |
| NSAIDs | 5 (3.7) | 7 (5.2) | 2 (1.5) | 2 (1.5) |
| Levothyroxine | 7 (5.2) | 5 (3.7) | 5 (3.7) | 5 (3.7) |
| Ursodesoxycholic acid | 1 (0.7) | 1 (0.7) | 1 (0.7) | 1 (0.7) |
| Levetiracetam | 0 | 1 (0.7) | 1 (0.7) | 1 (0.7) |
| Methimazole | 1 | 1 (0.7) | 1 (0.7) | 1 (0.7) |
| HSCT | 0 (0.7) | 0 | 0 | 1 (0.7) |

csDMARDs: conventional synthetic disease-modifying antirheumatic drugs; MMF:
mycophenolate mofetil; bDMARDs: biological disease-modifying antirheumatic drugs; IVIG:
intravenous immunoglobulin; NSAIDs: non-steroidal anti-inflammatory drugs; HSCT:
hematopoietic stem cell transplantation; Thalidomide was used only in the Chinese cohort,
whereas both anakinra and canakinumab were only used in the U.S. cohort.

Thalidomide was primarily used in patients with disease phenotypes of IBD, BD, arthritis, or PFAPA in the Chinese cohort. The numbers of patients receiving thalidomide at initial treatment, at maximal response, and at the most recent follow-up were 23 (26.7%), 27 (33.3%), and 25 (31.6%), respectively (Table 2). At the most recent follow-up, all 25 patients achieved either minimal disease activity (88.0%) or partial (12.0%) response, including 10 patients treated with thalidomide alone (Figure 4B Left). Among patients with the IBD phenotype, those treated with thalidomide were more likely to achieve a minimal disease activity compared to those not receiving thalidomide (*P* = 0.0309) (Figure 4B Right). MMF, CTX, tacrolimus, and cyclosporine were mainly administered in patients with autoimmune disease, especially those with SLE-like syndromes (Figure 4C and Supplementary Table 8).

TNF inhibitors were primarily administered to patients with BD, IBD, arthritis, or PFAPA (Figure 4D Left). At the most recent follow-up, among the 46 patients treated with TNF inhibitors, 63.0% achieved a minimal disease activity and 28.3% achieved a partial response (Figure 4D Left). The proportion of IBD patients achieving a minimal disease activity did not differ significantly between those treated with TNF inhibitors and those not receiving these agents (*P* = 0.207) (Figure 4D Right). Anti-TNFi antibodies were detected at least in four patients (Supplementary Table 9). Only one patient treated with TNF inhibitors for a presumed diagnosis of polyJIA developed lupus-like features that persisted after discontinuation of TNF inhibitor therapy.

Only patients in the U.S. cohort received IL-1 inhibitors, as these agents were not available for Chinese patients during the study period. IL-1 inhibitors were mainly used in patients with the disease phenotype of BD, IBD, arthritis, or TRAPS-like disease. At the most recent follow-up, among 18 patients, 61.1% had a minimal disease activity and 33.3% a partial response (Figure 4E).

One patient in the U.S. cohort underwent hematopoietic stem cell transplant successfully after losing responsiveness to multiple agents, including IL-1 and JAK inhibitors (Supplementary Table 8).

We further analyzed data from 22 representative patients with detailed longitudinal data, focusing on the timing of maximal response, disease flares, and disease flare triggers. Among these patients, the median follow-up duration was 39.5 months (range, 6–104 months), and the median longest duration of minimal disease activity was 14.7 months (range, 4–104 months).

None of the patients discontinued therapy during sustained periods of remission. Under the treatment guided by genetic diagnosis, a total of 24 disease flares were documented, corresponding to a mean rate of 0.016 flares per patient-year. Common triggers included infection, GC or tofacitinib tapering, and the development of anti-TNFi autoantibodies (Supplementary Table 9).

Among 165 HA20 patients with available clinical response data, 57.6% achieved minimal disease activity at maximal response, and 51.5% maintained this status at the last follow-up. Children showed a better overall response than adults at both time points (*P* = 0.0154 and 0.0335, respectively) (Figure 4F).

### Adverse events

At the most recent follow-up, all patients were alive except for two who were deceased from severe interstitial lung disease complicated by respiratory failure and pulmonary hypertension complicated by acute gastrointestinal bleeding during disease flare, respectively. During the follow-up, 41 patients (31.8%) suffered from various infections (Table 3 and Supplementary Table 10).

**Table 3.** Adverse events of HA20 patients during follow-up.

| Adverse events | Total, n | Percentage (%) |
| --- | --- | --- |
| Infections<sup>#</sup> | 41 | 31.8 |
| Upper respiratory infection | 24 | 18.6 |
| Bronchitis | 2 | 1.6 |
| Pneumonia | 10 | 7.8 |
| Gastrointestinal infection | 5 | 3.9 |
| Lymphadenitis | 2 | 1.6 |
| Urinary tract infection | 2 | 1.6 |
| Cutaneous cellulitis | 1 | 0.8 |
| Drug eruptions* | 13 | 10.1 |
| Injection reaction * | 7 | 5.4 |
| Subcutaneous edema* | 1 | 0.8 |
| Abdominal pain* | 5 | 3.9 |
| Nerve injury* | 3 | 2.3 |
| Dizziness * | 2 | 1.6 |
| Vomiting* | 5 | 3.9 |
| Acoustic neuroma<sup>#</sup> | 1 | 0.8 |
| Constipation* | 2 | 1.6 |
| Depression and anxiety<sup>#</sup> | 2 | 1.6 |
| Global amnesia and fainting spells<sup>\$</sup> | 1 | 0.8 |
| HLH secondary to EBV infection<sup>\$</sup> | 1 | 0.8 |
| Hemolytic anemia<sup>#</sup> | 1 | 0.8 |
| BK polyomavirus reactivation* | 1 | 0.8 |
| Blindness<sup>\$</sup> | 1 | 0.8 |
| Deceases<sup>\$</sup> | 2 | 1.6 |

A total of 129 patients were included in the follow-up for adverse events. Infections were defined as acute symptoms or signs accompanied by elevated inflammatory markers and positive pathogen detection, which were resolved by antibiotics or antiviral therapy rather than glucocorticoids or immunosuppressants. Adverse events secondary to thalidomide treatment included subcutaneous edema, nerve injury, dizziness, and constipation; Blindness was due to thrombocytopenia and intraocular bleeding. HLH: hemophagocytic lymphohistiocytosis. #: Adverse events that were possibly related to both the disease and the use of immunomodulatory medications. \*: Adverse events that were possibly related to the use of immunomodulatory medications. \$: Adverse events that were possibly related to the disease.

Among patients receiving thalidomide, three showed nerve injury and one had both subcutaneous edema and dizziness, leading to discontinuation of thalidomide; another two experienced constipation. Other rare adverse events possibly associated with HA20 or medications included cutaneous drug eruptions, abdominal pain, vomiting etc(Table 3 and Supplementary Table 10).

## DISCUSSION

We present an updated and comprehensive overview of the natural history of HA20 due to confirmed LOF *TNFAIP3* mutations, using a large international, multicenter cohort study.

Clinical phenotypes were highly variable, ranging from isolated oral aphthous stomatitis to autoinflammatory syndromes and severe autoimmune disease. The most common manifestations included mucocutaneous involvement, recurrent fever, gastrointestinal symptoms, cytopenias, arthritis/arthralgia, and recurrent infections. Ocular involvement was uncommon overall; however, uveitis occurred more frequently in adults, indicating that ocular disease should be regularly monitored even when baseline evaluations are normal. Growth delay was observed in nearly one third of patients, including many without prior glucocorticoid exposure, suggesting that this feature may reflect the disease itself rather than treatment-related adverse effects.

Clinical disease phenotypes of HA20 can be broadly categorized as autoinflammation-predominant, autoimmunity-predominant, or mixed. The most common autoinflammatory phenotypes included BD, IBD, PFAPA, and sJIA/TRAPS-like disease, while the most frequent autoimmunity-predominant phenotypes were SLE, polyJIA/oligoJIA/RA, autoimmune hepatitis or cirrhosis, and Hashimoto disease. Autoinflammatory diagnoses were more frequently observed in children, although this may partly reflect ascertainment bias, as patients with early-onset autoinflammation are more likely to undergo genetic testing. In contrast, autoimmune manifestations and abnormal laboratory findings were common in adults from the U.S. cohort, potentially reflecting disease evolution. For example, one patient initially presented with PFAPA in early childhood but later developed classic SLE with lupus nephritis. These observations suggest that disease phenotypes may vary by age and, in some cases, evolve from autoinflammatory to autoimmune presentations over time.

Mechanistically, A20 is an important regulator of NF-κB pathway and cell death pathway,^30^ and prevents spontaneous NLRP3 inflammasome activation.^31, 32^ In addition, A20 plays a crucial role in adaptive immunity, especially in the development and function of B cells.^33, 34^ Chronic inflammation caused by A20 deficiency drives activation of monocytes and B cells, predisposing to autoimmunity in HA20 patients. Moreover, HA20 is associated with expansion of self-reactive T cell and B cell clones,^35^ while A20 also prevents spontaneous type I interferon induction^36^—features that are implicated in both autoinflammatory and autoimmune diseases. Together, these mechanisms may explain how HA20 licenses dual patterns of immune dysregulation.

Three adult patients in the U.S. cohort had B-cell lymphoma. Somatic variants in A20 are specifically associated with B cell lymphoma,^37, 38^ and HA20 skews B cells towards pre-lymphoma immunophenotypes and transcriptomic signatures.^35^ One patient from NIH presented with GI B-cell lymphoma as the presenting manifestation, although he had reported a longstanding antecedent history of abdominal pain and oral ulcers. Two other patients reported diffuse large B-cell lymphomas, further suggesting that HA20 patients may require close monitoring for this potential complication. We also found instances of squamous cell carcinoma and acoustic neuroma. While A20 also acts as a tumor suppressor in hepatocellular and colorectal cancers,^39^ we did not see these cancers in our cohort, and they have not been described in HA20 patients. Long term follow-up and natural history studies will help to further evaluate the risk of various cancers in HA20 patients.

While large deletion size was associated with neurologic disease and developmental delay, we did not see other genotype-phenotype correlations. Contrary to prior studies, we specifically failed to see differences between variants in ZnF and OTU domains. There are several potential reasons for this, prior studies have largely been meta-analyses of case reports and case series, which could have led to over-representation or under-representation of specific clinical features depending on the level of detail in various case reports.^2^ This could have resulted in “false positive” genotype-phenotype associations. Another important difference is that prior meta-analyses have included patients with missense *TNFAIP3* variants, even those with inconsistent evidence regarding their disease-causing potential.^18^ Prior meta-analyses have not stratified genotype-phenotype associations by both domain and mutation type (missense vs. pLOF).

More broadly, the potential role of missense variants in HA20 remains controversial. We chose to exclude patients with missense variants from this cohort study, as the pathological significance of missense variants cannot be concluded solely based on *in silico* predictions, allele frequencies or patient symptoms. Although several missense variants have been reported to cause HA20, none of these reported variants were detected in our cohort. Moreover, most of these putative disease-causal *TNFAIP3* variants have not been compared to wild-type *TNFAIP3* using functional studies.^4, 18^ When missense variants have undergone testing, methodologies have varied between studies and have included A20 expression, A20 stability, assessment of A20-regulated pathways in primary cells, and assessment of A20-mediated inhibition in ectopic overexpression-based assays. Hence, future studies and standardized methodologies are needed to definitively establish the pathological significance of missense variants prior to including these patients in a HA20 cohort study.

Consistent with prior reports, most of the mutations we identified were fully penetrant. However, we found one asymptomatic patient with a LOF *TNFAIP3* mutation via family segregation analysis. She was further evaluated by kidney disease physicians to assess for subclinical disease. The presence of the asymptomatic infant within our cohort has raised the possibility that clinical disease has not yet developed but may develop in the future. It is also worth noting that minimally symptomatic disease (i.e. oral aphthous ulcers, arthralgia) in some Chinese adults did not lead to a clinical evaluation by a medical professional even after the genetic diagnosis. And several patients in the U.S. experienced diagnostic delays (up to 50 years after onset of symptoms) related to social determinants of health. In our study, which allowed electronic medical record review for qualitative experiences and feedback, some patients reported social stigma related to specific symptoms, especially genital ulcers. Many adult patients experienced disease onset in the pre-genetic testing era, possibly leading them to minimize symptoms until younger family members developed disease.^40^ Finally, many patients in resource-limited settings reported lack of access to providers familiar with genetic and autoinflammatory diseases.^41^ The effects of social determinants of health in the study and treatment of immune dysregulation diseases remains an area of active investigation.

Currently, there is no standard treatment protocol for HA20. MMF was often used in cases with SLE/SLE-like phenotype. Thalidomide could help to control inflammation in HA20 patients, especially those with IBD phenotype, but adverse reactions such as nerve injury should be carefully monitored. JAK inhibitors were reported to be effective in some patients whose interferon-stimulated genes (ISG) expression was elevated during the active disease phase.^36, 42^ In this cohort, JAK inhibitors helped to reduce GCs and alleviate arthritis in some patients, making JAK inhibitors potential effective options in some patients.

IL-1 inhibitors showed therapeutic effects in most patients who received them, ranging from partial response to minimal disease activity. One patient received tocilizumab and showed a good response. Patients presenting with BD, IBD, or arthritis generally responded well to TNF inhibitors. However, anti-drug antibodies should be evaluated in cases of poor response or disease flare. Collectively, IL-6-, IL-1-, and TNF inhibitors may appear to represent effective therapeutic strategies in HA20 patients with systemic autoinflammation. In contrast to other severe inborn errors of immunity (IEIs), treatment outcomes for patients in this cohort were favorable, resembling those observed in monogenic systemic autoinflammatory diseases.^43^ Nonetheless, infection risk should be carefully considered when selecting therapies, particularly for patients with underlying humoral immune deficiencies.

## LIMITATIONS

This study has several limitations. Differences in cohort composition and evaluation may have influenced the results. Most Chinese patients were referred from pediatric centers, whereas the U.S. cohort included a larger proportion of adults. Additionally, a subset of adults in the Chinese cohort was diagnosed through family segregation analysis rather than standardized clinical assessment. This could affect a modest underestimation of autoimmune features and affecting age-related comparisons. Treatment availability also differed substantially between countries; for example, IL-1 inhibitors recommended for autoinflammatory manifestations were not accessible to the Chinese cohort during the study period. These differences likely influenced treatment patterns, therapeutic responses, and overall comparability between cohorts.

The retrospective and cross-sectional nature of the study also imposes inherent constraints. Although treatment patterns were captured across multiple agents and time points, longitudinal data reflecting the full disease trajectory—such as phenotypic evolution, flare frequency, timing of treatment escalation, or achievement of sustained remission—were incomplete.

Variation in timing of clinical evaluations may have resulted in under– or over-representation of specific manifestations depending on disease activity or treatment status at assessment. Finally, genetic analyses excluded missense variants, and splice-site or deletion variants were not functionally validated; pathogenicity was inferred using standard ACMG criteria. These factors might have limited detailed exploration of genotype–phenotype relationships. Future studies incorporating transcript and functional analyses in larger, prospectively followed cohorts will be critical to fully define the pathogenic spectrum and clinical heterogeneity of *TNFAIP3*-related disease.

### Conclusions

HA20 is one of the most common dominantly inherited immune dysregulation disorders, with highly variable phenotypes ranging from isolated oral aphthous stomatitis to systemic autoinflammatory syndromes and severe autoimmune disease. No clear genotype–phenotype correlation explains this heterogeneity. However, disease evolution may follow an age-dependent pattern, shifting from autoinflammation-predominant presentations in childhood to mixed autoinflammation-autoimmunity disease in adulthood. Across centers and countries, this broad clinical spectrum presents major challenges for diagnosis and management, with patients experiencing an average delay of 7 years between symptom onset and diagnosis. Despite these challenges, most patients had favorable short– to medium-term outcomes when treated with targeted immunomodulatory therapies. This large, multicenter, international cohort provides a comprehensive characterization of HA20, highlighting diagnostic and therapeutic strategies to advance precision medicine for this complex disorder.

### Supplementary Material

Refer to Web version on PubMed Central for supplementary material.

## Data Availability

All data produced in the present study and not in the manuscript are available upon reasonable request to the authors

## Acknowledgments

We thank the patients and their family members for their participation and Eric Hanson for generously providing HEK293T cells.

## Contributors

Qing Zhou, Jun Yang and Daniella M. Schwartz directed and supervised the research. Tingyan He wrote the original draft. Tingyan He, Jun Wang, Chen Liu and Manuel Carpio Tumba analyzed the data. Shihao Wang, Youyou Luo, Jie Chen, Guomin Li, Zhou Shu, Song Zhang, Deborah L Stone, Yanyan Huang, Qianying Lv, Wen Xiong, Jinbo Wang, Zhongxun Yu, Elizabeth Kairis, Akuti Kethri, Atif Towheed, Kyr Goyette, Urekha Karri, Tina Romeo, Laia Alsina Manrique De Lara, Daniel L Rosenberg, Daniel Clemente, Juan Carlos Lopez Robledillo, Zanhua Rong, Xue Zhao, Lijun Jiang, Juan Carlos Aldave-Becerra, Ana Beatriz Muñoz-Urribarri, Prasad Thomas Oommen, Priscilla Campbell-Stokes, Meifang Zhu, Peng Liu, Li Guo, Yiping Xu, Zihua Yu, Huajuan Tong, Xiaojian Qiu, Yazhi Zhang, Hongbo Chen, Changming Zhang, Junbin Ou, Congcong Liu, Jinxiang Liu, Yunyan Shen, Jianshe Cao, Xinping Zhang, Kangkang Yang, Ying Bao, Zhijuan Li, Jie Cao, Yuanhui Duan, Fujuan Liu, Buyun Shi, Min Sun, Li Ma, Yongxing Chen, Wei Yang, Xu Han, Shuangyue Ma, Jie Luo, Weiyue Gu, Guoliang Yu, Weina Shi, Lei Sun, Wenhui Li, Yunfei An, Xuemei Tang, Xiaodong Zhao, Tongxin Han, Jin Ma, Yan Li, Yurong Piao, Fei Sun, Dongfeng Zhang, Meina Yin, Shaoling Zheng, Tianwang Li, Huizhong Niu, Lin Li, Shiyue Mei, Fang Zhou, Sirui Yang, Danlu Li, Mei Yan, Huasong Zeng, Ping Zeng, Wenjie Zheng, Xiaozhong Li, Xiaolin Li, Yuling Liu, Lijuan Huang, Haiguo Yu, Zhidan Fan, Min Shen, Yiping Xu, Meiping Lu, Zhihao Fang, Casey Allison Rimland, Hongmei Song, Lance W Peterson, Maryam Ali Yousuf Al-Nesf, Sami Aqel, Dalal Sideeg Mudawi, Nikita S Raje, Voytek Slowik, Julia Harris, Brenda Snyder, Megan A Cooper, Yu Lung Lau, Kader Cetin Gedik, Wenjie Wang, Xiaochuan Wang, Amanda K Ombrello, Ying Huang, Hai-Lin Wu, Li Sun, Huawei Mao, Xiaomin Yu, Zhihong Liu, Ivona Aksentijevich, Daniel L Kastner, Qing Zhou, Jun Yang and Daniella M. Schwartz provided clinical information. Wen Xiong and Charlotte Vera Cuff performed NF-κB reporter assay. Jiebiao Wang performed and supervised statistical analyses. Tingyan He, Jun Wang, Qing Zhou, Jun Yang and Daniella M. Schwartz revised the manuscript, with input from others.

## PATIENT AND PUBLIC INVOLVEMENT

Patients and/or the public were not involved in the design, conduct, reporting or dissemination plans of this research.

## PATIENT CONSENT FOR PUBLICATION

Compliance with Ethical Standards All participating members were enrolled with the approval of the ethics committee of Shenzhen Children’s Hospital or the local ethics committees. A waiver of written informed consent from the Chinese patients was granted as the research involved minimal risk and used anonymized data. For patients from the U.S. study, written informed consent was obtained from the parent, patients, or both (if more than 18 years of age).

## REFERENCE

1. Zhou Q, Wang H, Schwartz DM, Stoffels M, Park YH, Zhang Y, et al. Loss-of-function mutations in TNFAIP3 leading to A20 haploinsufficiency cause an early-onset autoinflammatory disease. Nature Genetics 2016; 48:67–73.

2. Chen Y, Ye Z, Chen L, Qin T, Seidler U, Tian Da, et al. Association of Clinical Phenotypes in Haploinsufficiency A20 (HA20) With Disrupted Domains of A20. Frontiers in Immunology 2020; 11.

3. Yu M-P, Xu X-S, Zhou Q, Deuitch N, Lu M-P. Haploinsufficiency of A20 (HA20): updates on the genetics, phenotype, pathogenesis and treatment. World Journal of Pediatrics 2020; 16:575–84.

4. Karri U, Harasimowicz M, Carpio Tumba M, Schwartz DM. The Complexity of Being A20: From Biological Functions to Genetic Associations. Journal of Clinical Immunology 2024; 44:76.

5. Li G-m, Liu H-m, Guan W-z, Xu H, Wu B-b, Sun L. Expanding the spectrum of A20 haploinsufficiency in two Chinese families: cases report. BMC Medical Genetics 2019; 20:124.

6. Jo KJ, Park SE, Cheon CK, Oh SH, Kim SH. Haploinsufficiency A20 misdiagnosed as PFAPA (periodic fever, aphthous stomatitis, pharyngitis, and cervical adenitis) syndrome with Kikuchi disease. Clin Exp Pediatr 2023; 66:82-4.

7. Lawless D, Pathak S, Scambler TE, Ouboussad L, Anwar R, Savic S. A Case of Adult-Onset Still’s Disease Caused by a Novel Splicing Mutation in TNFAIP3 Successfully Treated With Tocilizumab. Frontiers in Immunology 2018; 9.

8. Deshayes S, Bazille C, El Khouri E, Kone-Paut I, Giurgea I, Georgin-Lavialle S, et al. Chronic hepatic involvement in the clinical spectrum of A20 haploinsufficiency. Liver International 2021; 41:1894–900.

9. Takagi M, Ogata S, Ueno H, Yoshida K, Yeh T, Hoshino A, et al. Haploinsufficiency of TNFAIP3 (A20) by germline mutation is involved in autoimmune lymphoproliferative syndrome. Journal of Allergy and Clinical Immunology 2017; 139:1914–22.

10. Hori T, Ohnishi H, Kadowaki T, Kawamoto N, Matsumoto H, Ohara O, et al. Autosomal dominant Hashimoto’s thyroiditis with a mutation in *TNFAIP3*. Clinical Pediatric Endocrinology 2019; 28:91–6.

11. Cao C, Fu X, Wang X. Case Report: A novel mutation in TNFAIP3 in a patient with type 1 diabetes mellitus and haploinsufficiency of A20. Frontiers in Endocrinology 2023; 14.

12. Kadowaki T, Kadowaki S, Ohnishi H. A20 Haploinsufficiency in East Asia. Frontiers in Immunology 2021; 12.

13. Aeschlimann FA, Batu ED, Canna SW, Go E, Gül A, Hoffmann P, et al. A20 haploinsufficiency (HA20): clinical phenotypes and disease course of patients with a newly recognised NF-kB-mediated autoinflammatory disease. Annals of the Rheumatic Diseases 2018; 77:728–35.

14. Zhang C, Han X, Sun L, Yang S, Peng J, Chen Y, et al. Novel loss-of-function mutations in TNFAIP3 gene in patients with lupus nephritis. Clinical Kidney Journal 2022; 15:2027–38.

15. Wu CW, Sasa G, Salih A, Nicholas S, Vogel TP, Cahill G, et al. Complicated Diagnosis and Treatment of HA20 due to Contiguous Gene Deletions involving 6q23.3. Journal of Clinical Immunology 2021; 41:1420–3.

16. Polykratis A, Martens A, Eren RO, Shirasaki Y, Yamagishi M, Yamaguchi Y, et al. A20 prevents inflammasome-dependent arthritis by inhibiting macrophage necroptosis through its ZnF7 ubiquitin-binding domain. Nature Cell Biology 2019; 21:731–42.

17. Razani B, Whang MI, Kim FS, Nakamura MC, Sun X, Advincula R, et al. Non-catalytic ubiquitin binding by A20 prevents psoriatic arthritis–like disease and inflammation. Nature Immunology 2020; 21:422–33.

18. Elhani I, Riller Q, Boursier G, Hentgen V, Rieux-Laucat F, Georgin-Lavialle S. A20 Haploinsufficiency: A Systematic Review of 177 Cases. Journal of Investigative Dermatology 2024; 144:1282–94.e8.

19. Kadowaki T, Ohnishi H, Kawamoto N, Hori T, Nishimura K, Kobayashi C, et al. Haploinsufficiency of A20 causes autoinflammatory and autoimmune disorders. Journal of Allergy and Clinical Immunology 2018; 141:1485–8.e11.

20. Deuitch NT, Schwartz DM, Aksentijevich I. Haploinsufficiency of A20. In: Adam MP, Feldman J, Mirzaa GM, Pagon RA, Wallace SE, Amemiya A, editors. GeneReviews(®). Seattle (WA): University of Washington, Seattle Copyright © 1993-2025, University of Washington, Seattle. GeneReviews is a registered trademark of the University of Washington, Seattle. All rights reserved.; 1993.

21. Richards S, Aziz N, Bale S, Bick D, Das S, Gastier-Foster J, et al. Standards and guidelines for the interpretation of sequence variants: a joint consensus recommendation of the American College of Medical Genetics and Genomics and the Association for Molecular Pathology. Genetics in Medicine 2015; 17:405–24.

22. International Team for the Revision of the International Criteria for Behçet’s D, Davatchi F, Assaad-Khalil S, Calamia KT, Crook JE, Sadeghi-Abdollahi B, et al. The International Criteria for Behçet’s Disease (ICBD): a collaborative study of 27 countries on the sensitivity and specificity of the new criteria. Journal of the European Academy of Dermatology and Venereology 2014; 28:338–47.

23. Petty RE, Southwood TR, Manners P, Baum J, Glass DN, Goldenberg J, et al. International League of Associations for Rheumatology classification of juvenile idiopathic arthritis: second revision, Edmonton, 2001. J Rheumatol 2004; 31:390–2.

24. Martini A, Ravelli A, Avcin T, Beresford MW, Burgos-Vargas R, Cuttica R, et al. Toward New Classification Criteria for Juvenile Idiopathic Arthritis: First Steps, Pediatric Rheumatology International Trials Organization International Consensus. The Journal of Rheumatology 2019; 46:190–7.

25. Hochberg MC. Updating the American College of Rheumatology revised criteria for the classification of systemic lupus erythematosus. Arthritis Rheum 1997; 40:1725.

26. Petri M, Orbai A-M, Alarcón GS, Gordon C, Merrill JT, Fortin PR, et al. Derivation and validation of the Systemic Lupus International Collaborating Clinics classification criteria for systemic lupus erythematosus. Arthritis & Rheumatism 2012; 64:2677–86.

27. Sag E, Tartaglione A, Batu ED, Ravelli A, Khalil SM, Marks SD, et al. Performance of the new SLICC classification criteria in childhood systemic lupus erythematosus: a multicentre study. Clin Exp Rheumatol 2014; 32:440–4.

28. Amarilyo G, Rothman D, Manthiram K, Edwards KM, Li SC, Marshall GS, et al. Consensus treatment plans for periodic fever, aphthous stomatitis, pharyngitis and adenitis syndrome (PFAPA): a framework to evaluate treatment responses from the childhood arthritis and rheumatology research alliance (CARRA) PFAPA work group. Pediatric Rheumatology 2020; 18:31.

29. Fautrel B, Mitrovic S, De Matteis A, Bindoli S, Antón J, Belot A, et al. EULAR/PReS recommendations for the diagnosis and management of Still’s disease, comprising systemic juvenile idiopathic arthritis and adult-onset Still’s disease. Ann Rheum Dis 2024; 83:1614–27.

30. Catrysse L, Vereecke L, Beyaert R, van Loo G. A20 in inflammation and autoimmunity. Trends in Immunology 2014; 35:22–31.

31. Yu J, Li H, Wu Y, Luo M, Chen S, Shen G, et al. Inhibition of NLRP3 inflammasome activation by A20 through modulation of NEK7. Proceedings of the National Academy of Sciences 2024; 121:e2316551121.

32. Vande Walle L, Van Opdenbosch N, Jacques P, Fossoul A, Verheugen E, Vogel P, et al. Negative regulation of the NLRP3 inflammasome by A20 protects against arthritis. Nature 2014; 512:69–73.

33. Coornaert B, Carpentier I, Beyaert R. A20: Central Gatekeeper in Inflammation and Immunity*. Journal of Biological Chemistry 2009; 284:8217–21.

34. Chu Y, Vahl JC, Kumar D, Heger K, Bertossi A, Wójtowicz E, et al. B cells lacking the tumor suppressor TNFAIP3/A20 display impaired differentiation and hyperactivation and cause inflammation and autoimmunity in aged mice. Blood 2011; 117:2227–36.

35. Schultheiß C, Paschold L, Mohebiany AN, Escher M, Kattimani YM, Müller M, et al. A20 haploinsufficiency disturbs immune homeostasis and drives the transformation of lymphocytes with permissive antigen receptors. Science Advances 2024; 10:eadl3975.

36. Schwartz DM, Blackstone SA, Sampaio-Moura N, Rosenzweig S, Burma AM, Stone D, et al. Type I interferon signature predicts response to JAK inhibition in haploinsufficiency of A20. Annals of the Rheumatic Diseases 2020; 79:429–31.

37. Nocturne G, Boudaoud S, Miceli-Richard C, Viengchareun S, Lazure T, Nititham J, et al. Germline and somatic genetic variations of TNFAIP3 in lymphoma complicating primary Sjögren’s syndrome. Blood 2013; 122:4068–76.

38. Kato M, Sanada M, Kato I, Sato Y, Takita J, Takeuchi K, et al. Frequent inactivation of A20 through gene mutation in B-cell lymphomas. Rinsho Ketsueki 2011; 52:313–9.

39. Shi Y, Wang X, Wang J, Wang X, Zhou H, Zhang L. The dual roles of A20 in cancer. Cancer Letters 2021; 511:26–35.

40. Allen S, McNeill A, McDermott C, Boardman F, Howard J. The attitudes of individuals with or at risk of adult-onset genetic conditions on reproductive genetic testing: A systematic review. J Genet Couns 2025; 34:e70079.

41. Hemmesch AR, Bogart KR, Barnes E. “Lack” and “Finally”: A Qualitative Analysis of Barriers and Facilitators in Rare Disease Healthcare. Int J Environ Res Public Health 2025; 22.

42. Mulhern CM, Hong Y, Omoyinmi E, Jacques TS, D’Arco F, Hemingway C, et al. Janus kinase 1/2 inhibition for the treatment of autoinflammation associated with heterozygous TNFAIP3 mutation. J Allergy Clin Immunol 2019; 144:863–6.e5.

43. Ozen S, Aksentijevich I. The past 25 years in paediatric rheumatology: insights from monogenic diseases. Nat Rev Rheumatol 2024; 20:585–93.

