## Supplementary figures and images for "Multicenter international cohort study of HA20 reveals novel genetic architecture and phenotypic evolution"

### Supplementary Figures

#### Supplementary Figure 1

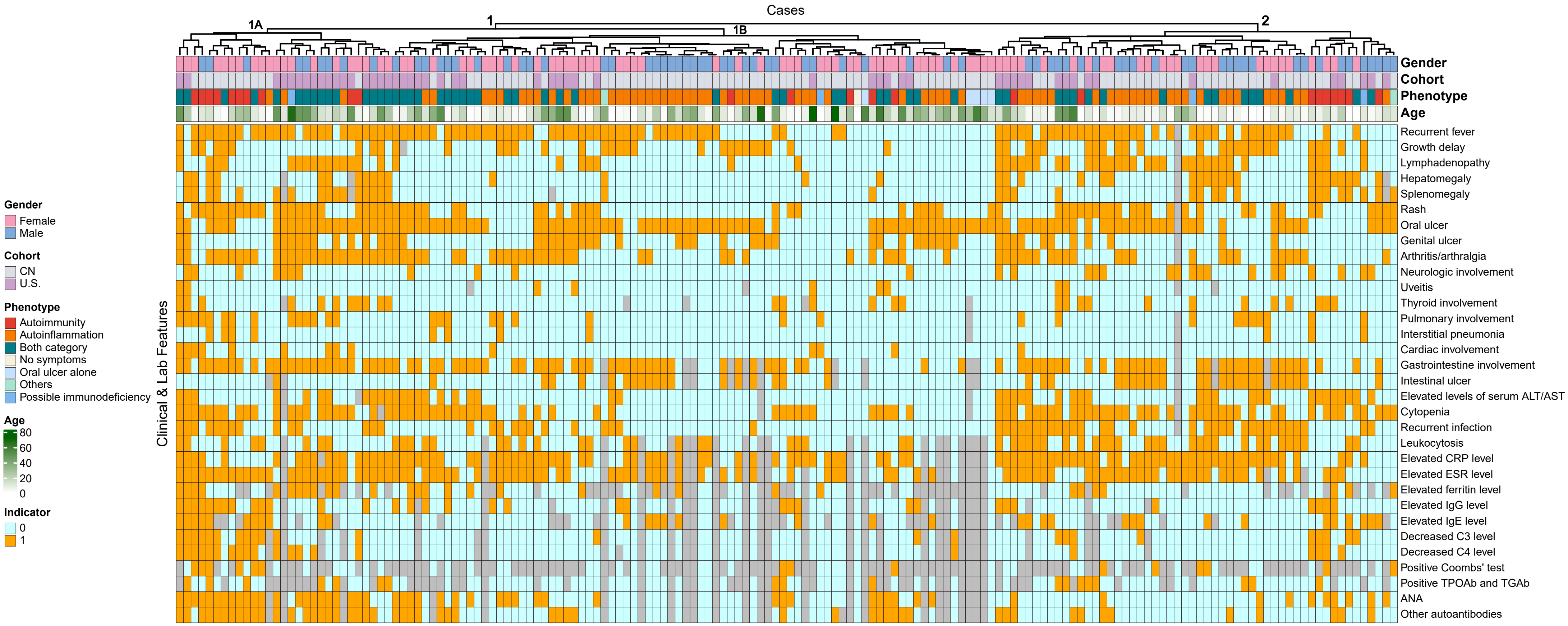

#### Supplementary Figure 2

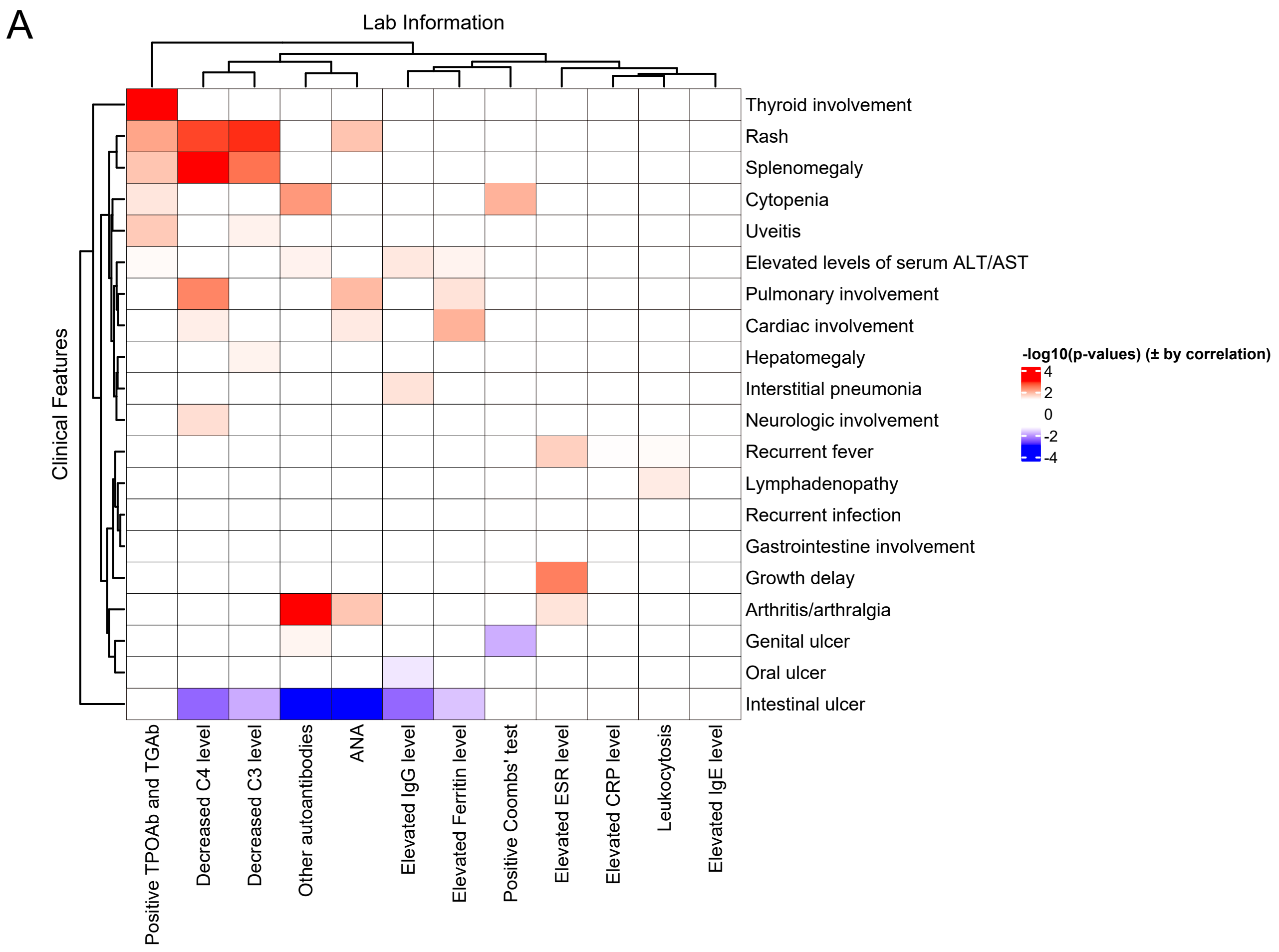


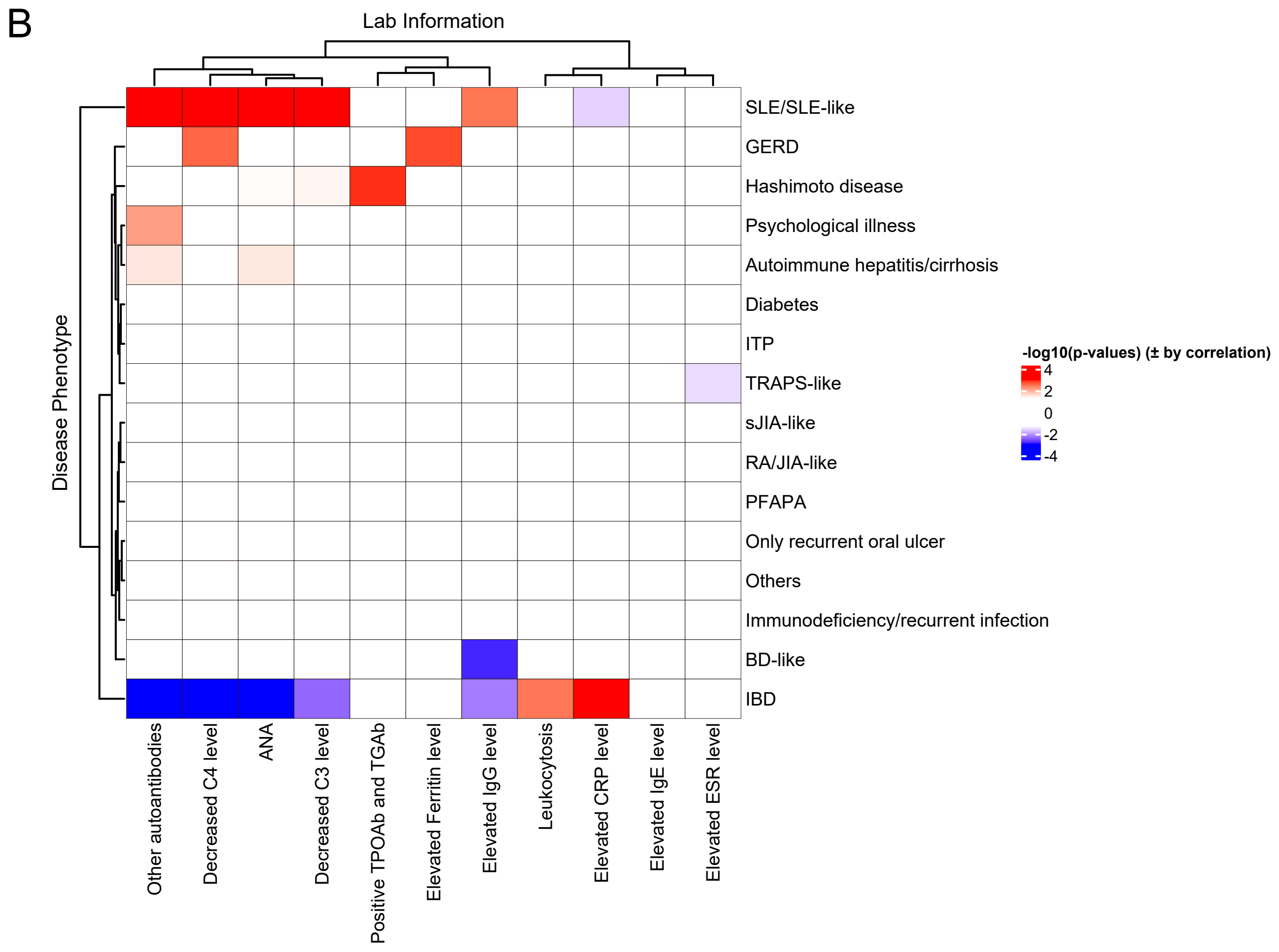

#### Supplementary Figure 3

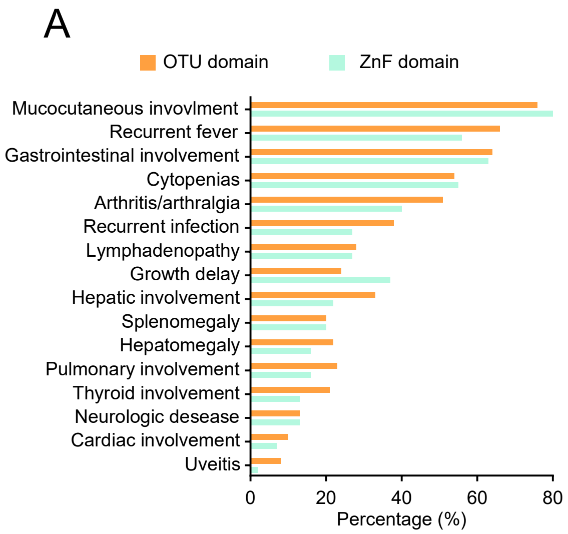


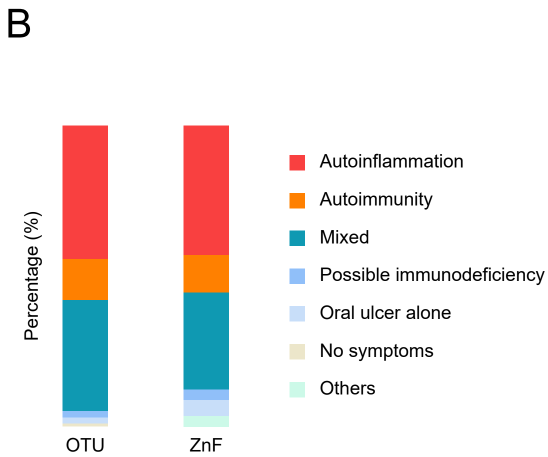

#### Supplementary Figure 4

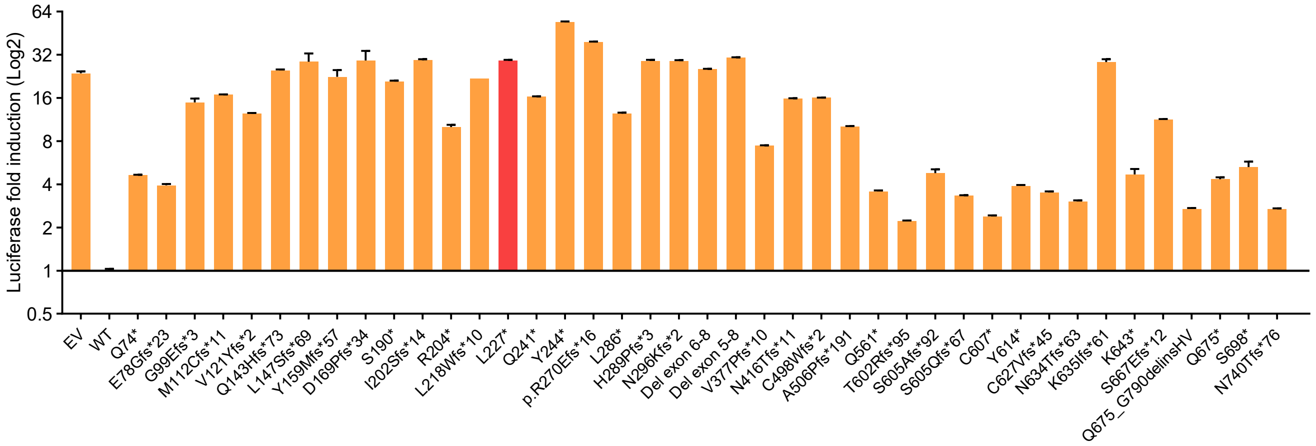
