## Supplementary Figure Legends for "Multicenter international cohort study of HA20 reveals novel genetic architecture and phenotypic evolution"

**Supplementary Fig 1. Hamming-based hierarchical clustering of patients**

Two major groups were identified, including Group 1 (n = 110; median age, 13 years) and Group 2 (n = 54; median age, 5.95 years) (*P* = 0.00027, Wilcoxon rank-sum test). CN, China; CRP, C-reactive protein; ESR, erythrocyte sedimentation rate；ANA, antinuclear antibodies

**Supplementary Fig 2. Correlation analysis between laboratory abnormalities and clinical phenotypes**

**A**, Correlation analysis between laboratory abnormalities and clinical manifestations using Fisher’s exact tests and Pearson correlation. Significant associations (*P* < 0.05) are displayed in red, with the darkest shade corresponding to *P* < 0.001. The color scale represents correlation direction, where positive correlations are shown in red and negative correlations in blue. ALT, alanine transaminase; AST, aspartate transaminase; ESR, erythrocyte sedimentation rate; CRP, C-reactive protein; ANA, antinuclear antibodies; TPOAb, anti-thyroid peroxidase antibody; TGAb, anti-thyroglobulin antibody.

**B**, Correlation analysis between laboratory abnormalities and disease phenotypes using Fisher’s exact tests and Pearson correlation. Significant associations (*P* < 0.05) are displayed in red, with the darkest shade corresponding to *P* < 0.001. The color scale reflects correlation direction as above.

SLE, systemic lupus erythematosus; GERD, gastroesophageal reflux disease; ITP, immune thrombocytopenia; TRAPS, tumor necrosis factor receptor-associated periodic syndrome; sJIA, systemic juvenile idiopathic arthritis; RA, rheumatoid arthritis; PFAPA, periodic fever with aphthous pharyngitis and adenitis; BD, Behçet’s disease; IBD, inflammatory bowel disease; ESR, erythrocyte sedimentation rate; CRP, C-reactive protein; ANA, antinuclear antibodies; TPOAb, anti-thyroid peroxidase antibody; TGAb, anti-thyroglobulin antibody.

**Supplementary Fig. 3 Genotype-phenotype analysis of HA20**

**A**, Bar graph shows percentage of patients with the OTU domain mutations or the ZnF domain mutations presenting with various clinical manifestations (all *P* > 0.05, Mann-Whitney U test).

**B**, Stacked bar chart with 100% total shows percentage of patients with OTU or ZnF domain presenting with autoinflammation-predominant disease, autoimmunity predominant disease, mixed autoinflammation-autoimmunity disease, or other predominant disease phenotypes (all *P* > 0.05, Mann-Whitney U test).

**Supplementary Fig. 4 Functional validation of novel coding mutations of A20**

HEK293T cells were transiently transfected with a NF-κB reporter plasmid, Renilla luciferase control vector, and expression plasmids for either GFP-tagged wild-type or mutant A20. A known pathogenic variant L227* served as a positive control. Results are plotted as luc/(ren x the mean value of WT) to compensate for differences in transfection efficiency. One representative result of three independent experiments is shown. Values are expressed as mean of duplicates ± S.E.M. Un, cells were transfected with empty vector but without TNF stimulation. EV, empty vector; WT, wild type.
