## Supplementary Tables for "Multicenter international cohort study of HA20 reveals novel genetic architecture and phenotypic evolution"

The file includes

Supplementary Table 1 to Table 10

**Supplementary Table 1 Clinical manifestations of HA20 patients.**

| General characteristics | Total | China | US | *P* value |
| --- | --- | --- | --- | --- |
| Male gender, n (%) | 78 (44.8, n = 174) | 52 (44.8) | 26 (44.8, n = 58) | > 0.9999 |
| Children (< 16 y), n (%) | 109 (63.3, n = 172) | 85 (73.3) | 24 (39.3, n = 61) | 0.0005 |
| Median age at onset (y) | 3.3 (n = 161) | 3 (n = 105) | 4.5 (n = 56) | 0.09 |
| Median age at diagnosis (y) | 10.2 (n = 165) | 6 (n = 111) | 18.9 (n = 54) | < 0.0001 |
| Median duration of follow-up (m) | 15 (n =144) | 23 (n = 88) | 1.2 (n = 56) | < 0.0001 |
| Clinical features, n (%) | n = 169 | n = 110 | n = 59 |  |
| Growth delay (n = 168) | 56 (33.3) | 43 (39.1) | 13 (22.4, n =58) | 0.0385 |
| Recurrent fever | 107 (63.3) | 66 (60) | 41 (69.5) | 0.2448 |
| Lymphadenopathy | 54 (32.0) | 27 (24.5) | 27 (45.8) | 0.006 |
| Hepatomegaly (n = 165) | 37 (21.9) | 22 (20) | 15 (27.3, n = 55) | 0.32 |
| Splenomegaly (n = 165) | 37 (21.9) | 16 (14.5) | 21 (38.2, n = 55) | 0.0008 |
| Recurrent infection | 60 (35.5) | 36 (32.7) | 24 (40.7) | 0.3164 |
| Respiratory tract infection | 41 (24.3) | 17 (15.5) | 24 (40.7) | 0.0004 |
| Gastrointestinal infection | 13 (7.7) | 7 (6.4) | 6 (10.2) |  |
| Salmonella infection | 6 (3.6) | 3 (2.7) | 3 (5.1) |  |
| Urinary tract infection | 11 (6.5) | 3 (2.7) | 8 (13.6) |  |
| Suppurative lymphnoditis | 2 (1.2) | 2 (1.8) | 0 |  |
| Septic shock | 3 (1.8) | 1 (0.9) | 2 (3.4) |  |
| Mucocutaneous involvement | 136 (80.5) | 81 (73.6) | 55 (93.2) | 0.002 |
| Rash | 74 (43.8) | 39 (35.4) | 35 (59.3) | 0.0035 |
| Urticaria-like rash | 16 (9.5) | 8 (7.3) | 8 (13.6) |  |
| Pustular eruption | 16 (9.5) | 3 (2.7) | 13 (22.0) |  |
| Vasculitis-like lesion | 8 (4.7) | 3 (2.7) | 5 (8.5) |  |
| Purpura | 2 (1.2) | 2 (1.8) | 0 |  |
| Epifolliculitis | 11 (6.5) | 4 (3.6) | 7 (11.9) |  |
| Chilblain-like rash | 6 (3.6) | 3 (2.7) | 3 (5.1) |  |
| Erythema nodosum | 6 (3.6) | 3 (2.7) | 3 (5.1) |  |
| Subcutaneous edema | 3 (1.8) | 1 (0.9) | 2 (3.4) |  |
| Vitiligo-like rash | 3 (1.8) | 1 (0.9) | 2 (3.4) |  |
| Non-specific rash | 18 (10.7) | 9 (8.2) | 9 (15.3) |  |
| Lipodystrophy | 1 (0.6) | 0 | 1 (1.7) |  |
| Alopecia | 3 (1.8) | 0 | 3 (5.1) |  |
| Acanthosis nigricans | 1 (0.6) | 0 | 1 (1.7) |  |
| Migratory glossitis | 4 (2.4) | 2 (1.8) | 2 (3.4) |  |
| Oral ulcers | 122 (72.2) | 69 (60.9) | 53 (89.8) | 0.0003 |
| Genital ulcers | 54 (32.0) | 22 (20) | 32 (54.2) | < 0.0001 |
| Perianal ulcers | 9 (5.3) | 4 (3.6) | 5 (8.5) | 0.28 |

**Supplementary Table 1** **(continued).**

| General characteristics | Total | China | US | *P* value |
| --- | --- | --- | --- | --- |
| Gastrointestinal involvement | 99 (58.6) | 63 (57.3) | 36 (61.0) | 0.734 |
| Abdominal pain | 62 (37.6) | 30 (28.3, n= 106) | 32 (54.2) | 0.0014 |
| Diarrhea | 64 (37.9) | 36 (32.7) | 28 (47.5) | 0.0686 |
| Esophageal ulcers (n = 157) | 6 (3.8) | 3 (3.0, n = 99)) | 3 (5.2, n = 58) | 0.67 |
| Gastric ulcers (n = 157) | 10 (6.4) | 9 (9.1, n = 99) | 1 (1.7, n = 58) | 0.09 |
| Intestinal ulcers (n = 155) | 43 (27.7) | 39 (40.2, n = 97) | 4 (6.9, n = 58) | < 0.0001 |
| Intestinal hemorrhage | 10 (6.0) | 8 (7.3) | 2 (3.4, n = 58) |  |
| Intestinal perforation | 6 (3.6) | 4 (3.6) | 2 (3.4, n = 58) |  |
| GERD | 8 (4.8) | 0 | 8 (13.8, n = 58) |  |
| Atrophic autoimmune gastritis | 1 (0.6) | 0 | 1 (1.7, n = 58) |  |
| Recurrent intussusception | 1 (0.6) | 1 (0.9) | 0 |  |
| Diverticulosis | 1 (0.6) | 0 | 1 (1.7, n = 58) |  |
| Eosinophilic esophagitis | 1 (0.6) | 0 | 1 (1.7, n = 58) |  |
| Hepatic involvement (n = 166) | 48 (28.9) | 17 (15.7, n = 108) | 31 (56.9, n = 58) | < 0.0001 |
| Jaundice | 7 (4.2) | 1 (0.9) | 6 (10.3) |  |
| Elevated levels of serum ALT/AST | 47 (28.3) | 17 (15.7) | 30 (51.7) | < 0.0001 |
| Autoimmune hepatitis | 19 (11.4) | 8 (7.4) | 11 (19.0) | 0.0745 |
| Interface hepatitis | 1 (0.6) | 0 | 1 (1.7) |  |
| Cirrhosis | 4 (2.4) | 1 (0.9) | 3 (5.2) | 0.15 |
| Cytopenias (n = 166) | 94 (56.6) | 47 (43.5, n = 108) | 47 (81.0, n = 58) | < 0.0001 |
| Anemia | 71 (42.8) | 41 (38.0) | 30 (51.7) | 0.1 |
| Neutropenia | 30 (18.1) | 12 (11.1) | 18 (31.0) | 0.0027 |
| Lymphopenia | 45 (27.1) | 9 (8.3) | 36 (62.1) | < 0.0001 |
| Thrombocytopenia | 29 (17.5) | 12 (11.1) | 17 (29.3) | 0.0049 |
| Musculoskeletal system | 80 (47.3) | 36 (32.7) | 44 (74.6) | < 0.0001 |
| Arthritis/arthralgia | 79 (46.7) | 35 (31.8) | 44 (74.6) | < 0.0001 |
| Myositis | 4 (2.4) | 2 (1.8) | 2 (3.4) |  |
| Myalgia without myositis | 12 (7.1) | 0 | 12 (20.3) |  |
| Thyroid involvement (n = 165) | 29 (17.6) | 11 (10.3, n = 107) | 18 (31.0, n = 58) | 0.0012 |
| Hypothyroidism | 19 (11.5) | 7 (6.5) | 12 (20.7) |  |
| Temporary hyperthyroidism | 3 (1.8) | 1 (0.9) | 2 (3.4) |  |
| Hashimoto disease | 15 (9.1) | 7 (6.5) | 8 (13.8) |  |
| Other thyroiditis | 7 (4.2) | 3 (2.8) | 4 (6.9) |  |

**Supplementary Table 1** **(continued).**

| General characteristics | Total | China | US | *P* value |
| --- | --- | --- | --- | --- |
| Neurologic involvement | 28 (16.6) | 12 (10.9) | 16 (27.1) | 0.009 |
| Seizure | 10 (5.9) | 8 (7.3) | 2 (3.4) |  |
| Meningitis | 7 (4.1) | 3 (2.7) | 4 (6.9) |  |
| Vasculitis | 4 (2.4) | 1 (0.9) | 3 (5.2) |  |
| Stroke | 4 (2.4) | 1 (0.9) | 3 (5.2) |  |
| Developmental delay | 10 (5.9) | 1 (0.9) | 9 (15.3) |  |
| Sensorineural hearing loss | 3 (1.8) | 0 | 3 (5.2) |  |
| Peripheral neuropathy | 2 (1.2) | 0 | 2 (3.4) |  |
| Pulmonary involvement (n = 168) | 34 (20.2) | 20 (18.3, n = 109) | 14 (23.7) | 0.4263 |
| Pleural effusion | 12 (7.1) | 7 (6.4) | 5 (8.5) |  |
| Interstitial pneumonia | 11 (6.5) | 10 (9.2) | 1 (1.7) |  |
| Pulmonary nodules | 11 (6.5) | 2 (1.8) | 9 (15.3) |  |
| Asthma | 4 (2.4) | 0 | 4 (6.8) |  |
| Pulmonary cyst | 1 (0.6) | 1 (0.9) | 0 |  |
| Pulmonary consolidation | 3 (1.8) | 2 (1.8) | 1 (1.7) |  |
| Pneumothorax | 1 (0.6) | 1 (0.9) | 0 |  |
| Pulmonary hypertension | 1 (0.6) | 0 | 1 (1.7) |  |
| Cardiac involvement | 16 (9.5) | 6 (5.5) | 10 (16.9) | 0.0248 |
| Pericardial effusion | 12 (7.1) | 4 (3.6) | 8 (13.6) |  |
| Decreased ejection fraction | 5 (3.0) | 3 (2.7) | 2 (3.4) |  |
| Coronary vasculitis/artery dilation | 5 (3.0) | 1 (0.9) | 4 (6.8) |  |
| Non-compaction cardiomyopathy | 1 (0.6) | 0 | 1 (1.7) |  |
| Congenital heart disease | 2 (1.2) | 2 (1.8) | 0 |  |
| Ophthalmologic involvement (n = 166) | 10 (6.0) | 1 (0.9, n = 107) | 9 (15.3) | 0.0004 |
| Retinal vasculitis | 2 (1.2) | 1 (0.9) | 1 (1.7) |  |
| Macular degeneration | 2 (1.2) | 1 (0.9) | 1 (1.7) |  |
| Uveitis | 9 (5.4) | 0 | 9 (15.3) | < 0.0001 |
| Xerophthalmia | 4 (2.4) | 0 | 4 (6.8) |  |
| Cataracts | 5 (3.0) | 2 (1.8) | 3 (5.1) |  |
| Corneal ulcers | 1 (0.6) | 0 | 1 (1.7) |  |
| Vitreous hemorrhage | 1 (0.6) | 0 | 1 (1.7) |  |

**Supplementary Table 1** **(continued).**

| General characteristics | Total | China | US | *P* value |
| --- | --- | --- | --- | --- |
| Others |  |  |  |  |
| Raynaud phenomenon | 10 (5.9) | 2 (1.8) | 8 (13.6) |  |
| Diabetes | 4 (2.4) | 1 (0.9) | 3 (5.1) |  |
| Depression disorder | 3 (1.8) | 0 | 3 (5.1) |  |
| Generalized anxiety disorder | 2 (1.2) | 0 | 2 (3.4) |  |
| Schizophrenia | 1 (0.6) | 0 | 1 (1.7) |  |
| ADHD | 2 (1.2) | 0 | 2 (3.4) |  |
| Autism spectrum disorder | 1 (0.6) | 0 | 1 (1.7) |  |
| Premature ovarian failure | 1 (0.6) | 0 | 1 (1.7) |  |
| Fibromyalgia | 3 (1.8) | 0 | 3 (5.1) |  |
| Varicose veins | 1 (0.6) | 0 | 1 (1.7) |  |
| B cell lymphoma | 3 (1.8) | 0 | 3 (5.1) |  |
| Large-vessel vasculitis | 1 (0.6) | 0 | 1 (1.7) |  |
| Ruptured corpus luteal cyst | 1 (0.6) | 0 | 1 (1.7) |  |
| Endometriosis | 1 (0.6) | 0 | 1 (1.7) |  |
| Recurrent Bell's Palsy | 3 (1.8) | 0 | 3 (5.1) |  |
| Autoimmune polyglandular syndrome | 1 (0.6) | 0 | 1 (1.7) |  |
| CVID | 2 (1.2) | 0 | 2 (3.4) |  |
| Microangiopathic hemolytic anemia | 1 (0.6) | 0 | 1 (1.7) |  |

ALT: alanine transaminase; AST: aspartate transaminase; GERD: Gastroesophageal reflux disease; ADHD: Attention deficit and hyperactivity disorder; CVID: common variable immunodeficiency. If clearly labeled, the number of patients in total equals the number, or the sum of numbers, in parentheses. Comparisons stratified by country of origin were performed using Mann-Whitney U test.

**Supplementary Table 2 Clinical features of each HA20 patient.**

| Cases | Gender | Age range,  onset (+/- 5 years) | Age range,  diagnosis (+/- 5 years) | Main disease phenotype before diagnosis of HA20 | Laboratory findings related to inflammation and autoimmunity at diagnosis or during disease flare |
| --- | --- | --- | --- | --- | --- |
| P1 | Male | 0-5 | 5-10 | Autoimmune hepatitis, Hashimoto disease | Serum IgG↑, FT3↓, FT4↓, TSH↑, AST↑, ALT↑, TPOAb (+), TGAb (+), C3↓, C4↓ |
| P2 | Male | 0-5 | 0-5 | IBD, Arthritis | CRP↑, ESR↑, SAA↑ |
| P3 | Male | 0-5 | 0-5 | IBD, Arthritis | WBC↑, CRP↑, ESR↑, SAA↑ |
| P4 | Male | 5-10 | 5-19 | IBD, Arthritis | WBC↑, CRP↑, ESR↑, SAA↑ |
| P5 | Male | 0-5 | 0-5 | PFAPA, Aplastic anemia-like, Interstitial pneumonia | ESR↑, FER↑, IgG↑, dsDNA (+) |
| P6  (P5's mother) | Female | 5-10 | 25-30 | BD-like, Cerebral infarction | CRP↑, ESR↑, SAA↑ |
| P7 | Male | 0-5 | 10-15 | IBD | CRP↑, ESR↑, SAA↑ |
| P8 (P7's father) | Male | 20-25 | 35-40 | BD-like, IBD, Arthritis | CRP↑, ESR↑, ANA (+) |
| P9 | Female | 0-5 | 15-20 | Refractory SLE | Serum IgG↑, ESR↑, ANA (+), anti-dsDNA (+), anti-SSA (+), anti-Scl70(+), anti-U1 RNP (+), antiribosomal P protein (+), C4↓ |
| P10 | Male | 0-5 | 0-5 | Thyroiditis, IBD, Pulmonary cysts, Autoimmune hepatitis | WBC↑, CRP↑, ESR↑, SAA↑, serum IgE↑, TPOAb(+), TGAb(+) |
| P11 | Female | 0-5 | 5-10 | IBD, PFAPA-like, Immunodeficiency, Ventriculomegaly | WBC↑, ESR↑, SAA↑, FER↑, ANA (+), Coomb's test (+) |
| P12 | Male | 0-5 | 5-10 | BD-like, PFAPA, IBD | ESR↑, serum IgE↑ |

**Supplementary Table 2** **(continued).**

| Cases | Gender | Age range,  onset (+/- 5 years) | Age range,  diagnosis (+/- 5 years) | Main disease phenotype before diagnosis of HA20 | Laboratory findings related to inflammation and autoimmunity at diagnosis or during disease flare |
| --- | --- | --- | --- | --- | --- |
| P13 | Male | 0-5 | 5-10 | BD-like, Arthritis, IBD, Atrial septal defect | CRP↑, ESR↑ |
| P14 | Female | 0-5 | 0-5 | IBD, Autoimmune hepatitis, Possible immunodeficiency, Pneumonia | WBC↑, CRP↑, ESR↑, SAA↑, ANA (±), anti-dsDNA (+), anti-Scl (+) |
| P15 | Female | 0-5 | 0-5 | IBD, possible immunodeficiency, interstitial pneumonia | WBC↑, CRP↑ |
| P16  (P17's mother) | Female | 20-25 | 25-30 | SLE-like, epilepsy, acute cerebral infarction, depression, acoustic neuroma | WBC↓, ESR↑, ANA (+), anti-dsDNA (+), anti-SSA (+), anti-cardiolipin (+), Coomb's test (+), C3↓, C4↓ |
| P17 | Female | No phenotype | 0-5 | None | - |
| P18 | Male | 25-30 | 25-30 | SLE-like | WBC↓, ESR↑, serum IgE↑, ANA (+), Anti-C1q (+), Anti-SSA-Ro60(+++), Anti-SSA-Ro52(+++), Anti-RPA (+++), TGAb (+), C3↓, C4↓ |
| P19  (P18's son) | Male | 5-10 | 5-10 | Henoch-Schoenlein purpura, Duodenal ulcer, Anemia | Serum IgE↑ |
| P20  (P18's mother) | Female | Unsure | 55-60 | Recurrent oral ulcer | NA |
| P21 | Female | 0-5 | 0-5 | BD-like, IBD | WBC↑, CRP↑, serum IL-6, TNF-a, and IFN-γ↑ |
| P22 | Female | 5-10 | 10-15 | BD-like, IBD | WBC↑, CRP↑, SAA↑, ANA (±), anti-dsDNA (+), anti-Scl (+) |

**Supplementary Table 2** **(continued).**

| Cases | Gender | Age range,  onset (+/- 5 years) | Age range,  diagnosis (+/- 5 years) | Main disease phenotype before diagnosis of HA20 | Laboratory findings related to inflammation and autoimmunity at diagnosis or during disease flare |
| --- | --- | --- | --- | --- | --- |
| P23  (P22's mother) | Female | 10-15 | 30-35 | Recurrent oral ulcer | NA |
| P24 | Male | 0-5 | 0-5 | BD-like, Possible immunodeficiency | ESR↑ |
| P25 | Female | 0-5 | 0-5 | PFAPA | WBC↑, CRP↑ |
| P26 | Female | 0-5 | 0-5 | IBD | WBC↑, CRP↑, ESR↑ |
| P27  (P24's mother) | Female | 20-25 | 30-35 | Recurrent oral ulcer | NA |
| P28 | Female | 0-5 | 0-5 | IBD | CRP↑, ESR↑, ANA (+), Anti-cardiolipin (+) |
| P29 | Male | 5-10 | 10-15 | BD-like, IBD, Arthritis | CRP↑, ESR↑ |
| P30 | Female | 0-5 | 0-5 | SLE-like, Immunodeficiency | ESR↑, serum IgG↑, ANA (+), anti-dsDNA(+), anti-SSA (+), anti-cardiolipin (+), C3↓, C4↓ |
| P31 | Female | 0-5 | 0-5 | BD-like, PFAPA | WBC↑, CRP↑, ESR↑, SAA↑ |
| P32 | Male | 5-10 | 10-15 | SLE-like, HLH secondary to EBV infection | WBC↓, ESR↑, serum IgG↑, serum IgE↑, ANA (+), anti-SSA (+), Coomb's test(+), TPOAb(+), TGAb (+), C3↓, C4↓ |
| P33 | Female | 0-5 | 0-5 | IBD, Arthritis | WBC↑, CRP↑, ESR↑, serum IgG↑, ANA (+) |
| P34 | Female | 0-5 | 0-5 | BD-like | CRP↑, ESR↑ |
| P35 | Female | 0-5 | 0-5 | PFAPA, Thyroiditis, Pulmonary nodules | CRP↑, ESR↑, Coomb's test(+), TPOAb(+), TGAb(+) |

**Supplementary Table 2** **(continued).**

| Cases | Gender | Age range,  onset (+/- 5 years) | Age range,  diagnosis (+/- 5 years) | Main disease phenotype before diagnosis of HA20 | Laboratory findings related to inflammation and autoimmunity at diagnosis or during disease flare |
| --- | --- | --- | --- | --- | --- |
| P36 | Female | 0-5 | 0-5 | PFAPA, IBD, Autoimmune hepatitis | WBC↑, CRP↑, Coombs' test (+), serum IgE↑ |
| P37 | Female | 0-5 | 0-5 | SLE-like, Recurrent pericarditis | WBC↓, CRP↑, ESR↑, SAA↑, serum IgG↑, ANA (+), anti-dsDNA(+), Coombs' test (+), C3↓, C4↓, serum IgE↑ |
| P38  (P45's daughter) | Female | 5-10 | 10-15 | SLE-like, Thyroiditis, Autoimmune hepatitis, Hepatic cirrhosis, Hepatic failure | ESR↑, serum IgE↑, serum IgG↑, ANA (+), Coomb's test (+), C3↓, C4↓ |
| P39  (P45's son) | Male | 0-5 | 5-10 | BD-like, IBD | CRP↑, ESR↑, serum IgE↑ |
| P40  (P46's daughter) | Female | 0-5 | 0-5 | SLE-like | WBC↑, CRP↑, ESR↑, SAA↑, serum IgG↑, ANA (+), anti-dsDNA(+), Coombs' test (+) |
| P41  (P46's son) | Male | 5-10 | 5-10 | Suspicion of sJIA | WBC↑, CRP↑, ESR↑, SAA↑, serum IgG↑, ANA (+) |
| P42 | Female | 0-5 | 0-5 | BD-like | ESR↑ |
| P43 | Female | 5-10 | 5-10 | Suspicion of sJIA | WBC↑, CRP↑, ESR↑, FER↑ |
| P44 | Female | 5-10 | 10-15 | SLE-like, Autoimmune hepatitis | Serum IgG↑, ANA (+), anti-SSA(+), C3↓, C4↓, serum IgE↑ |
| P45 | Male | 20-25 | 35-40 | BD-like | NA |
| P46 | Female | 15-20 | 25-30 | SLE-like | NA |
| P47 (P36's father) | Male | 20-25 | 35-40 | BD-like | NA |

**Supplementary Table 2** **(continued).**

| Cases | Gender | Age range,  onset (+/- 5 years) | Age range,  diagnosis (+/- 5 years) | Main disease phenotype before diagnosis of HA20 | Laboratory findings related to inflammation and autoimmunity at diagnosis or during disease flare |
| --- | --- | --- | --- | --- | --- |
| P48 | Female | 0-5 | 0-5 | BD-like, PFAPA | WBC↑, CRP↑, ESR↑, Coomb's test (+) |
| P49 | Female | 0-5 | 0-5 | BD-like, Hyperthyroidism | WBC↑, CRP↑, ESR↑ |
| P50  (P49's mother) | Female | 5-10 | 25-30 | Recurrent oral ulcer | NA |
| P51 | Female | 5-10 | 10-15 | BD-like, IBD | CRP↑, ESR↑ |
| P52 | Female | 0-5 | 5-10 | BD-like | CRP↑, ESR↑ |
| P53 | Female | 10-15 | 10-15 | BD-like | ESR↑, serum IgE↑ |
| P54 | Male | 5-10 | 5-10 | BD-like, PFAPA | CRP↑, ESR↑, serum IgE↑ |
| P55  (P54's mother) | Female | 5-10 | 30-35 | BD-like | NA |
| P56 | Female | 5-10 | 5-10 | SLE-like, Thyroiditis | ESR↑, ANA (+), anti-SSA (+), anti-dsDNA (+), Coombs' test (+) |
| P57 | Female | 0-5 | 0-5 | IBD | WBC↑, CRP↑, ESR↑ |
| P58  (P33's father) | Male | 20-25 | 30-35 | BD-like | CRP↑, ESR↑, SAA↑ |
| P59  (P51’s father) | Male | 30-35 | 40-45 | BD-like | NA |
| P60 | Male | 0-5 | 0-5 | SLE-like, Immunodeficiency, Autoimmune hepatitis | WBC↑, CRP↑, FER↑, serum IgG↑, TPOAb(+), TGAb(+), anti-PAIgG(+), anti-PAIgM(+) |
| P61 | Male | 0-5 | 5-10 | BD-like, IBD | WBC↑, CRP↑, ESR↑, C3↓ |
| P62 | Female | 5-10 | 5-10 | BD-like, IBD | CRP↑, ESR↑, C3↓, serum IgE↑ |

**Supplementary Table 2** **(continued).**

| Cases | Gender | Age range,  onset (+/- 5 years) | Age range,  diagnosis (+/- 5 years) | Main disease phenotype before diagnosis of HA20 | Laboratory findings related to inflammation and autoimmunity at diagnosis or during disease flare |
| --- | --- | --- | --- | --- | --- |
| P63 | Female | 15-20 | 35-40 | BD-like | ESR↑, serum IgG↑, C3↓, ANA (+) |
| P64 | Male | 15-20 | 40-45 | BD-like, IBD, Thyroiditis | CRP↑, ESR↑, FER↑, ANA (+), TPOAb(+), TGAb(+) |
| P65 | Female | 0-5 | 0-5 | PFAPA, IBD? | WBC↑, CRP↑, serum IgG↑, serum IgE↑ |
| P66  (P65's mother) | Female | 0-5 | 35-40 | PFAPA, Hashimoto disease | NA |
| P67 | Male | 0-5 | 0-5 | BD-like, IBD, Arthritis | WBC↑, CRP↑, ESR↑, SAA↑ |
| P68  (P67's brother) | Male | 0-5 | 0-5 | Recurrent oral ulcer, Arthritis | Normal WBC, CRP, and ESR |
| P69  (P67's father) | Male | 30-35 | 35-40 | BD-like, Arthritis | NA |
| P70 | Male | 0-5 | 0-5 | Autoimmune hepatitis | WBC↑, CRP↑, FER↑, C3↓, ANA (+), serum IgE↑ |
| P71  (P63's daughter) | Female | NA | NA | NA | NA |
| P72  (P64's daughter) | Female | NA | NA | NA | NA |
| P73 | Female | 5-10 | 5-10 | SLE-like, Thyroiditis | CRP↑, ESR↑, ANA (+), anti-Sm (+), C3↓, C4↓ |
| P74 | Male | NA | NA | NA | NA |
| P75 | Male | 0-5 | 30-35 | BD-like, PFAPA | NA |
| P76 | Female | 0-5 | 5-10 | BD-like, IBD | WBC↑, CRP↑, ESR↑, SAA↑, serum IgE↑ |
| P77 | Male | 0-5 | 0-5 | BD-like, IBD, Arthritis | CRP↑, ESR↑, serum IgE↑ |

**Supplementary Table 2** **(continued).**

| Cases | Gender | Age range,  onset (+/- 5 years) | Age range,  diagnosis (+/- 5 years) | Main disease phenotype before diagnosis of HA20 | Laboratory findings related to inflammation and autoimmunity at diagnosis or during disease flare |
| --- | --- | --- | --- | --- | --- |
| P78 | Male | 0-5 | 0-5 | IBD | CRP↑, ESR↑, serum IgE↑ |
| P79 | Female | 0-5 | 0-5 | Recurrent fever of unknown origin | ESR↑, SAA↑ |
| P80 | Female | 5-10 | 25-30 | BD-like | NA |
| P81 | Male | 5-10 | 5-10 | BD-like, IBD | WBC↑, CRP↑ |
| P82 | Female | 0-5 | 0-5 | IBD | WBC↑, CRP↑, ESR↑, SAA↑, serum IgG↑, serum IgE↑ |
| P83 | Female | 0-5 | 0-5 | BD-like, IBD, Interstitial pneumonia | WBC↑, CRP↑ |
| P84 | Female | 0-5 | 0-5 | Severe pneumonia, Immunodeficiency? Ventricular septal defect | FER↑ |
| P85 | Female | 0-5 | 0-5 | HLH-like | CRP↑, FER↑, ANA (+) |
| P86 | Male | 10-15 | 10-15 | SLE, Lupus nephritis, Thyroiditis, Panreatitis, Lung infection, Coronary dilation | WBC↓, CRP↑, ESR↑, FER↑, serum IgG↑, ANA (+), p-ANCA(+), Coombs' test (+), C3↓, serum IgE↑ |
| P87 | Male | 10-15 | 20-25 | BD-like, IBD | CRP↑, ESR↑, FER↑, serum IgE↑ |
| P88 (P87's father) | Male | 30-35 | 50-55 | Recurrent oral ulcer | NA |
| P89(P46's mother) | Female | NA | 70-75 | Rheumatic polymyositis | CRP↑, ESR↑, Anti-U1-snRNP(+) |
| P90 | Male | 0-5 | 0-5 | BD-like, Arthritis | ESR↑, serum IgE↑ |
| P91 | Female | 10-15 | 10-15 | BD-like | Normal WBC, CRP, and ESR, C3↓, C4↓ |

**Supplementary Table 2** **(continued).**

| Cases | Gender | Age range,  onset (+/- 5 years) | Age range,  diagnosis (+/- 5 years) | Main disease phenotype before diagnosis of HA20 | Laboratory findings related to inflammation and autoimmunity at diagnosis or during disease flare |
| --- | --- | --- | --- | --- | --- |
| P92 | Female | 0-5 | 0-5 | IBD | WBC↑, C3↓ |
| P93 | Female | 5-10 | 5-10 | BD-like, Thyroiditis, Arthritis | WBC↑ |
| P94  (P93's mother) | Female | NA | NA | NA | NA |
| P95 | Male | 0-5 | 0-5 | BD-like, Hepatitis, Aseptic meningoencephalitis | CRP↑, ESR↑ |
| P96 | Male | 0-5 | 0-5 | PFAPA-like | WBC↑, CRP↑ |
| P97  (P96's father) | Male | NA | NA | NA | NA |
| P98 | Male | 0-5 | 0-5 | BD-like, Arthritis, Immunodeficiency | WBC↑ |
| P99 | Female | 0-5 | 5-10 | Recurrent fever, Arthritis, Immunodeficiency | CRP↑, ESR↑, SAA↑, serum IgG↑ |
| P100 | Male | 15-20 | 35-40 | IBD | NA |
| P101 | Female | 0-5 | 0-5 | SLE-like | WBC↑, ESR↑, ANA (+), Coombs' test (+), C3↓, serum IgE↑ |
| P102 | Female | 0-5 | 0-5 | BD-like | NA |
| P103 | Male | 0-5 | 0-5 | BD-like | NA |
| P104 | Female | 0-5 | 5-10 | PFAPA, SLE, Lupus nephritis | WBC ↑ → WBC↓, ESR↑, ANA (+), Coombs' test (+), serum IgG↑, C3↓, C4↓, ANA (+), anti-dsDNA(+), anti-RNP Ab(+), anti-SSA Ab(+), serum IgE↑ |

**Supplementary Table 2** **(continued).**

| Cases | Gender | Age range,  onset (+/- 5 years) | Age range,  diagnosis (+/- 5 years) | Main disease phenotype before diagnosis of HA20 | Laboratory findings related to inflammation and autoimmunity at diagnosis or during disease flare |
| --- | --- | --- | --- | --- | --- |
| P105 | Male | 5-10 | 10-15 | Recurrent fever, Arthritis | CRP↑, ESR↑ |
| P106 | Male | 0-5 | 10-15 | PFAPA, BD-like | WBC↑, CRP↑, ESR↑, SAA↑, FER↑, serum IgE↑ |
| P107 | Female | 0-5 | 10-15 | IBD | NA |
| P108 | Male | 5-10 | 5-10 | PFAPA, Sclerosing cholangitis | CRP↑, ESR↑ |
| P109 | Male | 0-5 | 0-5 | PFAPA, Immunodeficiency | CRP↑, TPOAb(+), TGAb(+) |
| P110 | Male | 0-5 | 5-10 | HLH-like | WBC↓, FER↑, Coombs' test (+) |
| P111 | Male | NA | 35-40 | Type 1 diabetes, Recurrent infection | NA |
| P112 | Male | 0-5 | 0-5 | IBD, Recurrent infection | WBC↑, CRP↑, ESR↑, SAA↑, Coombs' test (+), serum IgG↑ |
| P113 | Female | NA | 35-40 | BD-like, IBD | WBC↑, CRP↑, ESR↑, SAA↑ |
| P114 | Female | NA | 65-70 | BD-like | NA |
| P115 | Female | 5-10 | 10-15 | Recurrent oral ulcer, Epifolliculitis | Normal WBC, CRP, and ESR |
| P116 | Male | 0-5 | 5-10 | PFAPA | WBC↑, CRP↑, ESR↑ |
| P117 | Female | 0-5 | 5-10 | Autoimmune hepatitis, Hashimoto's disease, ITP | ESR↑, FER↑, serum IgG↑, serum IgE↑, ANA (+), Anti-dsDNA(+) |
| P118 | Male | 10-15 | 70-75 | BD-like | CRP↑ |
| P119 | Female | 10-15 | 25-30 | BD-like, SLE-like, CNS vasculitis | WBC↑, CRP↑, ESR↑, FER↑, IgG↑, IgE↑, C3↓, C4↓, TPOAb(+), TGAb(+), ANA (+), LAC(+), Anti-cardiolipin Ab(+), Anti-SmRNP Ab(+) |
| P120 | Female | 5-10 | 50-55 | BD-like, Pustular psoriasis, SLE-like | CRP↑, ESR↑, Anti-cardiolipin Ab(+), LAC(+) |

**Supplementary Table 2** **(continued).**

| Cases | Gender | Age range,  onset (+/- 5 years) | Age range,  diagnosis (+/- 5 years) | Main disease phenotype before diagnosis of HA20 | Laboratory findings related to inflammation and autoimmunity at diagnosis or during disease flare |
| --- | --- | --- | --- | --- | --- |
| P121 | Female | 5-10 | 25-30 | BD-like, SLE-like | CRP↑, ESR↑, FER↑, C4↓, ANA (+), LAC(+), Anti-RNP Ab(+) |
| P122 | Male | 10-15 | 10-15 | Autoimmune hepatitis, IBD, BD-like | CRP↑, ESR↑, FER↑, p-ANCA(+), Anti-GAD(+) |
| P123 | Female | 0-5 | 10-15 | PFAPA-like, IBD-like, BD-like | WBC↑, CRP↑, ESR↑, IgE↑, |
| P124 | Female | 0-5 | 15-20 | BD-like, IBD-like | WBC↑, CRP↑, ESR↑, ANA (low titer), serum IgD and IgA↑ |
| P125 | Female | 0-5 | 20-25 | Inflammatory arthritis, PFAPA-like | CRP↑, ESR↑, ANA (+), LAC(+) |
| P126 | Female | 15-20 | 50-55 | BD-like | WBC↑, CRP↑, LAC(+) |
| P127 | Female | 0-5 | 5-10 | BD-like, IBD-like, Hashimoto's disease | CRP↑, ESR↑, TPOAb(+), TGAb(+) |
| P128 | Male | 0-5 | 10-15 | Autoimmune hepatitis, BD-like, Lipodystrophy | WBC↑, ESR↑, IgG↑, IgE↑, TPOAb(+), TGAb(+), ANA (+), Anti-smooth muscle Ab(+), Anti-mitochondrial Ab(+), LAC(+), TPOAb(+), Anti-Sm Ab(+), RF(+) |
| P129 | Male | 5-10 | 45-50 | BD-like, IBD-like, Lymphoproliferation, sJIA-like, Diffuse large B-cell lymphoma (jejunum) | WBC↑, CRP↑, ESR↑, FER↑, ANA (+), TTG IgA(+) |
| P130 | Male | 0-5 | 0-5 | PFAPA-like | WBC↑, CRP↑, ESR↑, IgG↑, ANA (+), Anti-dsDNA(+) |

**Supplementary Table 2** **(continued).**

| Cases | Gender | Age range,  onset (+/- 5 years) | Age range,  diagnosis (+/- 5 years) | Main disease phenotype before diagnosis of HA20 | Laboratory findings related to inflammation and autoimmunity at diagnosis or during disease flare |
| --- | --- | --- | --- | --- | --- |
| P131 | Female | 5-10 | 20-25 | SLE-like, RA/JIA, TRAPS-like | WBC↑, CRP↑, ESR↑, FER↑, IgG↑, ANA (+), Anti-CCP(+), RF(+), LAC(+), SSA(+) |
| P132 | Female | 15-20 | 20-25 | SLE-like, BD-like | CRP↑, ESR↑, FER↑, TPOAb(+), TGAb(+), ANA (+), Platelets autoantibodies(+), Anti-RNP Ab(+) |
| P133 | Female | 5-10 | 45-50 | BD-like, TRAPS-like | CRP↑, ESR↑, Anti-cardiolipin Ab(+), LAC(+) |
| P134 | Male | 0-5 | 5-10 | FMF-like, IBD-like | WBC↑, CRP↑, ESR↑, SAA↑, FER↑, IgE↑, ANA (+), Anti-dsDNA(+) |
| P135 | Female | 5-10 | 10-15 | BD-like | IgE↑, ANA (+), Gliadin IgG positive 118.8 (<15) |
| P136 | Male | 5-10 | 15-20 | BD with CNS vasculitis | WBC↑, CRP↑, IgG↑, Anti-SSB ↑, Anti-proteinase-3 Ab(+) , Anti-cardiolipin Ab(+), Anti-histone Ab(+), DRVVT(+) |
| P137 | Male | 5-10 | 50-55 | BD with coronary vasculitis | CRP↑, ESR↑, ANA (+), Anti-SSB Ab(+), Anti-cardiolipin Ab (+), Antiphospholipid Ab (+) |
| P138 | Female | 10-15 | 45-50 | BD/RA/Sjogren's/Lupus/TRAPS-like | ANA (+), Anti-dsDNA(+), Anti-smooth muscle Ab(+) |
| P139 | Male | 10-15 | 20-25 | Autoimmune hepatitis type 2, ITP | CRP↑, ESR↑, C4↓, Anti-Liver cytosol 1 Ab(+), Platelets autoantibodies(+) |
| P140 | Female | 10-15 | 15-20 | Autoimmune hepatitis type 2, Inflammatory arthritis, ITP | WBC↑, CRP↑, ESR↑, IgG↑, ANA (+), Anti-Liver cytosol 1 Ab(+), Platelets autoantibodies(+) |

**Supplementary Table 2** **(continued).**

| Cases | Gender | Age range,  onset (+/- 5 years) | Age range,  diagnosis (+/- 5 years) | Main disease phenotype before diagnosis of HA20 | Laboratory findings related to inflammation and autoimmunity at diagnosis or during disease flare |
| --- | --- | --- | --- | --- | --- |
| P141 | Male | 5-10 | 10-15 | PFAPA-like, FMF-like, sJIA-like | CRP↑, ESR↑ |
| P142 | Female | 5-10 | 35-40 | BD-like, PFAPA-like | CRP↑, ESR↑,IgG↑, Anti-dsDNA(+), p-ANCA(+) |
| P143 | Female | 0-5 | 55-60 | BD-like, Inflammatory arthritis | CRP↑, FER↑, C3↓, C4↓, Anti-dsDNA(+), LAC(+) |
| P144 | Female | 5-10 | 30-35 | TRAPS-like, IBD-like | CRP↑ |
| P145 | Male | 0-5 | 5-10 | BD-like, Autoimmune hepatitis | ESR↑, Anti-Gliadin Ab (+), ANA (+), Anti-smooth muscle Ab(+) |
| P146 | NA | NA | NA | NA | NA |
| P147 | NA | NA | NA | NA | NA |
| P148 | NA | NA | NA | NA | NA |
| P149 | NA | NA | NA | NA | NA |
| P150 | NA | NA | NA | NA | NA |
| P151 | NA | NA | NA | NA | NA |
| P152 | Female | 5-10 | 10-15 | BD-like, SLE-like, PFAPA-like, Lymphoproliferation, B cell lymphoma | WBC↑, CRP↑, IgG↑, Anti-β2GP1 IgG(+), Anti-β2GP1 IgM(+), Platelets autoantibodies(+) |
| P153 | NA | NA | NA | Autoimmune hepatitis | NA |
| P154 | NA | NA | NA | NA | NA |
| P155 | Male | 0-5 | 0-5 | BD-like, IBD-like, PFAPA-like | WBC↑, CRP↑, ESR↑, IgE↑ |
| P156 | NA | NA | NA | NA | NA |
| P157 | NA | NA | NA | NA | NA |

**Supplementary Table 2** **(continued).**

| Cases | Gender | Age range,  onset (+/- 5 years) | Age range,  diagnosis (+/- 5 years) | Main disease phenotype before diagnosis of HA20 | Laboratory findings related to inflammation and autoimmunity at diagnosis or during disease flare |
| --- | --- | --- | --- | --- | --- |
| P158 | Female | 0-5 | 25-30 | BD-like, IBD-like, Neurologic disease | CRP↑, ESR↑, ANA (+), TPOAb(+), TGAb(+) |
| P159 | Female | 0-5 | 45-50 | BD-like, Recurrent infection, Recurrent liver inflammation | CRP↑, ESR↑, ANA (+), TPOAb(+), TGAb(+) |
| P160 | Female | 0-5 | 0-5 | Autoimmune hepatitis, BD-like | CRP↑, ESR↑, ANA (+) |
| P161 | Male | 0-5 | 0-5 | BD-like, Autoimmune hepatitis | IgE↑, ANA (+), Anti-centromere Ab (+), Anti-gliadin (+), Anti-Gliadin Ab(+) |
| P162 | Female | 5-10 | 55-60 | BD-like | CRP↑, ANA (+), Anti-Smooth muscle Ab(+) |
| P163 | Female | 0-5 | 5-10 | BD-like | WBC↑, ESR↑, CRP↑, ANA (+), Anti-centromere Ab(+), Gliadin Ab IgG(+), TTG(+) |
| P164 | Male | 0-5 | 35-40 | BD-like (oral ulceration) | CRP↑, FER↑, ANA (+), Anti-Smooth muscle Ab(+) |
| P165 | Female | 5-10 | 45-50 | Refractory RA, BD-like, Sensorineural hearing loss | WBC↑, ESR↑, CRP↑, IgG↑, IgE↑, ANA (+), Anti-CCP IgG(+), RF(+), C3↓, C4↓ |
| P166 | Female | 0-5 | 45-50 | BD-like, SLE-like, TRAPS-like, Sensorineural hearing loss, Autoimmune hepatitis | CRP↑, FER↑, C3↓, C4↓, ANA (+), LAC(+), IgG2↓ |
| P167 | Male | 15-20 | 15-20 | Sensorineural hearing loss, Vitiligo, Immunodeficiency | WBC↑, CRP↑, ESR↑, IgG2↓, IgG4↓, CD3+T↓ |

**Supplementary Table 2** **(continued).**

| Cases | Gender | Age range,  onset (+/- 5 years) | Age range,  diagnosis (+/- 5 years) | Main disease phenotype before diagnosis of HA20 | Laboratory findings related to inflammation and autoimmunity at diagnosis or during disease flare |
| --- | --- | --- | --- | --- | --- |
| P168 | Male | 0-5 | 10-15 | BD-like | ANA(±) |
| P169 | NA | NA | NA | NA | NA |
| P170 | Male | 15-20 | 45-50 | Autoimmune polyglandular syndrome type 2, CAPS-like, TRAPS-like, BD-like | WBC↑, CRP↑, C3↓, TPOAb(+), TGAb(+) |
| P171 | Male | 10-15 | 40-45 | Hashimoto's disease, TRAPS-like, BD-like, SLE-like | CRP↑, C3↓, TPOAb(+), TGAb(+) |
| P172 | Female | 0-5 | 15-20 | BD-like, IBD-like, SLE-like | NA |
| P173 | Male | 0-5 | NA | Autoimmune hepatitis, SLE-like, BD-like (oral ulceration), Microangiopathic hemolytic anemia, Pulmonary hypertension | WBC↑, CRP↑, ESR↑, FER↑, IgG↑, C3↓, C4↓, ANA (+), Anti-Smooth muscle Ab(+) |
| P174 | Male | NA | NA | BD-like | CRP↑ |
| P175 | Male | NA | NA | BD-like | IgE↑ |
| P176 | Male | 0-5 | 5-10 | Chronic nonbacterial osteomyelitis | WBC↑, CRP↑, ESR↑, FER↑ |
| P177 | Female | 0-5 | 65-70 | BD-like, Lymphoproliferation, Sensorineural hearing loss, Immunodeficiency, Diffuse large B-cell lymphoma | RF(+), ESR↑, FER↑, C4↓, IgG2↓, IgG4↓ |

**Supplementary Table 2** **(continued).**

| Cases | Gender | Age range,  onset (+/- 5 years) | Age range,  diagnosis (+/- 5 years) | Main disease phenotype before diagnosis of HA20 | Laboratory findings related to inflammation and autoimmunity at diagnosis or during disease flare |
| --- | --- | --- | --- | --- | --- |
| P178 | Male | 0-5 | 5-10 | Immunodeficiency, Autoimmune lymphoproliferative syndrome | CRP↑, IgE↑ |
| P179 | Male | 0-5 | NA | NA | NA |
| P180 | Male | 0-5 | 45-50 | BD-like oral ulceration, Genital ulcers, Polyarthritis, Nephritis | CRP↑, ESR↑, FER↑, C4↓, ANA (+), Anti-Smith(+) |
| P181 | Female | 0-5 | 5-10 | BD-like | Anti-smooth muscle Ab(+) |
| P182 | Female | 0-5 | 5-10 | BD-like, Aseptic meningitis, Arthritis, IBD-like, Hypothyroid without autoAb | CRP↑ |
| P183 | Female | 10-15 | 10-15 | BD-like, Pericarditis, IBD-like | NA |
| P184 | Male | 10-15 | 10-15 | IBD-like | WBC↑, CRP↑, ESR↑, FER↑, IgG↑ |
| P185 | Female | 0-5 | 15-20 | BD-like | C4↓, ANA (+) |

IBD: inflammatory bowel disease; PFAPA: periodic fever with aphthous pharyngitis and adenitis; BD: Behçet’s disease; sJIA: systemic juvenile idiopathic arthritis; SLE: systemic lupus erythematosus; ITP: immune thrombocytopenia; TRAPS: tumor necrosis factor receptor–associated periodic syndrome; RA: rheumatoid arthritis; WBC: white blood cells; ESR: erythrocyte sedimentation rate; CRP: C-reactive protein; SAA: serum amyloid A; ANA: antinuclear antibodies; TPOAb: anti-thyroid peroxidase antibody; TGAb: anti-thyroglobulin antibody; NA: not available.

**Supplementary Table 3 Comparison of clinical features and laboratory findings between two age groups.**

| General characteristics, n (%) | Age ＜ 16 years (n = 108) | Age ≥16 years (n = 60) | *P* value |
| --- | --- | --- | --- |
| Male gender, n (%) | 50 (47.2, n = 106) | 22 (36.7) | 0.3293 |
| Growth delay | 42 (39.3, n =107) | 5 (8.3) | 0.0001 |
| Recurrent fever | 71 (66.3, n =107) | 32 (53.3) | 0.099 |
| Lymphadenopathy | 36 (33.3) | 17 (28.8, n = 59) | 0.49 |
| Hepatomegaly | 28 (26.4, n = 106) | 7 (12.1, n = 58) | 0.03 |
| Splenomegaly | 22 (20.8, n = 106) | 12 (21.1, n = 57) | > 0.9999 |
| Recurrent infection | 40 (37.7, n = 106) | 16 (28.1, n =57) | 0.228 |
| Mucocutaneous involvement | 78 (73.6, n = 106) | 52 (91.2, n = 57) | 0.0124 |
| Rash | 44 (40.7) | 28 (46.7) | 0.626 |
| Oral ulcers | 68 (64.2) | 51 (85.0) | 0.004 |
| Genital ulcers | 22 (20.4) | 30 (50.0) | 0.002 |
| Gastrointestinal involvement | 67 (64.4, n = 104) | 29 (50, n = 58) | 0.0952 |
| Intestinal ulcers | 36 (35.0, n = 103) | 6 (11.3, n = 53) | 0.002 |
| Elevated levels of serum ALT/AST | 32 (30.2, n = 106) | 14 (24.1, n = 58) | 0.4697 |
| Cytopenias | 60 (56.1, n = 107) | 29 (50.9, n = 57) | 0.511 |
| Anemia | 51 (47.7, n = 107) | 18 (31.6, n = 57) | 0.0672 |
| Lymphopenia | 19 (17.6, n = 107) | 25 (43.9, n = 57) | 0.0005 |
| Thrombocytopenia | 14 (13.1, n = 107) | 13 (22.8, n = 57) | 0.1915 |
| Arthritis/arthralgia | 44 (41.5, n = 106) | 33 (55.9, n = 59) | 0.0742 |
| Thyroid involvement | 16 (15.5, n =103) | 13 (23.2, n = 56) | 0.2831 |
| Neurologic involvement | 17 (16.0, n = 106) | 10 (17.2, n = 58) | > 0.9999 |
| Pulmonary involvement | 21 (19.8, n = 106) | 12 (21.4, n = 56) | 0.8391 |
| Cardiac involvement | 8 (7.5, n = 106) | 6 (10.5, n = 57) | 0.5638 |
| Uveitis | 1 (0.96, n = 104) | 8 (14.3, n = 56) | 0.001 |

**Supplementary Table 3** **(continued).**

| General characteristics, n (%) | Age ＜ 16 years (n = 108) | Age ≥16 years (n = 60) | *P* value |
| --- | --- | --- | --- |
| Laboratory findings related  to inflammation |  |  |  |
| Leukocytosis | 47 (46.1, n = 102) | 10 (23.8, n = 42) | 0.015 |
| Elevated CRP level | 68 (67.3, n = 101) | 34 (81.0, n = 42) | 0.11 |
| Elevated ESR level | 69 (69.7, n = 99) | 29 (69.0, n = 42) | ＞0.9999 |
| Elevated SAA level | 18 (40.9, n = 44) | 2 (33.3, n = 6) | ＞0.9999 |
| Elevated ferritin level | 17 (23.0, n = 74) | 13 (38.2, n = 34) | 0.11 |
| Laboratory findings related  to autoimmunity |  |  |  |
| Elevated IgG level | 26 (26.5, n = 98) | 7 (17.9, n = 39) | 0.28 |
| Elevated IgE level | 31 (35.6, n = 87) | 4 (13.8, n = 29) | 0.03 |
| Decreased C3 level | 14 (15.1, n = 93) | 10 (25.0, n = 40) | 0.22 |
| Decreased C4 level | 10 (10.8, n = 93) | 11 (27.5, n = 40) | 0.02 |
| Positive Coombs' test | 14 (33.3, n = 42) | 1 (6.3, n = 16) | 0.046 |
| Positive TPOAb or TGAb | 10 (12.0, n = 83) | 7 (33.3, n = 21) | 0.026 |
| Positive ANA | 30 (30.3, n = 99) | 22 (53.7, n = 41) | 0.023 |
| Other autoantibodies | 26 (26.5, n = 98) | 25 (61.0, n = 41) | 0.0005 |

ALT: alanine transaminase; AST: aspartate transaminase; ESR: erythrocyte sedimentation rate; CRP: C-reactive protein; SAA: serum amyloid A; ANA: antinuclear antibodies; TPOAb: anti-thyroid peroxidase antibody; TGAb: anti-thyroglobulin antibody. If clearly labeled, the number of patients in total equals the number, or the sum of numbers, in parentheses. Comparisons stratified by age were performed using Mann-Whitney U test.

**Supplementary Table 4 Comparison of clinical manifestations in children between two cohorts.**

| Characteristics, n (%) | China (n = 83) | US (n = 23) | *P* value |
| --- | --- | --- | --- |
| Male gender, n (%) | 37 (44.6, n = 83) | 13 (54.2) | 0.3514 |
| Median age at onset (y) | 2 | 3 | 0.167 |
| Median age at diagnosis (y) | 3.9 | 9.4 | 0.0009 |
| Growth delay | 38 (45.8) | 5 (22.7, n = 22) | 0.477 |
| Recurrent fever | 55 (66.2) | 16 (69.6) | 0.6163 |
| Lymphadenopathy | 26 (31.3) | 9 (39.1) | 0.323 |
| Hepatomegaly | 21 (25.3) | 6 (28.6, n = 21) | 0.582 |
| Splenomegaly | 14 (16.9) | 7 (33.3, n = 21) | 0.042 |
| Recurrent infection | 32 (38.6) | 8 (34.8) | > 0.9999 |
| Respiratory tract infection | 16 (19.3) | 8 (34.8) | 0.06 |
| Mucocutaneous involvement | 58 (69.9) | 20 (87.0) | 0.056 |
| Rash | 31 (37.3) | 14 (60.9) | 0.013 |
| Oral ulcers | 46 (55.4) | 19 (82.6) | 0.007 |
| Genital ulcers | 13 (15.7) | 9 (39.1) | 0.007 |
| Perianal ulcers | 3 (3.6) | 3 (13.0) | 0.115 |
| Gastrointestinal involvement | 54 (67.5, n = 80) | 13 (56.5) | 0.6203 |
| Abdominal pain | 27 (34.2, n = 79) | 12 (52.2) | 0.0879 |
| Diarrhea | 34 (41.0) | 9 (39.1) | > 0.9999 |
| Esophageal ulcers | 3 (3.8, n = 80) | 1 (4.5, n = 22) | > 0.9999 |
| Gastric ulcers | 7 (8.8, n = 80) | 0 | 0.209 |
| Intestinal ulcers | 34 (43.0, n = 79) | 2 (9.1, n = 22) | 0.0047 |
| Hepatic involvement | 17 (20.5) | 13 (56.5, n = 22) | 0.0002 |
| Elevated levels of serum ALT/AST | 17 (20.5) | 13 (56.5) | 0.0002 |
| Autoimmune hepatitis | 8 (9.6) | 6 (27.3) | 0.0413 |
| Cytopenias | 43 (51.8) | 17 (77.3, n = 22) | 0.0144 |
| Anemia | 39 (47.0) | 12 (54.5) | 0.6333 |
| Neutropenia | 10 (12.0) | 3 (13.6) | > 0.9999 |
| Lymphopenia | 7 (8.4) | 10 (45.5) | 0.0002 |
| Thrombocytopenia | 10 (12.0) | 4 (18.1) | 0.4852 |
| Musculoskeletal system | 28 (33.7) | 16 (69.6) | 0.0008 |
| Arthritis/arthralgia | 28 (33.7) | 16 (69.6) | 0.0008 |
| Thyroid involvement | 10 (12.0) | 6 (27.3, n = 22) | 0.0972 |
| Hypothyroidism | 7 (8.4) | 2 (9.1) | > 0.9999 |
| Hashimoto disease | 9 (10.8) | 3 (13.6) | > 0.9999 |

**Supplementary Table 4** **(continued).**

| Characteristics, n (%) | China (n = 83) | US (n = 23) | *p*-value |
| --- | --- | --- | --- |
| Neurologic involvement | 10 (12.0) | 7 (30.4) | 0.0157 |
| Seizure | 7 (8.4) | 2 (9.1, n = 22) | > 0.9999 |
| Meningitis | 3 (3.6) | 1 (4.5, n = 22) | > 0.9999 |
| Developmental delay | 1 (1.2) | 6 (26.1) | < 0.0001 |
| Pulmonary involvement | 18 (21.7) | 3 (13.0) | 0.4008 |
| Pleural effusion | 6 (7.2) | 1 (4.3) | 0.6986 |
| Interstitial pneumonia | 8 (9.6) | 1 (4.3) | 0.6801 |
| Pulmonary nodules | 2 (2.4) | 1 (4.3) | > 0.9999 |
| Cardiac involvement | 5 (6.0) | 3 (13.0) | 0.3667 |
| Pericardial effusion | 3 (3.6) | 3 (13.0) | 0.1147 |
| Decreased ejection fraction | 2 (2.4) | 0 | > 0.9999 |
| Ophthalmologic involvement | 0 | 1 (4.3) | 0.2212 |
| Uveitis | 0 | 1 (4.3) | 0.2212 |

ALT: alanine transaminase; AST: aspartate transaminase. If clearly labeled, the number of patients in total equals the number, or the sum of numbers, in parentheses. Comparisons in children stratified by country of origin were performed using Mann-Whitney U test.

**Supplementary Table 5 Comparison of clinical manifestations between genders.**

| General characteristics, n (%) | Male (n = 78) | Female (n = 96) | *P* value |
| --- | --- | --- | --- |
| Median age at onset (y) | 3.3 | 3.1 | 0.8268 |
| Median age at diagnosis (y) | 9.9 | 11.8 | 0.4817 |
| Short stature | 26 (34.2, n = 76) | 24 (31.6, n =76) | > 0.9999 |
| Recurrent fever | 44 (57.9, n = 76) | 61 (66.3, n = 92) | 0.2609 |
| Lymphadenopathy | 26 (34.2, n = 76) | 27 (29.3, n = 92) | 0.6221 |
| Hepatomegaly | 16 (21.6, n = 74) | 20 (22.2, n = 90) | 0.8524 |
| Splenomegaly | 17 (23.0, n = 74) | 18 (20.2, n = 89) | 0.8509 |
| Recurrent infection | 27 (34.6) | 31 (34.8) | > 0.9999 |
| Respiratory tract infection | 16 (20.5) | 23 (24.0) | 0.4723 |
| Mucocutaneous involvement | 63 (80.8) | 72 (75.0) | 0.6935 |
| Rash | 28 (35.9) | 44 (45.8) | 0.1217 |
| Oral ulcers | 56 (71.8) | 64 (66.7) | 0.7311 |
| Genital ulcers | 14 (17.9) | 38 (39.6) | 0.001 |
| Perianal ulcers | 4 (5.1) | 4 (4.2) | > 0.9999 |
| Gastrointestinal involvement | 45 (61.6, n = 73) | 52 (60.5, n = 86) | > 0.9999 |
| Abdominal pain | 28 (38.3, n = 73) | 33 (36.3, n = 91) | > 0.9999 |
| Diarrhea | 25 (32.1) | 37 (38.5) | 0.3368 |
| Esophageal ulcers | 3 (4.2, n = 72) | 3 (3.6, n = 84) | > 0.9999 |
| Gastric ulcers | 6 (8.3, n = 72) | 4 (4.8, n = 84) | 0.5148 |
| Intestinal ulcers | 23 (32.9, n = 70) | 20 (23.8, n = 92) | 0.2791 |
| Hepatic involvement | 21 (26.9) | 25 (27.2 n = 92) | > 0.9999 |
| Elevated levels of serum ALT/AST | 20 (25.6) | 25 (27.2) | > 0.9999 |
| Autoimmune hepatitis | 10 (13.7) | 8 (8.7) | 0.3259 |
| Cytopenias | 44 (56.4) | 48 (53.9, n = 89) | 0.7543 |
| Anemia | 32 (41.0) | 38 (42.7) | > 0.9999 |
| Neutropenia | 13 (16.7) | 15 (16.9) | > 0.9999 |
| Lymphopenia | 23 (29.5) | 21 (23.6) | 0.3793 |
| Thrombocytopenia | 12 (15.4) | 15 (16.9) | 0.8358 |
| Musculoskeletal system | 37 (47.4) | 42 (43.8) | 0.8769 |
| Arthritis/arthralgia | 37 (47.4) | 41 (42.7) | 0.7569 |
| Thyroid involvement | 10 (13.5, n = 74) | 19 (21.1, n = 90) | 0.224 |
| Hypothyroidism | 7 (9.5) | 12(13.3) | 0.4738 |
| Hashimoto disease | 5 (6.8) | 10 (11.1) | 0.4198 |

**Supplementary Table 5** **(continued).**

| General characteristics, n (%) | Male (n = 78) | Female (n = 96) | *P* value |
| --- | --- | --- | --- |
| Neurologic involvement | 9 (11.5) | 18 (19.4, n = 93) | 0.1507 |
| Seizure | 2 (2.6) | 8 (8.6) | 0.1879 |
| Meningitis | 2 (2.6) | 5 (5.4, n = 92) | 0.4595 |
| Developmental delay | 5 (6.4) | 4 (4.3) | 0.755 |
| Pulmonary involvement | 14 (18.4, n = 76) | 20 (22.0, n = 91) | 0.4008 |
| Pleural effusion | 6 (7.9) | 6 (6.6) | 0.7719 |
| Interstitial pneumonia | 3 (3.9) | 8 (8.8) | 0.2323 |
| Pulmonary nodules | 5 (6.6) | 6 (6.6) | > 0.9999 |
| Cardiac involvement | 4 (5.3, n = 76) | 11 (12.0, n = 92) | 0.1759 |
| Pericardial effusion | 3 (3.9) | 9 (9.8) | 0.2284 |
| Decreased ejection fraction | 0 | 4 (4.3) | 0.1265 |
| Ophthalmologic involvement | 3 (4, n = 75) | 7 (7.8, n = 90) | 0.3496 |
| Uveitis | 3 (4) | 6 (6.7) | 0.5126 |

ALT: alanine transaminase; AST: aspartate transaminase. If clearly labeled, the number of patients in total equals the number, or the sum of numbers, in parentheses. Comparisons stratified by gender were performed using Mann-Whitney U test.

**Supplementary Table 6. Multiple linear regression analysis of the autoinflammation-predominant phenotype.**

| Parameter estimates | Variable | 95%CI | t value | *P* value |
| --- | --- | --- | --- | --- |
| β0 | Intercept | 0.1921 to 0.4771 | 4.637 | < 0.0001 |
| β1 | Age | -0.4004 to -0.04006 | 2.414 | 0.0169 |
| β2 | Country of origin | 0.06230 to 0.3809 | 2.747 | 0.0067 |

Results from multivariable linear regression analysis are shown. β: regression coefficients; CI: confidence interval;

**Supplementary Table 7 Genetic analysis of HA20 patients.**

| Cases | cDNA change | Amino acid Substitution | Variant class | *Inheritance* |
| --- | --- | --- | --- | --- |
| P1 | c.610A>T | p.R204* | Novel | *De novo* |
| P2 | del exon 6-8 | － | Novel | *De novo* |
| P3 | c.732_733del | p.Y244* | Novel | *De novo* |
| P4 | c.910_914del | p.K304Vfs*27 | Reported | *De novo* |
| P5 | c.1690A>T | p.K564* | Reported | Maternally |
| P6 (P5's mother) | c.1690A>T | p.K564* | Reported | Unknown |
| P7 | c.2184_2190del | p.E730Sfs*84 | Reported | Paternally |
| P8 (P7's father) | c.2184_2190del | p.E730Sfs*84 | Reported | Unknown |
| P9 | c.258_261del | p.C86Wfs*9 | Reported | *De novo* |
| P10 | c.1247_1251del | p.N416Tfs*11 | Novel | *De novo* |
| P11 | del exon 5-8 | － | Novel | *De novo* |
| P12 | c.334del | p.M112Cfs*11 | Novel | *De novo* |
| P13 | c.1315_1316insA | p.R439Qfs*6 | Reported | *De novo* |
| P14 | c.361del | p.V121Yfs*2 | Novel | *De novo* |
| P15 | c.569C>G | p.S190* | Novel | *De novo* |
| P16 (P17's mother) | c.811C>T | p.R271* | Reported | Maternally |
| P17 | c.811C>T | p.R271* | Reported | Unknown |
| P18 | c.634+2T>C | p.D212Gfs*38 | Reported | Maternally |
| P19 (P18's son) | c.634+2T>C | p.D212Gfs*38 | Reported | Paternally |
| P20 (P18's mother) | c.634+2T>C | p.D212Gfs*38 | Reported | Unknown |
| P21 | c.259C>T | p.R87* | Reported | *De novo* |

**Supplementary Table 7** **(continued).**

| Cases | cDNA change | Amino acid Substitution | Variant class | *Inheritance* |
| --- | --- | --- | --- | --- |
| P22 | c.1906+2T>G | － | Reported | Maternally |
| P23 (P22's mother) | c.1906+2T>G | － | Reported | Unknown |
| P24 | c.857T>G | p.L286* | Novel | Maternally |
| P25 | c.2274dup | p.K759Qfs*10 | Reported | *De novo* |
| P26 | c.1108del | p.Q370Rfs*16 | Reported | *De novo* |
| P27 (P24's mother) | c.857T>G | p.L286* | Novel | Unknown |
| P28 | c.440_441del | p.L147Qfs*7 | Reported | *De novo* |
| P29 | del exon 2-9 | － | Novel | *De novo* |
| P30 | c.1300_1301delinsTA | p.A434* | Reported | *De novo* |
| P31 | c.1129_1133del | p.V377Pfs*10 | Novel | *De novo* |
| P32 | c.429_433delinsTTTA | p.Q143Hfs*73 | Novel | *De novo* |
| P33 | c.547C>T | p.R183* | Reported | Paternally |
| P34 | c.133C>T | p.R45* | Reported | *De novo* |
| P35 | c.811C>T | p.R271* | Reported | *De novo* |
| P36 | c.887dup | p.N296Kfs*2 | Novel | *De novo* |
| P37 | del exon 7-8 | － | Reported | *De novo* |
| P38 (P45's daughter) | c.559C>T | p.Q187* | Reported | Paternally |

**Supplementary Table 7** **(continued).**

| Cases | cDNA change | Amino acid Substitution | Variant class | *Inheritance* |
| --- | --- | --- | --- | --- |
| P39 (P45's son) | c.559C>T | p.Q187* | Reported | Paternally |
| P40 (P46's daughter) | c.1760dup | p.A588Cfs*84 | Reported | Maternally |
| P41 (P46's son) | c.1760dup | p.A588Cfs*84 | Reported | Maternally |
| P42 | c.721C>T | p.Q241* | Novel | *De novo* |
| P43 | c.259C>T | p.R87* | Reported | *De novo* |
| P44 | Del 6q23.3-6q24.1 | － | Novel | *De novo* |
| P45 | c.559C>T | p.Q187* | Reported | Unknown |
| P46 | c.1760dup | p.A588Cfs*84 | Reported | Paternally |
| P47 (P36's father) | c.887dup | p.N296Kfs*2 | Novel | Unknown |
| P48 | c.1820_1821GCdelinsAA | p.C607* | Novel | *De novo* |
| P49 | c.1805del | p.T602Rfs*95 | Novel | Maternally |
| P50 (P49's mother) | c.1805del | p.T602Rfs*95 | Novel | Unknown |
| P51 | c.1247_1251del | p.N416Tfs*11 | Novel | Paternally |
| P52 | c.811C>T | p.R271* | Reported | *De novo* |
| P53 | c.1906+1G>A | － | Reported | *De novo* |
| P54 | c.811C>T | p.R271* | Reported | Maternally |
| P55(P54's mother) | c.811C>T | p.R271* | Reported | Unknown |
| P56 | c.1901del | p.N634Tfs*63 | Novel | *De novo* |
| P57 | c.133C>T | p.R45* | Reported | *De novo* |
| P58 (P33's father) | c.547C>T | p.R183* | Reported | Unknown |
| P59 (P51’s father) | c.1247_1251del | p.N416Tfs*11 | Novel | Unknown |

**Supplementary Table 7** **(continued).**

| Cases | cDNA change | Amino acid Substitution | Variant class | *Inheritance* |
| --- | --- | --- | --- | --- |
| P60 | c.971_975del | p.L324Qfs*7 | Reported | *De novo* |
| P61 | c.1135C>T | p.Q379* | Reported | *De novo* |
| P62 | c.866del | p.H289Pfs*3 | Novel | *De novo* |
| P63 | c.1906+2T>G | － | Reported | Unknown |
| P64 | c.811C>T | p.R271* | Reported | Unknown |
| P65 | c.1877dup | p.C627Vfs*45 | Novel | Maternally |
| P66 (P65's mother) | c.1877dup | p.C627Vfs*45 | Novel | Unknown |
| P67 | c.2274dup | p.K759Qfs*10 | Reported | Paternally |
| P68 (P67's brother) | c.2274dup | p.K759Qfs*10 | Reported | Paternally |
| P69 (P67's father) | c.2274dup | p.K759Qfs*10 | Reported | Unknown |
| P70 | c.1681C>T | p.Q561* | Novel | Unknown |
| P71 (P63's daughter) | c.1906+2T>G | － | Reported | Maternally |
| P72 (P64's daughter) | c.811C>T | p.R271* | Reported | Maternally |
| P73 | c.547C>T | p.R183* | Reported | Paternally |
| P74 (P73's father) | c.547C>T | p.R183* | Reported | Unknown |
| P75 | c.2274dup | p.K759Qfs*10 | Reported | *De novo* |
| P76 | c.866del | p.H289Pfs*3 | Novel | *De novo* |
| P77 | del(6)q23.3 | － | Novel | *De novo* |
| P78 | c.1904_1907del | p.K635Ifs*61 | Novel | *De novo* |
| P79 | c.133C>T | p.R45* | Reported | Maternally |

**Supplementary Table 7** **(continued).**

| Cases | cDNA change | Amino acid Substitution | Variant class | *Inheritance* |
| --- | --- | --- | --- | --- |
| P80 | c.133C>T | p.R45* | Reported | Unknown |
| P81 | c.133C>T | p.R45* | Reported | *De novo* |
| P82 | c.738C>A | P.Y246* | Reported | *De novo* |
| P83 | c.259C>T | p.R87* | Reported | *De novo* |
| P84 | c.133C>T | p.R45* | Reported | *De novo* |
| P85 | c.1840_1841insGA | p.Y614* | Novel | Unknown |
| P86 | c.2023C>T | p.Q675* | Novel | Unknown |
| P87 | c.1812del | p.S605Afs*92 | Novel | Paternally |
| P88 (P87's father) | c.1812del | p.S605Afs*92 | Novel | Unknown |
| P89 (P46's mother) | c.1760dup | p.A588Cfs*84 | Reported | Unknown |
| P90 | c.2024_2025insTGTTTGAAG | p.Q675_G790delinsHV | Novel | *De novo* |
| P91 | c.604del | p.I202Sfs*14 | Novel | *De novo* |
| P92 | c.438del | p.L147Sfs*69 | Novel | *De novo* |
| P93 | c.292_295dup | p.G99Efs*3 | Novel | Maternally |
| P94 (P93's mother) | c.292_295dup | p.G99Efs*3 | Novel | Unknown |
| P95 | c.1812dup | p.S605Qfs*67 | Novel | *De novo* |
| P96 | c.653del | p.L218Wfs*10 | Novel | Paternally |
| P97 (P96's father) | c.653del | p.L218Wfs*10 | Novel | Unknown |
| P98 | c.2217del | p.N740Tfs*76 | Novel | *De novo* |
| P99 | c.971_975del | p.L324Qfs*7 | Reported | Paternally |
| P100 (P99's father) | c.971_975del | p.L324Qfs*7 | Reported | Unknown |
| P101 | c.220C>T | p.Q74* | Novel | *De novo* |

**Supplementary Table 7** **(continued).**

| Cases | cDNA change | Amino acid Substitution | Variant class | *Inheritance* |
| --- | --- | --- | --- | --- |
| P102 | c.259C>T | p.R87* | Reported | *De novo* |
| P103 | c.811C>T | p.R271* | Reported | Unknown |
| P104 | c.259C>T | p.R87* | Reported | *De novo* |
| P105 | c.1067G>A | p.Trp356* | Reported | Unknown |
| P106 | c.1976_1997dup | p.S667Efs*12 | Novel | Unknown |
| P107 | c.547C>T | p.R183* | Reported | *De novo* |
| P108 | c.1516del | p.A506Pfs*191 | Novel | Paternally |
| P109 | c.971_975del | p.L324Qfs*7 | Reported | *De novo* |
| P110 | c.2256_2259del | p.A753Pfs*62 | Novel | *De novo* |
| P111 (P112's father) | c.1927A>T | p.K643* | Novel | Maternally |
| P112 | c.1927A>T | p.K643* | Novel | Paternally |
| P113 (P112's aunt) | c.1927A>T | p.K643* | Novel | Unknown |
| P114 (P112's grandma) | c.1927A>T | p.K643* | Novel | Paternally |
| P115 (P112's sister) | c.1927A>T | p.K643* | Novel | Maternally |
| P116 (P112's nephew) | c.1927A>T | p.K643* | Novel | Maternally |
| P117 | c.1876_1877del | p.L626Vfs*45 | Reported | *De novo* |
| P118 | c.503G>A | p.W168* | Reported | Unknown |
| P119 | c.671del | p.F224Sfs*4 | Reported | Unknown |
| P120 | c.671del | p.F224Sfs*4 | Reported | Unknown |
| P121 | c.671del | p.F224Sfs*4 | Reported | Maternally |

**Supplementary Table 7** **(continued).**

| Cases | cDNA change | Amino acid Substitution | Variant class | *Inheritance* |
| --- | --- | --- | --- | --- |
| P122 | Deletion of entire coding sequence (and surrounding genes) | - | Reported | Unknown |
| P123 | c.1809del | p.T604Rfs*93 | Reported | *De novo* |
| P124 | c.680T>A | p.L227* | Reported | Maternally |
| P125 | c.680T>A | p.L227* | Reported | Maternally |
| P126 | c.680T>A | p.L227* | Reported | Unknown |
| P127 | c.120del | p.F40Lfs*56 | Reported | De novo |
| P128 | c.811del | p.R27Efs*16 | Novel | De novo |
| P129 | c.503G>A | p.W168* | Reported | Paternally |
| P130 | c.505_544del | p.D169Pfs*34 | Novel | Paternally |
| P131 | c.1904_1905del | p.K635Tfs*36 | Reported | Maternally |
| P132 | c.1904_1905del | p.K635Tfs*36 | Reported | Maternally |
| P133 | c.1904_1905del | p.K635Tfs*36 | Reported | Unknown |
| P134 | c.811C>T | p.R271* | Reported | Paternally |
| P135 | c.811C>T | p.R271* | Reported | Paternally |
| P136 | c.811C>T | p.R271* | Reported | Paternally |
| P137 | c.811C>T | p.R271* | Reported | Paternally |
| P138 | c.811C>T | p.R271* | Reported | Paternally |
| P139 | Deletion of entire coding sequence | - | Reported | Maternally |
| P140 | Deletion of entire coding sequence | - | Reported | Maternally |
| P141 | Deletion of entire coding sequence | - | Reported | Maternally |
| P142 | Deletion of entire coding sequence | - | Reported | Unknown |

**Supplementary Table 7** **(continued).**

| Cases | cDNA change | Amino acid Substitution | Variant class | *Inheritance* |
| --- | --- | --- | --- | --- |
| P143 | c.671del | p.F224Sfs*4 | Reported | Unknown |
| P144 | c.235del | p.S79Afs*17 | Reported | Maternally |
| P145 | c.235del | p.S79Afs*17 | Reported | Maternally |
| P146 | c.2093C>A | p.S698* | Novel | Unknown |
| P147 | c.805+1G>T | - | Novel | *De novo* |
| P148 | c.226dup | p.T76Nfs*25 | Reported | Maternally |
| P149 | c.226dup | p.T76Nfs*25 | Reported | Maternally |
| P150 | c.226dup | p.T76Nfs*25 | Reported | Unknown |
| P151 | c.226dup | p.T76Nfs*25 | Reported | Unknown |
| P152 | Deletion of entire coding sequence (and surrounding genes) | - | Reported | *De novo* |
| P153 | c.475del | p.Y159Mfs*57 | Novel | Unknown |
| P154 | c.950G>A | p.W317* | Reported | Unknown |
| P155 | c.640dup | p.M214Nfs*40 | Reported | *De novo* |
| P156 | c.918C>G | p.Y306* | Reported | Unknown |
| P157 | c.1484_1493dup | p.C498Wfs*2 | Novel | Unknown |
| P158 | c.468dup | p.L157Afs*4 | Reported | Maternally |
| P159 | c.468dup | p.L157Afs*4 | Reported | Unknown |
| P160 | c.468dup | p.L157Afs*4 | Reported | Maternally |
| P161 | c.235del | p.S79Afs*17 | Reported | Maternally |
| P162 | c.235del | p.S79Afs*17 | Reported | Paternally |

**Supplementary Table 7** **(continued).**

| Cases | cDNA change | Amino acid Substitution | Variant class | *Inheritance* |
| --- | --- | --- | --- | --- |
| P163 | c.235del | p.S79Afs*17 | Reported | Paternally |
| P164 | c.235del | p.S79Afs*17 | Reported | Maternally |
| P165 | c.2217_2242del | p.N740Pfs*3 | Reported | Maternally |
| P166 | c.2217_2242del | p.N740Pfs*3 | Reported | Maternally |
| P167 | c.2217_2242del | p.N740Pfs*3 | Reported | Maternally |
| P168 | c.468dup | p.L157Afs*4 | Reported | Maternally |
| P169 | c.1694C>G | p.S565* | Reported | Unknown |
| P170 | c.547C>T | p.R183* | Reported | Unknown |
| P171 | c.547C>T | p.R183* | Reported | Unknown |
| P172 | c.547C>T | p.R183* | Reported | Paternally |
| P173 | c.232dup | p.E78Gfs*23 | Novel | Paternally |
| P174 | c.232dup | p.E78Gfs*23 | Novel | *De novo* |
| P175 | c.232dup | p.E78Gfs*23 | Novel | Paternally |
| P176 | c.971_975del | p.L324Qfs*7 | Reported | Maternally |
| P177 | c.2217_2242del | p.N740Pfs*3 | Reported | Unknown |
| P178 | Deletion in heterozygosity chr6:135286410-145172140 | - | Novel | Unknown |
| P179 | c.505_544del | p.D169Pfs*34 | Novel | Maternally |

**Supplementary Table 7** **(continued).**

| Cases | cDNA change | Amino acid Substitution | Variant class | *Inheritance* |
| --- | --- | --- | --- | --- |
| P180 | c.1068G>A | p.W356* | Reported | *Unknown* |
| P181 | c.1068G>A | p.W356* | Reported | *Paternally* |
| P182 | TNFAIP3 del (whole genome microarray revealed a loss of 6q23.3q24 region of the long arm of chromosome 6 spanning approximately 1.782 Mb and encompassing at least 20 genes (OLIG3, WAKMAR2, TNFAIP3, PERP, ARFGEF3, PBOV1, HEBP2, CCDC28A, REPS1, HECA, NHSL1, ECT2L, GVQW2, ABRACL)) | - | Reported | *De novo* |
| P183 | c.503G>A | p.W168* | Reported | Paternally |
| P184 | c.811C>T | p.R271* | Reported | Maternally |
| P185 | Deletion of entire coding sequence | - |  | Unknown |

*De novo*: pathogenic variants not detected in either healthy parent; Unknown inheritance: pathogenic variants could not be confirmed due to inability to obtain DNA samples from one or both parents; Paternally: the same pathogenic variant was confirmed by genetic testing in the patient’s father; Maternally: the same pathogenic variant was confirmed by genetic testing in the patient’s mother.

**Supplementary Table 8 Treatment and clinical response of HA20 patients.**

| Cases | Treatment before genetic diagnosis | Initial treatment post-genetic diagnosis | Treatment at maximal response | Maximal response | Treatment at the most recent follow up | Clinical response at the most recent follow up |
| --- | --- | --- | --- | --- | --- | --- |
| P1 | Prednisone, levothyroxine | Prednisone, azathioprine, HCQ, levothyroxine | Prednisone (1.25mg/d), azathioprine, HCQ, levothyroxine | Minimal disease activity | Azathioprine, HCQ, levothyroxine | Minimal disease activity |
| P2 | Prednisone, MTX, antibiotics | Prednisone, infliximab, NSAIDs | Prednisone (1.25mg/d),  adalimumab, colchicine, tofacitinib | Minimal disease activity | Adalimumab→etanercept, colchicine, tofacitinib | Partial response |
| P3 | Prednisone, MTX, antibiotics | Prednisone, MTX, adalimumab | Prednisone (1.25mg/d), adalimumab, thalidomide/colchicine | Minimal disease activity | Prednisone, adalimumab→etanercept→infliximab, colchicine | Partial response |
| P4 | Prednisone, antibiotics | Prednisone, MTX, etanercept, NSAIDs | Prednisone, MTX, etanercept | Minimal disease activity | Prednisone, etanercept | Minimal disease activity |
| P5 | Antibiotics, IVIG | Prednisone | Prednisone, thalidomide, colchicine | Minimal disease activity | Thalidomide, colchicine | Partial response |

**Supplementary Table 8** **(continued).**

| Cases | Treatment before genetic diagnosis | Initial treatment post-genetic diagnosis | Treatment at maximal response | Maximal response | Treatment at the most recent follow up | Clinical response at the most recent follow up |
| --- | --- | --- | --- | --- | --- | --- |
| P6 (P5's mother) | Prednisone, thalidomide | Prednisone, infliximab | PSL→ prednisone + CTX →Prednisone + azathioprine + thalidomide | Minimal disease activity | Prednisone, azathioprine | Minimal disease activity |
| P7 | Prednisone | Prednisone, thalidomide, infliximab | Prednisone, thalidomide, azathioprine | Minimal disease activity | Thalidomide, azathioprine | Minimal disease activity |
| P8  (P7's father) | Prednisone, antibiotics | Prednisone, thalidomide, NSAIDs | Prednisone, thalidomide | Minimal disease activity | Prednisone, thalidomide | Minimal disease activity |
| P9 | Prednisone, MTX | Prednisone, CTX → MMF | Prednisone, MMF, etanercept | Partial response | Prednisone, MMF | Partial response |
| P10 | Antibiotics, Vitamin B12 | Thalidomide, Vitamin B12 | Thalidomide, Vitamin B12 | Minimal disease activity | Thalidomide, Vitamin B12 | Minimal disease activity |
| P11 | Prednisone, antibiotics | Prednisone | Prednisone | Partial response | NA | NA |
| P12 | Prednisone, MTX | Prednisone, infliximab | Prednisone, infliximab | Partial response | Prednisone, infliximab | Partial response |
| P13 | Prednisone, MTX | Prednisone, MTX, infliximab | Thalidomide, mesalazine | Partial response | Thalidomide, mesalazine, tofacitinib | Partial response |

**Supplementary Table 8 (continued).**

| Cases | Treatment before genetic diagnosis | Initial treatment post-genetic diagnosis | Treatment at maximal response | Maximal response | Treatment at the most recent follow up | Clinical response at the most recent follow up |
| --- | --- | --- | --- | --- | --- | --- |
| P14 | Prednisone | Prednisone, mesalazine | Infliximab | Minimal disease activity | Infliximab | Minimal disease activity |
| P15 | Antibiotics | NA | NA | NA | NA | NA |
| P16  (P17's mother) | Prednisone, antibiotics | Prednisone, MMF, HCQ | Prednisone, MMF, tacrolimus | Minimal disease activity | Prednisone, MMF | Minimal disease activity |
| P17 | ─ | ─ | ─ | ─ | ─ | ─ |
| P18 | Prednisone | Prednisone, MMF | Prednisone, HCQ, tacrolimus | Minimal disease activity | Prednisone, HCQ, tacrolimus | Minimal disease activity |
| P19  (P18's son) | Prednisone | NA | NA | NA | NA | NA |
| P20  (P18's mother) | NA | NA | NA | NA | NA | NA |
| P21 | Antibiotics | Prednisone, adalimumab, tofacitinib | Prednisone, adalimumab, tofacitinib | Minimal disease activity | Prednisone, adalimumab, tofacitinib | Minimal disease activity |

**Supplementary Table 8 (continued).**

| Cases | Treatment before genetic diagnosis | Initial treatment post-genetic diagnosis | Treatment at maximal response | Maximal response | Treatment at the most recent follow up | Clinical response at the most recent follow up |
| --- | --- | --- | --- | --- | --- | --- |
| P22 | Prednisone, antibiotics | Prednisone, adalimumab, tofacitinib | Prednisone, adalimumab, tofacitinib | Minimal disease activity | Adalimumab | Minimal disease activity |
| P23  (P22's mother) | ─ | Thalidomide | Thalidomide | Minimal disease activity | Drug withdrawal | Minimal disease activity |
| P24 | Antibiotics, mesalazine | Mesalazine | Mesalazine | Minimal disease activity | Mesalazine | Minimal disease activity |
| P25 | NA | NA | NA | NA | NA | NA |
| P26 | Antibiotics | Prednisone, mesalazine, infliximab | Prednisone, mesalazine, IVIG | Partial response | Prednisone, adalimumab, tofacitinib, MTX | Partial response |
| P27  (P24's mother) | NA | NA | NA | NA | NA | NA |
| P28 | Prednisone | Prednisone, etanercept | Prednisone, etanercept, salazosulfadimidine | Minimal disease activity | Prednisone, etanercept, salazosulfadimidine | Partial response |

**Supplementary Table 8** **(continued).**

| Cases | Treatment before genetic diagnosis | Initial treatment post-genetic diagnosis | Treatment at maximal response | Maximal response | Treatment at the most recent follow up | Clinical response at the most recent follow up |
| --- | --- | --- | --- | --- | --- | --- |
| P29 | Prednisone, antibiotics | Prednisone, celecoxib, infliximab →Prednisone, CTX, thalidomide | Prednisone, colchicine, thalidomide | Partial response | Prednisone, MMF, leflunomide | Partial response |
| P30 | Dexamethasone | Prednisone, HCQ, tofacitinib, adalimumab | Prednisone, HCQ,colchicine, belimumab | Minimal disease activity | Prednisone, HCQ, colchicine, belimumab | Minimal disease activity |
| P31 | Antibiotics | Prednisone, etanercept, MTX, tofacitinib | Prednisone, colchicine, tofacitinib | Minimal disease activity | Prednisone, colchicine, tofacitinib | Minimal disease activity |
| P32 | Prednisone, antibiotics | PSL → prednisone, IVIG, tofacitinib | Prednisone, tofacitinib, belimumab | Minimal disease activity | Prednisone, tofacitinib, belimumab | Minimal disease activity |
| P33 | Antibiotics, etanercept, thalidomide, MTX | Prednisone, thalidomide, HCQ, tocilizumab | Prednisone, thalidomide, HCQ, tocilizumab | Minimal disease activity | Prednisone, thalidomide, HCQ, tocilizumab | Minimal disease activity |
| P34 | ─ | Prednisone, Cyclosporine | NA | NA | NA | NA |

**Supplementary Table 8 (continued).**

| Cases | Treatment before genetic diagnosis | Initial treatment post-genetic diagnosis | Treatment at maximal response | Maximal response | Treatment at the most recent follow up | Clinical response at the most recent follow up |
| --- | --- | --- | --- | --- | --- | --- |
| P35 | HCQ, levothyroxine | Thalidomide, HCQ, levothyroxine | Thalidomide, levothyroxine | Minimal disease activity | Thalidomide, levothyroxine | Minimal disease activity |
| P36 | Antibiotics, IVIG | Prednisone, thalidomide | Prednisone, thalidomide | Minimal disease activity | Thalidomide | Minimal disease activity |
| P37 | Prednisone, antibiotics | PSL → prednisone, Cyclosporine | Prednisone, HCQ, belimumab, Cyclosporine | Minimal disease activity | Prednisone, HCQ, belimumab | Minimal disease activity |
| P38  (P45's daughter) | Levothyroxine | Prednisone, etanercept, HCQ, MMF, levothyroxine | Prednisone, etanercept, HCQ, MMF, levothyroxine | Partial response | Prednisone, etanercept, HCQ, MMF, levothyroxine | Partial response |
| P39  (P45's son) | ─ | Etanercept, MTX | Etanercept, MTX | Minimal disease activity | Etanercept, MTX | Minimal disease activity |
| P40 (P46's daughter) | ─ | Prednisone, HCQ, MMF | Prednisone, etanercept, HCQ, MMF | Minimal disease activity | Prednisone, etanercept, HCQ, MMF | Minimal disease activity |

**Supplementary Table 8** **(continued).**

| Cases | Treatment before genetic diagnosis | Initial treatment post-genetic diagnosis | Treatment at maximal response | Maximal response | Treatment at the most recent follow up | Clinical response at the most recent follow up |
| --- | --- | --- | --- | --- | --- | --- |
| P41 (P46's son) | ─ | Etanercept, HCQ | Etanercept, HCQ | Minimal disease activity | Etanercept, HCQ | Minimal disease activity |
| P42 | Antibiotics | Prednisone, etanercept | Prednisone, etanercept | Minimal disease activity | Etanercept, thalidomide | Minimal disease activity |
| P43 | Prednisone, antibiotics, Cyclosporine | Prednisone, etanercept | Prednisone, etanercept | Minimal disease activity | Prednisone, etanercept | Disease flare |
| P44 | Prednisone | Prednisone, HCQ, CTX →thalidomide | Prednisone, HCQ, thalidomide | Minimal disease activity | Prednisone, HCQ, thalidomide | Minimal disease activity |
| P45 | NA | NA | NA | NA | NA | NA |
| P46 | NA | NA | NA | NA | NA | NA |
| P47 (P36's father) | Prednisone | Prednisone | Prednisone | Minimal disease activity | Drug withdrawal | Minimal disease activity |

**Supplementary Table 8 (continued).**

| Cases | Treatment before genetic diagnosis | Initial treatment post-genetic diagnosis | Treatment at maximal response | Maximal response | Treatment at the most recent follow up | Clinical response at the most recent follow up |
| --- | --- | --- | --- | --- | --- | --- |
| P48 | Adalimumab, mesalazine | Adalimumab, mesalazine | Adalimumab, mesalazine | Minimal disease activity | Adalimumab, mesalazine | Minimal disease activity |
| P49 | Antibiotics, thalidomide, methimazole | Thalidomide, methimazole | Thalidomide, methimazole | Minimal disease activity | Thalidomide, methimazole | Minimal disease activity |
| P50 (P49's mother) | NA | NA | NA | NA | NA | NA |
| P51 | Prednisone, antibiotics | Prednisone, colchicine, HCQ, azathioprine | Prednisone, colchicine, HCQ, azathioprine | Minimal disease activity | Prednisone, adalimumab, thalidomide | Minimal disease activity |
| P52 | Prednisone, antibiotics | Prednisone, MTX, infliximab | Prednisone, MTX, infliximab | Minimal disease activity | MTX, infliximab | Minimal disease activity |
| P53 | Thalidomide | Thalidomide, MTX → etanercept | Thalidomide, adalimumab | Minimal disease activity | Thalidomide, adalimumab | Minimal disease activity |
| P54 | Antibiotics | Adalimumab, MTX | Adalimumab, MTX | Minimal disease activity | Thalidomide, adalimumab | Partial response |

**Supplementary Table 8 (continued).**

| Cases | Treatment before genetic diagnosis | Initial treatment post-genetic diagnosis | Treatment at maximal response | Maximal response | Treatment at the most recent follow up | Clinical response at the most recent follow up |
| --- | --- | --- | --- | --- | --- | --- |
| P55 (P54's mother) | Thalidomide | Drug withdrawal | Drug withdrawal | NA | Drug withdrawal | NA |
| P56 | Prednisone, antibiotics, levothyroxine, CTX | PSL→ prednisone, HCQ, MMF | PSL→ prednisone, HCQ, CTX | Partial response | prednisone, HCQ, MMF | Partial response |
| P57 | Antibiotics | Thalidomide | Thalidomide | Minimal disease activity | Thalidomide | Minimal disease activity |
| P58 (P33's father) | Thalidomide | Thalidomide | Thalidomide | Partial response | Thalidomide | Partial response |
| P59 (P51’s father) | NA | NA | NA | NA | NA | NA |
| P60 | Prednisone, antibiotics | Prednisone, levothyroxine | NA | NA | NA | NA |
| P61 | Prednisone, infliximab | Prednisone, infliximab, azathioprine | Adalimumab | Minimal disease activity | Adalimumab | Partial response |
| P62 | Prednisone, infliximab, thalidomide | Prednisone, adalimumab | Adalimumab | Minimal disease activity | Prednisone, adalimumab | Partial response |

**Supplementary Table 8 (continued).**

| Cases | Treatment before genetic diagnosis | Initial treatment post-genetic diagnosis | Treatment at maximal response | Maximal response | Treatment at the most recent follow up | Clinical response at the most recent follow up |
| --- | --- | --- | --- | --- | --- | --- |
| P63 | ─ | Thalidomide | Thalidomide | Minimal disease activity | Thalidomide | Minimal disease activity |
| P64 | Prednisone, infliximab, sulfasalazine | Prednisone, infliximab, sulfasalazine | NA | NA | NA | NA |
| P65 | Prednisone, antibiotics | Prednisone, thalidomide | NA | NA | NA | NA |
| P66 (P65's mother) | NA | NA | NA | NA | NA | NA |
| P67 | Prednisone, colchicine, thalidomide, IVIG | Prednisone, colchicine, thalidomide, MTX, etanercept | Thalidomide, MTX, etanercept | Minimal disease activity | Thalidomide, MTX, etanercept | Minimal disease activity |
| P68  (P67's brother) | ─ | Etanercept | Etanercept | Minimal disease activity | Etanercept | Minimal disease activity |
| P69  (P67's father) | Colchicine | Colchicine, etanercept | Colchicine, etanercept | NA | Etanercept, MTX | NA |
| P70 | NA | NA | NA | NA | NA | NA |

**Supplementary Table 8** **(continued).**

| Cases | Treatment before genetic diagnosis | Initial treatment post-genetic diagnosis | Treatment at maximal response | Maximal response | Treatment at the most recent follow up | Clinical response at the most recent follow up |
| --- | --- | --- | --- | --- | --- | --- |
| P71 (P63's daughter) | NA | NA | NA | NA | NA | NA |
| P72 (P64's daughter) | NA | NA | NA | NA | NA | NA |
| P73 | Prednisone, CTX → MMF | Prednisone, CTX → MMF | Prednisone, CTX → MMF, IVIG | Minimal disease activity | Prednisone, MMF | Minimal disease activity |
| P74 (P73's father) | NA | NA | NA | NA | NA | NA |
| P75 | NA | NA | NA | NA | NA | NA |
| P76 | Prednisone, antibiotics, thalidomide | Adalimumab → infliximab | NA | Partial response | NA | NA |
| P77 | Prednisone, antibiotics, thalidomide | Prednisone, thalidomide | Thalidomide | Minimal disease activity | Thalidomide | Minimal disease activity |
| P78 | Antibiotics, thalidomide | Thalidomide | Thalidomide | Minimal disease activity | Thalidomide | Minimal disease activity |
| P79 | Antibiotics | Prednisone | Thalidomide | Minimal disease activity | Thalidomide | Minimal disease activity |

**Supplementary Table 8 (continued).**

| Cases | Treatment before genetic diagnosis | Initial treatment post-genetic diagnosis | Treatment at maximal response | Maximal response | Treatment at the most recent follow up | Clinical response at the most recent follow up |
| --- | --- | --- | --- | --- | --- | --- |
| P80 | NA | NA | NA | NA | NA | NA |
| P81 | Antibiotics | Adalimumab | Adalimumab | Minimal disease activity | Adalimumab | Minimal disease activity |
| P82 | Prednisone | Prednisone, colchicine | Prednisone | Minimal disease activity | Prednisone | Minimal disease activity |
| P83 | Prednisone, antibiotics | Prednisone, thalidomide | Thalidomide | Minimal disease activity | Thalidomide | Minimal disease activity |
| P84 | Prednisone, antibiotics | Prednisone | Prednisone | NA | NA | NA |
| P85 | Methylprednisolone, antibiotics | Prednisone | Prednisone | Partial response | Prednisone | Partial response |
| P86 | Prednisone, antibiotics, levothyroxine, CTX | Prednisone, CTX → tacrolimus, HCQ, belimumab | Prednisone, CTX → tacrolimus, HCQ, belimumab | Partial response | Prednisone, tacrolimus, HCQ, belimumab | Partial response |
| P87 | Prednisone, thalidomide, adalimumab | Prednisone, thalidomide, adalimumab | Prednisone, thalidomide, adalimumab | Minimal disease activity | Prednisone, thalidomide, adalimumab | Minimal disease activity |

**Supplementary Table 8** **(continued).**

| Cases | Treatment before genetic diagnosis | Initial treatment post-genetic diagnosis | Treatment at maximal response | Maximal response | Treatment at the most recent follow up | Clinical response at the most recent follow up |
| --- | --- | --- | --- | --- | --- | --- |
| P88  (P87's father) | NA | NA | NA | NA | NA | NA |
| P89  (P46's father) | NA | NA | NA | NA | NA | NA |
| P90 | ─ | Prednisone, adalimumab, azathioprine | Adalimumab, azathioprine | Minimal disease activity | Adalimumab, azathioprine, colchicine | Minimal disease activity |
| P91 | ─ | Adalimumab, azathioprine, colchicine | Adalimumab, colchicine | Minimal disease activity | Adalimumab | Minimal disease activity |
| P92 | ─ | Prednisone, adalimumab | Adalimumab | Minimal disease activity | Adalimumab | Minimal disease activity |
| P93 | ─ | Prednisone, adalimumab, mesalazine | Adalimumab, MTX | Minimal disease activity | MTX | Minimal disease activity |
| P94 (P93's mother) | NA | NA | NA | NA | NA | NA |

**Supplementary Table 8 (continued).**

| Cases | Treatment before genetic diagnosis | Initial treatment post-genetic diagnosis | Treatment at maximal response | Maximal response | Treatment at the most recent follow up | Clinical response at the most recent follow up |
| --- | --- | --- | --- | --- | --- | --- |
| P95 | ─ | Prednisone, adalimumab, MTX | Adalimumab, MTX | Minimal disease activity | Adalimumab, MTX | Minimal disease activity |
| P96 | NA | NA | NA | NA | NA | NA |
| P97 (P96's father) | NA | NA | NA | NA | NA | NA |
| P98 | ─ | Prednisone, thalidomide | Prednisone, thalidomide, MTX | Minimal disease activity | Thalidomide, MTX | Minimal disease activity |
| P99 | ─ | Thalidomide | Thalidomide | Minimal disease activity | Thalidomide | Minimal disease activity |
| P100 (P99s father) | NA | NA | NA | NA | NA | NA |
| P101 | Prednisone, etanercept, MMF | Prednisone, etanercept | Prednisone, MTX | Minimal disease activity | MTX, etanercept | Minimal disease activity |
| P102 | NA | NA | NA | NA | NA | NA |
| P103 | NA | NA | NA | NA | NA | NA |
| P104 | ─ | Prednisone, HCQ, MMF | Prednisone, HCQ, MMF | Partial response | HCQ, MMF | Minimal disease activity |

**Supplementary Table 8 (continued).**

| Cases | Treatment before genetic diagnosis | Initial treatment post-genetic diagnosis | Treatment at maximal response | Maximal response | Treatment at the most recent follow up | Clinical response at the most recent follow up |
| --- | --- | --- | --- | --- | --- | --- |
| P105 | Prednisone | Prednisone, adalimumab | Adalimumab | Minimal disease activity | Adalimumab | Minimal disease activity |
| P106 | NA | NA | NA | NA | NA | NA |
| P107 | Prednisone, infliximab, mesalazine, MTX, thalidomide | Thalidomide | Thalidomide | Minimal disease activity | Adalimumab | Minimal disease activity |
| P108 | Ursodesoxycholic acid | Etanercept, ursodesoxycholic acid | Etanercept, ursodesoxycholic acid | Minimal disease activity | Etanercept, ursodesoxycholic acid | Minimal disease activity |
| P109 | Methylprednisolone, IVIG | Prednisone, levetiracetam | Prednisone,  etanercept | Minimal disease activity | Prednisone, etanercept | Minimal disease activity |
| P110 | NA | NA | NA | NA | NA | NA |
| P111 | NA | NA | NA | NA | NA | NA |
| P112 | Antibiotics | Infliximab | Infliximab | Partial response | Infliximab | Minimal disease activity |
| P113 | NA | NA | NA | NA | NA | NA |
| P114 | NA | NA | NA | NA | NA | NA |
| P115 | NA | NA | NA | NA | NA | NA |
| P116 | Antibiotics | ─ | ─ | ─ | ─ | ─ |

**Supplementary Table 8 (continued).**

| Cases | Treatment before genetic diagnosis | Initial treatment post-genetic diagnosis | Treatment at maximal response | Maximal response | Treatment at the most recent follow up | Clinical response at the most recent follow up |
| --- | --- | --- | --- | --- | --- | --- |
| P117 | Antibiotics, levothyroxine | Tofacitinib | Tofacitinib | Minimal disease activity | Tofacitinib | Partial response |
| P118 | NA | ─ | ─ | ─ | ─ | ─ |
| P119 | Antibiotics, NSAIDs, methylprednisolone, Autologous hematopoietic stem cell transplant, MMF, etanercept, cyclophosphamide, thalidomide, rituximab, infliximab, MTX, adalimumab, azathioprine | Anakinra, prednisone | (1) MMF, prednisone --> lost remission after 2-3 years  (2) Anakinra, AZA, prednisone --> lost remission after 6 years | Minimal disease activity | Post allo-HSCT | Minimal disease activity |
| P120 | Antibiotics | NA | Anakinra | Minimal disease activity | Etanercept | No response |
| P121 | Prednisone, acyclovir, colchicine, etanercept | HCQ, Cyclosporine, ibuprofen | Anakinra, tofacitinib | Minimal disease activity | Tofacitinib | Minimal disease activity |

**Supplementary Table 8 (continued).**

| Cases | Treatment before genetic diagnosis | Initial treatment post-genetic diagnosis | Treatment at maximal response | Maximal response | Treatment at the most recent follow up | Clinical response at the most recent follow up |
| --- | --- | --- | --- | --- | --- | --- |
| P122 | Antibiotics, cetirizine | Prednisone | Cyclosporine | Minimal disease activity | Cyclosporine | Minimal disease activity |
| P123 | Prednisone, colchicine | Prednisone | Tofacitinib | Minimal disease activity | Upadacitinib, prednisone | Minimal disease activity |
| P124 | Cyclosporine, azathioprine, prednisone, infliximab | Infliximab | Infliximab | Minimal disease activity | Infliximab | Minimal disease activity |
| P125 | MTX, leflunomide, naproxen | MTX, naproxen | Etanercept | Minimal disease activity | MTX, naproxen | Partial response |
| P126 | Etanercept, prednisone | Infliximab | Infliximab | Minimal disease activity | Infliximab | Minimal disease activity |
| P127 | Prednisone, colchicine, mercaptopurine | Colchicine | Colchicine, etanercept, levothyroxine | Partial response | Colchicine, etanercept, levothyroxine | Partial response |
| P128 | Prednisone | Methylprednisolon → prednisone | Azathioprine, prednisolone | Minimal disease activity | Azathioprine, prednisolone | Minimal disease activity |

**Supplementary Table 8 (continued).**

| Cases | Treatment before genetic diagnosis | Initial treatment post-genetic diagnosis | Treatment at maximal response | Maximal response | Treatment at the most recent follow up | Clinical response at the most recent follow up |
| --- | --- | --- | --- | --- | --- | --- |
| P129 | Antibiotics, NSAIDs, prednisone, rituximab, etoposide, doxorubicin, vincristine, cyclophosphamide, pelfilgrastim | NA | Anakinra | Minimal disease activity | Adalimumab, budesonide | Partial response |
| P130 | Prednisone, NSAID | Colchicine | Canakinumab | Partial response | Canakinumab, colchicine | Partial response |
| P131 | Prednisone, HCQ, MTX, etanercept, azathioprine, tofacitinib | NA | Azathioprine, HCQ, rituximab, colchicine, prednisone | Partial response | Azathioprine, celecoxib, HCQ, IVIG, prednisone, rituximab | Minimal disease activity |
| P132 | HCQ; Pulses of methylprednisolone; anakinra | Prednisone | Tacrolimus, MMF, prednisone, HCQ | Minimal disease activity | Tacrolimus, MMF | Minimal disease activity |
| P133 | Prednisone, colchicine, azathioprine, HCQ, etanercept, pentoxifylline, adalimumab | Prednisone | Canakinumab, prednisone | Minimal disease activity | Canakinumab, colchicine, anakinra PRN | Minimal disease activity |

**Supplementary Table 8 (continued).**

| Cases | Treatment before genetic diagnosis | Initial treatment post-genetic diagnosis | Treatment at maximal response | Maximal response | Treatment at the most recent follow up | Clinical response at the most recent follow up |
| --- | --- | --- | --- | --- | --- | --- |
| P134 | Antibiotics, prednisone, colchicine | Colchicine, prednisone | Canakinumab | Minimal disease activity | Canakinumab | Minimal disease activity |
| P135 | Colchicine | Colchicine | Colchicine | Minimal disease activity | Colchicine | Minimal disease activity |
| P136 | ASA, methylprednisolone, prednisone | Colchicine, adalimumab | Anakinra, roflumilast | Minimal disease activity | Anakinra, roflumilast | Minimal disease activity |
| P137 | ASA, clopidogrel | Clopidogrel, eliquis | Anakinra, roflumilast | Minimal disease activity | Anakinra | Minimal disease activity |

**Supplementary Table 8 (continued).**

| Cases | Treatment before genetic diagnosis | Initial treatment post-genetic diagnosis | Treatment at maximal response | Maximal response | Treatment at the most recent follow up | Clinical response at the most recent follow up |
| --- | --- | --- | --- | --- | --- | --- |
| P138 | MTX, infliximab, etanercept, HCQ, abatacept, rituximab, adalimumab, cosetyx, otezla, rinvoq, colchicine + HCQ | No immunosuppressives | Canakinumab, anakinra PRN | Minimal disease activity | Canakinumab, anakinra PRN, roflumilast | Minimal disease activity |
| P139 | NA | Azathioprine, prednisone | Tofacitinib, azathioprine, prednisolone | Minimal disease activity | Tofacitinib, azathioprine, prednisolone | Minimal disease activity |
| P140 | Azathioprine, prednison, IVIG | Azathioprine, prednisone | Azathioprine, prednisone | Minimal disease activity | Azathioprine, canakinumab, prednisone | Minimal disease activity |
| P141 | None | Colchicine | Canakinumab | Minimal disease activity | Canakinumab | Minimal disease activity |
| P142 | HCQ | Tofacitinib | Canakinumab | Partial response | Canakinumab | Partial response |
| P143 | Ibuprofen | Ibuprofen | Tofacitinib, anakinra | Minimal disease activity | Anakinra | Minimal disease activity |

**Supplementary Table 8 (continued).**

| Cases | Treatment before genetic diagnosis | Initial treatment post-genetic diagnosis | Treatment at maximal response | Maximal response | Treatment at the most recent follow up | Clinical response at the most recent follow up |
| --- | --- | --- | --- | --- | --- | --- |
| P144 | Prednisone, mesalazine, colchicine | Apremilast | Apremilast, canakinumab | Minimal disease activity | Canakinumab, apremilast, MTX | Partial response |
| P145 | NA | Tofacitinib | Baricitinib | Minimal disease activity | Baricitinib | Minimal disease activity |
| P146 | NA | NA | NA | NA | NA | NA |
| P147 | NA | NA | NA | NA | NA | NA |
| P148 | NA | NA | NA | NA | NA | NA |
| P149 | NA | NA | NA | NA | NA | NA |
| P150 | NA | NA | NA | NA | NA | NA |
| P151 | NA | NA | NA | NA | NA | NA |
| P152 | Prednisone, rituximab | Methylprednisolone pulse → prednisone, rituximab, anakinra | Rituximab | NA | Prednisone, IVIG, ASA | NA |
| P153 | Colchicine, antibiotics, AZA | Colchicine, prednisone, AZA | Colchicine, prednisone, AZA | Partial response | Colchicine, prednisone, AZA | Partial response |
| P154 | Antibiotics | Anakinra | Anakinra | Minimal disease activity | Anakinra | Minimal disease activity |

**Supplementary Table 8 (continued).**

| Cases | Treatment before genetic diagnosis | Initial treatment post-genetic diagnosis | Treatment at maximal response | Maximal response | Treatment at the most recent follow up | Clinical response at the most recent follow up |
| --- | --- | --- | --- | --- | --- | --- |
| P155 | Prednisone, colchicine,  sulfasalazine | Prednisone, colchicine, infliximab | Prednisone, colchicine, MTX, adalimumab | Minimal disease activity | Prednisone, colchicine, MTX, adalimumab | Partial response |
| P156 | NA | NA | NA | NA | NA | NA |
| P157 | NA | NA | NA | NA | NA | NA |
| P158 | None | Etanercept | Colchicine, infliximab, prednisone; anakinra | No response | HCQ | No response |
| P159 | None | Colchicine, anakinra | Etanercept, anakinra | Partial response | Etanercept | Partial response |
| P160 | NA | MMF | MMF | Partial response | MMF | Partial response |
| P161 | NA | NA | NA | NA | NA | NA |
| P162 | Colchicine | Colchicine | Colchicine | Minimal disease activity | Colchicine | Minimal disease activity |
| P163 | Colchicine | Colchicine | Colchicine | Minimal disease activity | Colchicine | Minimal disease activity |

**Supplementary Table 8 (continued).**

| Cases | Treatment before genetic diagnosis | Initial treatment post-genetic diagnosis | Treatment at maximal response | Maximal response | Treatment at the most recent follow up | Clinical response at the most recent follow up |
| --- | --- | --- | --- | --- | --- | --- |
| P164 | Colchicine, apremilast | Apremilast | Apremilast | Minimal disease activity | Apremilast | Minimal disease activity |
| P165 | Colchicine, etanercept, infliximab, MMF, kevzara (anti-IL6) | Methylprednisolone, anakinra, IVIG | Methylprednisolone, anakinra, IVIG | Partial response | Methylprednisolone, anakinra, IVIG, colchicine | Partial response |
| P166 | HCQ | HCQ, MMF | colchicine, anakinra | Partial response | Anakinra, colchicine | Partial response |
| P167 | Colchicine,  naproxen | Colchicine, ibuprofen | Colchicine, ibuprofen | Minimal disease activity | Colchicine, ibuprofen | Minimal disease activity |
| P168 | NA | NA | NA | NA | Colchicine | NA |
| P169 | NA | NA | NA | NA | NA | NA |
| P170 | Fludrocortisone, levothyroxine, hydrocortisone | Fludrocortisone, levothyroxine, prednisone, colchicine | Fludrocortisone, levothyroxine, prednisone, anakinra | Minimal disease activity | Fludrocortisone, levothyroxine, prednisone, anakinra | Minimal disease activity |
| P171 | Antibiotics, ibuprofen | Colchicine | Prednisone | Partial response | Canakinumab | Partial response |

**Supplementary Table 8 (continued).**

| Cases | Treatment before genetic diagnosis | Initial treatment post-genetic diagnosis | Treatment at maximal response | Maximal response | Treatment at the most recent follow up | Clinical response at the most recent follow up |
| --- | --- | --- | --- | --- | --- | --- |
| P172 | NA | NA | NA | NA | NA | NA |
| P173 | Azathioprine, prednisone, sildenafil, furosemide | NA | Azathioprine, prednisone, sildenafil, furosemide | Partial response | Azathioprine, prednisone, sildenafil, furosemide | Partial response |
| P174 | NA | NA | NA | NA | NA | NA |
| P175 | NA | NA | NA | NA | NA | NA |
| P176 | NA | Anakinra | Canakinumab | Minimal disease activity | Canakinumab | Minimal disease activity |
| P177 | NA | NA | NA | NA | NA | NA |
| P178 | NA | IVIG, MMF | Methylprednisolone, IVIG, MMF | Partial response | IVIG, MMF | Disease flare |
| P179 | NA | Colchicine | Colchicine | Partial response | Colchicine | Partial response |

**Supplementary Table 8 (continued).**

| Cases | Treatment before genetic diagnosis | Initial treatment post-genetic diagnosis | Treatment at maximal response | Maximal response | Treatment at the most recent follow up | Clinical response at the most recent follow up |
| --- | --- | --- | --- | --- | --- | --- |
| P180 | MMF, HCQ daily, prednisone, belimumab, tacrolimus, anifrolumab | Anakinra | Anakinra | Partial response | Anakinra | Partial response to disease flare |
| P181 | HCQ, MMF, anakinra, anifrolumab | Colchicine | Colchicine | Partial response | Colchicine | Partial response to disease flare |
| P182 | Prednisone, colchicine | Anakinra | Anakinra | Partial response | Anakinra | Partial response |
| P183 | NA | Colchicine | Adalimumab | NA | Adalimumab | NA |
| P184 | Antibiotics | NA | Colchicine, Adalimumab | Minimal disease activity | Colchicine, adalimumab | Minimal disease activity |
| P185 | Aciclovir, valganciclovir, ganciclovir | NA | Colchicine | Partial response | Colchicine | Partial response |

MTX: methotrexate; MMF: mycophenolate mofetil; IVIG: intravenous immunoglobulin; CTX: cyclophosphamide; HCQ, hydroxychloroquine; NSAIDs: non-steroidal anti-inflammatory drugs; NA: data not available or patient lost to follow-up.

**Supplementary Table 9 The durability of clinical response in HA20 patients.**

| Cases | Follow-up (months) | Longest duration, minimal disease activity（months) | Disease flares during follow-up (number) | Triggers for disease flares | | | |
| --- | --- | --- | --- | --- | --- | --- | --- |
|  |  |  |  | Infection (number) | Drugs tapering | Anti-TNFi  antibodies (number) | Others |
| P1 | 48 | 37.2 | 0 |  |  |  |  |
| P2 | 54 | 14.4 | 2 |  |  | 2 |  |
| P3 | 37 | 16.8 | 2 |  |  | 2 |  |
| P5 | 43 | 7.2 | 1 |  | GCs discontinuation |  |  |
| P7 | 47 | 44.4 | 1 |  |  |  | Infusion reaction while on infliximab, triggering autoinflammation |
| P37 | 48 | 34 | 0 |  |  |  |  |
| P38 | 72 | 10 | 2 |  | GCs Tapering |  |  |
| P39 | 104 | 104 | 0 |  |  |  |  |
| P40 | 51 | 13 | 0 |  |  |  |  |
| P41 | 6 | 6 | 0 |  |  |  |  |
| P42 | 60 | 24 | 2 | 2 |  |  |  |
| P43 | 18 | 26 | 3 | 3 |  |  |  |
| P44 | 20 | 15 | 0 |  |  |  |  |
| P61 | 42 | 4 | 3 | 2 |  | 1 |  |
| P62 | 19 | 7 | 4 |  | GCs Tapering | 1 |  |

**Supplementary Table 9 (continued).**

| Cases | Follow-up (months) | Longest duration, minimal disease activity（months) | Disease flares during follow-up (number) | Triggers for disease flares | | | |
| --- | --- | --- | --- | --- | --- | --- | --- |
|  |  |  |  | Infection (number) | Drugs tapering | Anti-TNFi  antibodies (number) | Others |
| P104 | 15 | 8.4 | 0 |  |  |  |  |
| P107 | 16 | 12 | 0 |  |  |  |  |
| P112 | 13 | 13.2 | 0 |  |  |  |  |
| P136 | 24 | 8 | 2 | 2 |  |  |  |
| P137 | 30 | 24 | 0 |  |  |  |  |
| P144 | 27 | 17 | 1 |  |  |  | Unknown |
| P145 | 44 | 22 | 1 |  | Tof discontinuation |  |  |

GCs: glucocorticoids; Tof: tofacitinib.

**Supplementary Table 10 Adverse events and outcome of HA20 patients.**

| Cases | Follow-up (months) | Adverse events | | | Survival |
| --- | --- | --- | --- | --- | --- |
|  |  | Infection | Allergic reaction | Others |  |
| P1 | 48 | URI, three times | No | No | Yes |
| P2 | 54 | URI, twice; acute enteritis, twice | No | Constipation | Yes |
| P3 | 37 | URI, four times | No | No | Yes |
| P4 | 25 | URI, three times | No | No | Yes |
| P5 | 43 | URI, twice | No | No | Yes |
| P6 (P5's mother) | 43 | No | No | No | Yes |
| P7 | 47 | URI, twice | No | Infusion reaction while on infliximab | Yes |
| P8 (P7's father) | 26 | No | No | No | Yes |
| P9 | 26 | No | No | No | Yes |
| P10 | 62 | No | No | No | Yes |
| P11 | 36 | No | Drug eruptions | No | Yes |
| P12 | 15 | No | No | No | Yes |
| P13 | 64 | No | No | Neuromuscular injury | Yes |
| P14 | 3 | URI, three times | No | No | Yes |
| P15 | NA | ─ | ─ | ─ | ─ |
| P16 (P17's mother) | 241 | No | No | Acoustic neuroma, depression and anxiety | Yes |
| P17 | NA | ─ | ─ | ─ | ─ |
| P18 | 100 | UTI, once | No | No | Yes |
| P19 (P18's son) | 38 | No | No | No | Yes |
| P20 (P18's mother) | NA | NA | NA | NA | NA |
| P21 | 7 | URI, twice | No | No | Yes |
| P22 | 21 | URI, twice | No | No | Yes |
| P23 (P22's mother) | 21 | URI, three times | No | No | Yes |
| P24 | 15 | No | No | No | Yes |
| P25 | 34 | No | No | No | Yes |
| P26 | 83 | Intestinal infection, once | No | Peripheral nerve injury | Yes |

**Supplementary Table 10 (continued).**

| Cases | Follow-up (months) | Adverse events | | | Survival |
| --- | --- | --- | --- | --- | --- |
|  |  | Infection | Allergic reaction | Others |  |
| P27 (P24's mother) | 15 | No | No | No | Yes |
| P28 | 39 | URI, five times; intestinal infection, three times | No | No | Yes |
| P29 | 42 | No | No | Facial neuritis, vomiting | Yes |
| P30 | 41 | Pneumonia, twice; bronchitis, once; acute enteritis, three times; chronic peridontitis | No | No | Yes |
| P31 | 43 | Anorectal infection, once; cervical lymphadenitis, once; submaxillary cellulitis, once | No | No | Yes |
| P32 | 18 | Cervical lymphadenitis, twice | No | HLH secondary to EBV infection | Yes |
| P33 | 20 | URI, three times | No | No | Yes |
| P34 | 1 | No | No | No | Yes |
| P35 | 69 | URI, once | No | No | Yes |
| P36 | 12 | No | No | Abdominal pain | Yes |
| P37 | 48 | No | No | No | Yes |
| P38 (P45's daughter) | 72 | No | No | No | Yes |
| P39 (P45's son) | 104 | No | No | No | Yes |
| P40 (P46's daughter) | 51 | No | No | No | Yes |
| P41 (P46's son) | 6 | No | No | No | Yes |
| P42 | 60 | URI, twice | No | No | Yes |
| P43 | 18 | Pneumonia, once | No | No | No |
| P44 | 20 | No | No | No | Yes |

**Supplementary Table 10 (continued).**

| Cases | Follow-up (months) | Adverse events | | | Survival |
| --- | --- | --- | --- | --- | --- |
|  |  | Infection | Allergic reaction | Others |  |
| P45 | NA | NA | NA | NA | NA |
| P46 | NA | NA | NA | NA | NA |
| P47 (P36's father) | NA | NA | NA | NA | NA |
| P48 | 6 | No | No | No | Yes |
| P49 | 5 | No | No | No | Yes |
| P50 (P49's mother) | NA | NA | NA | NA | NA |
| P51 | 23 | No | No | No | Yes |
| P52 | 20 | No | No | No | Yes |
| P53 | 94 | No | No | No | Yes |
| P54 | 21 | No | No | No | Yes |
| P55(P54's mother) | 33 | No | No | Dizziness and vomiting | Yes |
| P56 | 2 | Pneumonia, once | No | No | Yes |
| P57 | 60 | No | No | No | Yes |
| P58 (P33's father) | 20 | No | No | No | Yes |
| P59 (P51’s father) | NA | NA | NA | NA | NA |
| P60 | 41 | URI, once; pneumonia, once | No | No | Yes |
| P61 | 42 | URI, seven times (Sinusitis, six times; acute tonsillitis, once); pneumonia, once | No | No | Yes |
| P62 | 19 | No | No | Subcutaneous edema | Yes |
| P63 | 3 | No | No | No | Yes |
| P64 | NA | NA | NA | NA | NA |
| P65 | 0.3 | No | No | No | Yes |
| P66 (P65's mother) | 0.3 | No | No | No | Yes |
| P67 | 31 | No | No | No | Yes |

**Supplementary Table 10 (continued).**

| Cases | Follow-up (months) | Adverse events | | | Survival |
| --- | --- | --- | --- | --- | --- |
|  |  | Infection | Allergic reaction | Others |  |
| P68 (P67's brother) | 31 | URI, three times; parotiditis, once | No | No | Yes |
| P69 (P67's father) | 31 | No | No | No | Yes |
| P70 | NA | NA | NA | NA | NA |
| P71 (P63's daughter) | NA | NA | NA | NA | NA |
| P72 (P64's daughter) | NA | NA | NA | NA | NA |
| P73 | 36 | No | Drug eruptions | No | Yes |
| P74 (P73's father) | NA | NA | NA | NA | NA |
| P75 | NA | NA | NA | NA | NA |
| P76 | 40 | Respiratory infection, ten times | Injection reaction during infliximab transfusion | No | Yes |
| P77 | 26 | URI, four times; pneumonia, twice | No | No | Yes |
| P78 | 3 | URI, once; hand-foot-mouth disease, once | No | No | Yes |
| P79 | 13 | Respiratory infection, once | No | No | Yes |
| P80 | NA | NA | NA | NA | NA |
| P81 | 24 | No | No | No | Yes |
| P82 | NA | NA | NA | NA | NA |
| P83 | 2 | No | No | No | Yes |
| P84 | NA | NA | NA | NA | NA |
| P85 | 6 | Respiratory infection, once | NA | NA | NA |
| P86 | 4 | No | No | No | Yes |
| P87 | 5 | No | No | No | Yes |

**Supplementary Table 10 (continued).**

| Cases | Follow-up (months) | Adverse events | | | Survival |
| --- | --- | --- | --- | --- | --- |
|  |  | Infection | Allergic reaction | Others |  |
| P88 (P87's father) | NA | NA | NA | NA | NA |
| P89 (P46's father) | NA | NA | NA | NA | NA |
| P90 | 1 | No | No | No | Yes |
| P91 | 7 | No | No | No | Yes |
| P92 | 9 | No | No | No | Yes |
| P93 | 6 | No | No | No | Yes |
| P94 (P93's mother) | NA | NA | NA | NA | NA |
| P95 | 12 | No | No | No | Yes |
| P96 | NA | NA | NA | NA | NA |
| P97 (P96's father) | NA | NA | NA | NA | NA |
| P98 | 32 | URI, ten times | No | No | Yes |
| P99 | 36 | URI, ten times; pneumonia, three times | No | No | Yes |
| P100 (P99's father) | NA | NA | NA | NA | NA |
| P101 | NA | NA | NA | NA | NA |
| P102 | NA | NA | NA | NA | NA |
| P103 | NA | NA | NA | NA | NA |
| P104 | 15 | Salmonella infection, once | No | No | Yes |
| P105 | 48 | Pneumonia, twice | No | No | Yes |
| P106 | NA | NA | NA | NA | NA |
| P107 | 16 | No | Yes | No | Yes |
| P108 | 30 | No | No | No | Yes |
| P109 | 30 | No | No | No | Yes |
| P110 | NA | NA | NA | NA | NA |
| P111 | 8 | No | No | No | Yes |
| P112 | 13 | No | No | No | Yes |
| P113 | 8 | No | No | No | Yes |
| P114 | 8 | No | No | No | Yes |

**Supplementary Table 10 (continued).**

| Cases | Follow-up (months) | Adverse events | | | Survival |
| --- | --- | --- | --- | --- | --- |
|  |  | Infection | Allergic reaction | Others |  |
| P115 | 8 | No | No | No | Yes |
| P116 | 8 | No | No | No | Yes |
| P117 | 0.17 | No | No | No | Yes |
| P118 | 2.05 | No | No | No | No |
| P119 | 0.45 | Yes | Urticarial rash when on rituximab | Headaches while  on IVIG | Yes |
| P120 | 1.48 | No | No | No | Yes |
| P121 | 0.73 | Pneumonia, once | No | No | Yes |
| P122 | 0.08 | No | Injection reaction when on anakinra | Aplastic anemia while on azathioprine | Yes |
| P123 | 0.43 | Multiple infections (urinary and respiratory) | No | Erratic mood swings and emotional instability while on colchicine | Yes |
| P124 | 156 | NA | NA | NA | NA |
| P125 | 0.71 | No | No | No | Yes |
| P126 | 1 | No | No | No | Yes |
| P127 | 0.21 | NA | NA | Elevated transaminases while on colchcine; cytopenias while on mercaptopurine | NA |
| P128 | 0.32 | NA | NA | NA | Yes |
| P129 | 1.23 | No | No | Systemic adverse events attributed to budesonide; headaches and fevers while on canakinumab | Yes |
| P130 | 0.02 | Influenza B infection | Injection reaction when on Canakinumab | GI symptoms while on colchicine | Yes |

**Supplementary Table 10 (continued).**

| Cases | Follow-up (months) | Adverse events | | | Survival |
| --- | --- | --- | --- | --- | --- |
|  |  | Infection | Allergic reaction | Others |  |
| P131 | 0.49 | Sepsis due to bacterial pneumonia, Complicated acute pyelonephritis, otitis media and other upper respiratory tract infection, pneumonia due to COVID infection | No | No | Yes |
| P132 | 0.03 | No | No | No | Yes |
| P133 | 1.37 | No | Two episodes of blisters, weakness, fatigue, myalgias while on etanercept | No | Yes |
| P134 | 0.21 | No | Injection reaction while on anakinra | No | Yes |
| P135 | 0.16 | No | No | No | Yes |
| P136 | 24 | No | No | No | Yes |
| P137 | 30 | No | No | No | Yes |
| P138 | 6 | Salmonella infection while on adalimumab | No | Leukopenia, mouth ulcers worse, diarrhea, dizziness | Yes |
| P139 | 0.25 | No | No | No | Yes |
| P140 | 135.5 | No | Injection reaction while on Anakinra | Osteoporosis, Fatigue, weight gain, and hair loss while on JAK inhibitor | Yes |
| P141 | 11 | NA | No | No | Yes |
| P142 | NA | NA | No | No | Yes |
| P143 | 1.78 | No | Facial rashes while on tofacitinib | No | Yes |
| P144 | 27 | No | Injection reaction while on anakinra | GI symptoms | Yes |

**Supplementary Table 10 (continued).**

| Cases | Follow-up (months) | Adverse events | | | Survival |
| --- | --- | --- | --- | --- | --- |
|  |  | Infection | Allergic reaction | Others |  |
| P145 | 44 | No | No | GI symptoms and BK viral reactivation while on JAK inhibitor | Yes |
| P146 | NA | NA | NA | NA | NA |
| P147 | NA | NA | NA | NA | NA |
| P148 | NA | NA | NA | NA | NA |
| P149 | NA | NA | NA | NA | NA |
| P150 | NA | NA | NA | NA | NA |
| P151 | NA | NA | NA | NA | NA |
| P152 | NA | No | No | Blindness due to thrombocytopenia and intraocular bleeding; many social problems | Yes |
| P153 | 86 | NA | NA | NA | NA |
| P154 | NA | NA | NA | NA | NA |
| P155 | 24 | Pharyngitis monthly | NA | NA | Yes |
| P156 | NA | NA | NA | NA | NA |
| P157 | NA | NA | NA | NA | NA |
| P158 | 10 | No | No | Global amnesia and fainting spells while on duloxetine | Yes |
| P159 | 10 | NA | NA | Fatigue while on anakinra; GI upset while on colchicine | NA |
| P160 | 57 | NA | NA | GI upset and myalgia while on colchicine | NA |
| P161 | 0.05 | No | No | No | Yes |
| P162 | 1.76 | No | No | No | Yes |
| P163 | 0.16 | No | No | No | Yes |
| P164 | 4 | No | No | No | Yes |
| P165 | 1.23 | UTIs | Rash while on MMF and MTX | Injection site reactions while on abatacept and golimumab; infusion reaction while on infliximab | Yes |
| P166 | 1.45 | No | Hemolytic anemia while on IVIG 2g/kg | No | Yes |
| P167 | 0.11 | No | No | Diarrhea while on colchicine | Yes |
| P168 | 1 | No | No | No | Yes |

**Supplementary Table 10 (continued).**

| Cases | Follow-up (months) | Adverse events | | | Survival |
| --- | --- | --- | --- | --- | --- |
|  |  | Infection | Allergic reaction | Others |  |
| P169 | NA | NA | NA | NA | NA |
| P170 | 0.91 | No | No | No | Yes |
| P171 | 1.11 | No | No | No | Yes |
| P172 | 0.59 | NA | NA | NA | NA |
| P173 | 53.87 | NA | NA | NA | Yes |
| P174 | NA | NA | NA | NA | Yes |
| P175 | NA | NA | NA | NA | Yes |
| P176 | 27 | No | Serum-sickness like reaction to amoxicillin; erythematous nodules at anakinra injection sites | No | Yes |
| P177 | 2.17 | NA | NA | NA | Yes |
| P178 | 156 | No | No | No | Yes |
| P179 | 0 | NA | NA | NA | Yes |
| P180 | 46 | NA | NA | NA | Yes |
| P181 | 9 | NA | NA | NA | Yes |
| P182 | 1 | NA | NA | NA | Yes |
| P183 | 1 | NA | NA | NA | Yes |
| P184 | 10 | No | No | No | Yes |
| P185 | 0.55 | No | No | No | Yes |

URI, upper respiratory infection; UTI, urinary tract infection; HLH, hemophagocytic lymphohistiocytosis; EBV, Epstein–Barr virus; IVIG, intravenous immunoglobulin; MMF, mycophenolate mofetil; MTX, methotrexate; JAK inhibitor, Janus kinase inhibitor; GI, gastrointestinal; COVID, coronavirus disease 2019; BK, BK polyomavirus; NA indicates data not available or patient lost to follow-up.
